# Quantitative and qualitative patient-reported analysis of misdiagnosis and/or late diagnosis of metastatic lobular cancer

**DOI:** 10.64898/2026.04.16.26348799

**Authors:** Morgan Elizabeth Cody, Hung-Ching (Rick) Chang, Julia Foldi, Rachel C. Jankowitz, Marija Balic, Tracy Cushing, Carla Donnelly, Siobhán Freeney, Julia Levine, Lori Petitti, Nancy Ryan, Kirstin Spencer, Claire Turner, George C. Tseng, Christine Desmedt, Steffi Oesterreich, Adrian V. Lee

**Affiliations:** School of Medicine, University of Pittsburgh, Pittsburgh, PA, USA; Biostatistics, University of Pittsburgh, Pittsburgh, PA, USA; UPMC Hillman Cancer Center, University of Pittsburgh Medical Center, Pittsburgh, PA, USA; Abramson Cancer Center, University of Pennsylvania, Philadelphia, PA, USA; Independent Patient Advocate, Boulder, CO, USA; Independent Patient Advocate, Pittsburgh, PA, USA; Non-profit organization, Lobular Ireland, Dublin, IRELAND; Non-profit organization, Lobular Breast Cancer Alliance, Delaware, DE, USA; Independent Patient Advocate, Los Angeles, CA, USA; Independent Patient Advocate, Portland, ME, USA; Independent Patient Advocate, Gloucestershire, UNITED KINGDOM; Non-profit organization, Lobular Breast Cancer UK, Manchester, UNITED KINGDOM; Computational & Systems Biology, University of Pittsburgh School of Public Health, Pittsburgh, PA, USA; Laboratory for Translational Breast Cancer Research, Department of Oncology, KU Leuven, Leuven, BELGIUM; European Lobular Breast Cancer Consortium, Utrecht, NETHERLANDS; Women’s Cancer Research Center UPMC Hillman Cancer Center and Magee Women’s Research Institute, University of Pittsburgh Medical Center, Pittsburgh, PA, USA; Department of Pharmacology and Chemical Biology, University of Pittsburgh School of Medicine, Pittsburgh, PA, USA; Institute for Precision Medicine, University of Pittsburgh and UPMC, Pittsburgh, PA, USA

## Abstract

**Background:** Invasive lobular breast cancer (ILC) is the most commonly diagnosed special histological subtype of breast cancer (BC). Metastatic ILC (mILC) is less sensitive to FDG-PET imaging and often metastasizes to unusual sites —peritoneum, gastrointestinal (GI) tract, ovaries, urinary tract, and orbit—which may go unrecognized after a long disease-free interval. Some metastatic sites cause nonspecific symptoms, like abdominal/epigastric pain, with numerous published case reports of mILC misdiagnosed as gastric cancer. These atypical BC metastatic sites may lead to late and/or misdiagnosis, thereby delaying effective treatments.

**Objective:** We developed a patient survey to investigate the patient-reported prevalence of delayed diagnosis or misdiagnosis of mILC and their potential impact upon treatment outcomes.

**Methods:** A 45-question survey was developed and piloted with breast cancer researchers, clinical oncologists, and patient advocates. This IRB-approved survey was then distributed to patients with ILC. Analyses including data QC and visualization were conducted in R using descriptive statistics. Incomplete or inconsistent responses were excluded, and summary statistics were stratified by four common mILC sites to highlight subgroup differences.

**Results:** 525 patient surveys were completed, with 450 patients diagnosed with ILC, and of those 321 diagnosed with mILC. For those with mILC, 33.3% (n=107) were diagnosed with *de novo* mILC at initial presentation. Of the patients diagnosed with mILC, 32.1% (n=103) presented with other medical conditions at diagnosis. Misdiagnosis was reported by 26.2% (n=84) of patients with mILC, and of these cases, 31% (n=26) had ≥2 misdiagnoses. The top 5 misdiagnoses were bone-related condition (24.7%), benign breast condition (23.4%), another type of BC (7.8%), diagnostic delay (7.8%), and menopause related (5.2%). 44.5% of patients waited ≥1 year for an accurate diagnosis. 49 patients were treated for their misdiagnosis, and 6 received incorrect cancer treatments. The most frequently reported contributors to delayed or misdiagnosis were inconclusive imaging, providers’ lack of ILC knowledge, and initial misdiagnosis. Of the 321 patients with mILC, 138 (42.9%) reported symptoms before diagnosis; the most common were back pain (16.5%), fatigue/malaise (14.9%), GI symptoms (11.8%), bloating (8.4%), and weight loss (8.1%). Although 40% of patients reported having a mammogram at the time of their initial misdiagnosis, ILC was detected in only 20.5% (24/116) of these cases, and mammography detected only 5 (25%) of the 20 de novo mILC cases. Patients reported additional diagnostic testing within 1-3 months of their initial mammogram, includingbiopsy, ultrasound (US), and MRI. 47.9% of patients were in active BC surveillance after curative intent therapy at the time of their mILC diagnosis; however, no statistical difference was seen in time to diagnosis versus those patients not under surveillance.

**Conclusion:** Our survey results underscore the urgent need to improve diagnostic strategies for mILC. Addressing delays and diagnostic errors in mILC is critical to optimizing treatment strategies and improving patient outcomes.

## Introduction

Invasive lobular breast cancer (ILC) is the most commonly diagnosed special histological subtype of breast cancer (BC) in women, after invasive breast cancer of no special type (NST), formerly called invasive ductal carcinoma (IDC). ILC accounts for up to 15% of all invasive breast cancers in women^1,2^. A recent study using United States population-based cancer registry data found rates of ILC have increased by 2.8% per year from 2012 to 2021, compared with 0.8% per year for all other breast cancers combined^3^. The last five years have seen an accumulating body of scientific and clinical evidence demonstrating that ILC has a distinct clinical, histopathological, and molecular profile from NST^2^. Distinct features of ILC include the loss of cell-cell adhesion molecule E-cadherin (*CDH1*) protein expression observed in ∼90% of ILCs^2,4,5^. ILCs are most commonly estrogen receptor (ER) and progesterone receptor (PR) positive, and HER2-negative^6^; however, they can present more aggressively as HER2-positive or triple-negative (TNBC)^7,8,9^. Compared to NSTs, ILCs are more often multicentric, multifocal, and bilateral; larger in tumor size at primary diagnosis, and more frequently diagnosed in older women, often with various degrees of lymph node involvement^10,11,12^. The diffuse growth pattern of ILC makes it difficult to detect early during routine screening by physical examination, as it often lacks a palpable mass, and standard mammography techniques have reported low sensitivity compared to NST, resulting in a potential false-negative rate of nearly 30%^2,13^.

The initial clinical prognosis of ILC is often more favorable than NST^14^, however, ILC relapse can occur more than 10 years after diagnosis of the primary tumor, largely affecting the post-menopausal segment^15,16^. Because the majority of ILC presents as ER-positive, the risk of late relapse due to endocrine treatment resistance is a concern. The International Breast Cancer Study Group (IBCSG) analyzed data on more than 13,000 patients with early BC, demonstrating a favorable outcome for ILC patients during the first decade of follow-up, but a worse outcome beyond 10 years^13^. In a more recent study by Foldi J et al evaluating recurrences and overall survival of over 12,000 patients from 4 prospective randomized phase III National Surgical Adjuvant Breast and Bowel Project (NSABP) trials found patients with ILC have elevated risk of late recurrence and death compared with patients with NST^17^. Other recent studies, comparing the disease-free survival (DFS) and overall survival (OS) of lobular and ductal carcinoma breast cancer, confirmed this pattern^16,18,19,20^.

ILC exhibits a unique pattern of metastatic spread compared to NST^21^. Metastatic invasion of the bone and liver is more frequent in patients with NST, however, next to invasion in more standard anatomical sites, ILC is associated with invasion to unique sites such as the peritoneum, gastrointestinal tract, ovaries, urinary tract, leptomeninges, skin, and orbit^22,23,24,25^. Metastases to these sites can lead to misleading symptoms, such as bowel obstruction or abdominal bloating, that are not typically associated with breast cancer. Metastases to these sites can lead to misleading symptoms, such as bowel obstruction or abdominal bloating, that are not typically associated with breast cancer. Similar to primary ILC in the breast, mILC tends to infiltrate the affected organs diffusely without forming a discrete tumor mass^26^. Understanding these challenges is critical for improving the diagnosis and treatment of ILC.

To better understand whether the unique characteristics of ILCs and their tendency to recur at unusual metastatic sites lead to misdiagnosis or delayed time to an accurate diagnosis and treatment initiation, we initially performed a literature review, which revealed that no prior studies have systematically investigated mILC misdiagnosis. However, we found twenty-four case reports that indicated the most common misdiagnosed mILC sites were to the stomach, duodenum, and colon. Symptoms such as epigastric pain and obstruction often led to misdiagnoses of primary GI conditions, and the time between initial ILC diagnosis and metastatic confirmation ranged from 0.25 to 30 years^27^.

Given the challenges associated with the distinct histopathologic and molecular differences in ILC, we developed a patient survey to investigate the patient-reported prevalence of delayed diagnosis or misdiagnosis of mILC and the impact on treatment outcomes.

## Methods

A 45-question survey to assess patients’ history, symptoms, diagnosis, and the impact of misdiagnosis or delays on treatment was developed and piloted with a team of four breast cancer researchers, three clinical oncologists, and six patient advocates. After five rounds of revisions, the final University of Pittsburgh IRB-approved English-language-only survey (STUDY24100051) was distributed from March through May 31, 2025, using the Qualtrics© platform to two physicians to their patients, a patient advocate network, and social media. The survey was distributed to patients with ILC via social media advocacy groups, ILC advocacy organizations, and ILC conferences. The research team collected responses and conducted descriptive and comparative analyses with a biostatistician. Data quality control included removal of test responses, exclusion of invalid entries, and consistency checks across related survey items. Survey responses were summarized using frequencies and proportions, with stratified analyses performed by relevant respondent characteristics. All data processing, analysis, and visualization were conducted in *R* (version 4.4.2). Incomplete or inconsistent responses were excluded, and summary statistics were stratified by four common mILC sites (gastrointestinal (GI), genitourinary (GU), neurology (neuro), hematology (hemato)) to highlight subgroup differences.

### AI-assisted Free-Text Analysis

To efficiently process and categorize open-ended survey responses, we used ChatGPT (GPT-4o) as a natural language processing assistant. Patients’ free-text answers were first reviewed and lightly preprocessed to remove irrelevant formatting. We then prompted ChatGPT to generate summarized categories and assign them to the responses. This helped organize diverse free-text inputs into structured formats such as frequency tables. When the AI-generated output included overly detailed or unclear categories, we manually refined the results through an iterative review process to ensure clinical relevance and interpretability. This approach provided an efficient and adaptable method for analyzing unstructured qualitative data. See supplementary information for more details.

## Results

### Patient Characteristics and Burden of Misdiagnosis

A total of 525 patient surveys were completed, with 450 patients diagnosed with ILC and 321 diagnosed with mILC. The majority (67%) of survey respondents in both the ILC and mILC subgroups were between 50-79 years old (see supplementary information for more details). Of those patients with mILC, 33.3% (n=107), were diagnosed with *de novo* mILC at initial presentation. Of the patients diagnosed with mILC, 32.1% (n=103), presented with other medical conditions at diagnosis. Misdiagnosis was reported by 26.2% (n=84) of patients with mILC, and 31% (n=26) had ≥2 misdiagnoses before receiving an accurate one (Figure 1).

**Figure 1.**
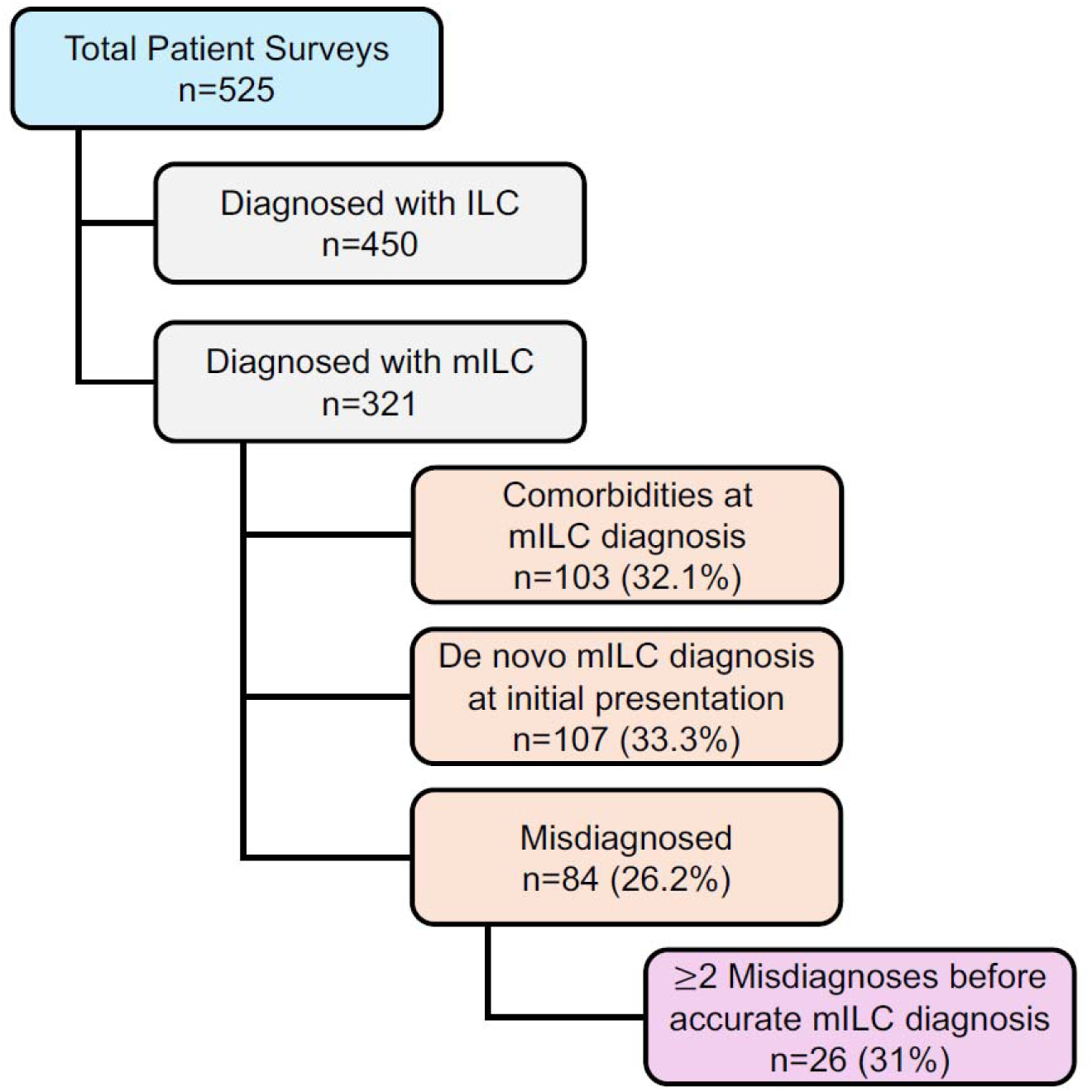
ILC Patient Survey Demographics. A total of 525 patient surveys were delivered; 450 patients were diagnosed with ILC and 321 with mILC. Of the 321 patients with mILC, 32% presented with comorbidities, 33.3% were diagnosed with de novo mILC at initial presentation, and 26.2% were misdiagnosed, with 31% reporting 2 or more misdiagnoses before receiving an accurate diagnosis.

### Patterns and Consequences of Misdiagnosis

Among the 84 patients with mILC who were misdiagnosed, proportionally, the five most common misdiagnoses reported during their diagnostic workup were bone-related condition (24.7%), benign breast condition (23.4%), another type of BC (7.8%), diagnostic delay (7.8%), and menopause related (5.3%) (Figure 2). Nearly half (44.5%) of those patients waited ≥1 year for an accurate diagnosis.

**Figure 2.**
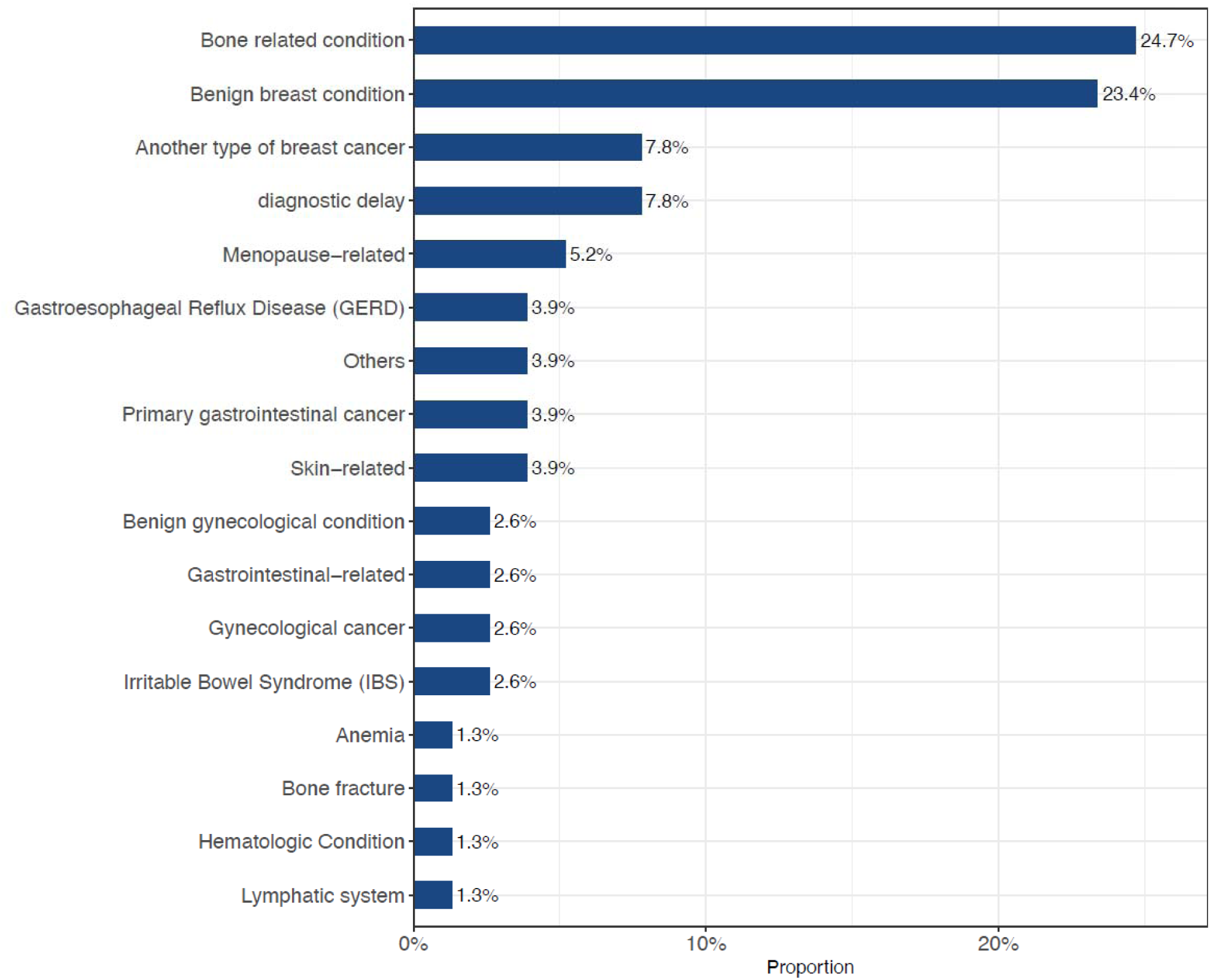
Patient Reported Misdiagnoses. Among the 321 patients with mILC, 84 (26.2%) reported receiving one or more misdiagnoses before receiving an accurate diagnosis. Proportionally, bone related condition (24.7%), and benign breast condition (23.4%) were the most common misdiagnoses reported.

Forty-seven (56%) of the 84 patients with mILC that reported a misdiagnosis also provided more details about their experiences, describing a total of 72 misdiagnoses: 23 GI, 19 GU, 15 neurological, and 15 hematological (Figure 3). This includes patients who reported >1 misdiagnosis of the same type, e.g., 17 patients had GU misdiagnosis, but the total number of GU misdiagnoses reported was 19. Over a third (38%) of those misdiagnosed provided details of their treatments, with musculoskeletal, pain, or GI therapies among the top treatments described; 6 patients received unnecessary cancer treatments (Figure 4). Diagnostic work-up for most patients included CT, biopsy, MRI, labs, and ultrasound (US).

**Figure 3.**
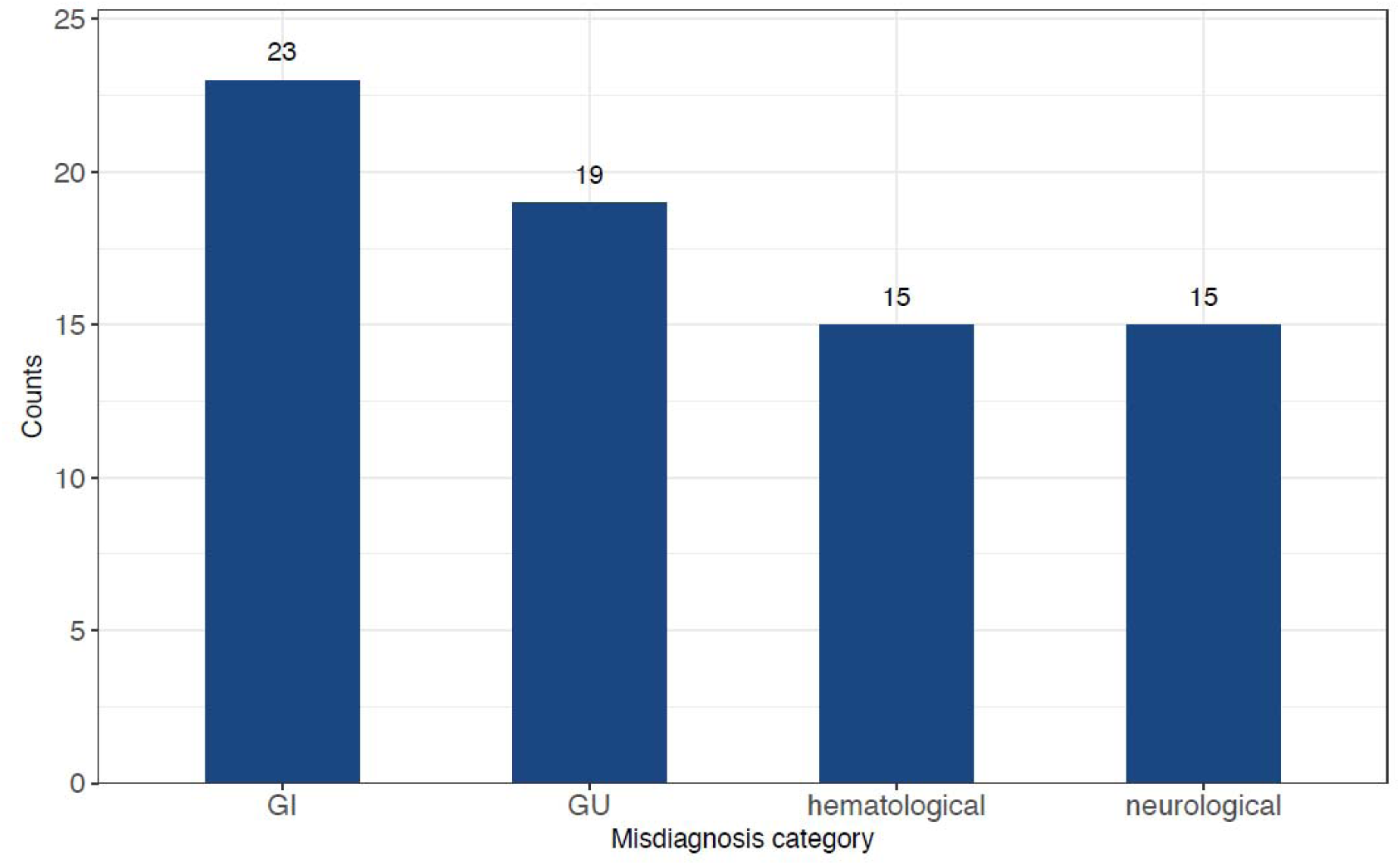
Number of patient-reported misdiagnoses by primary therapeutic subclass. 47 (56%) of the 84 patients with mILC who reported one or more misdiagnoses provided more details. They collectively identified 72 distinct types of misdiagnoses, which were categorized by the primary therapeutic subclass. For example, while 17 patients were misdiagnosed in the GU category, the total count of GU misdiagnoses reported reached 19. GI: gastrointestinal, GU: genitourinary.

**Figure 4.**
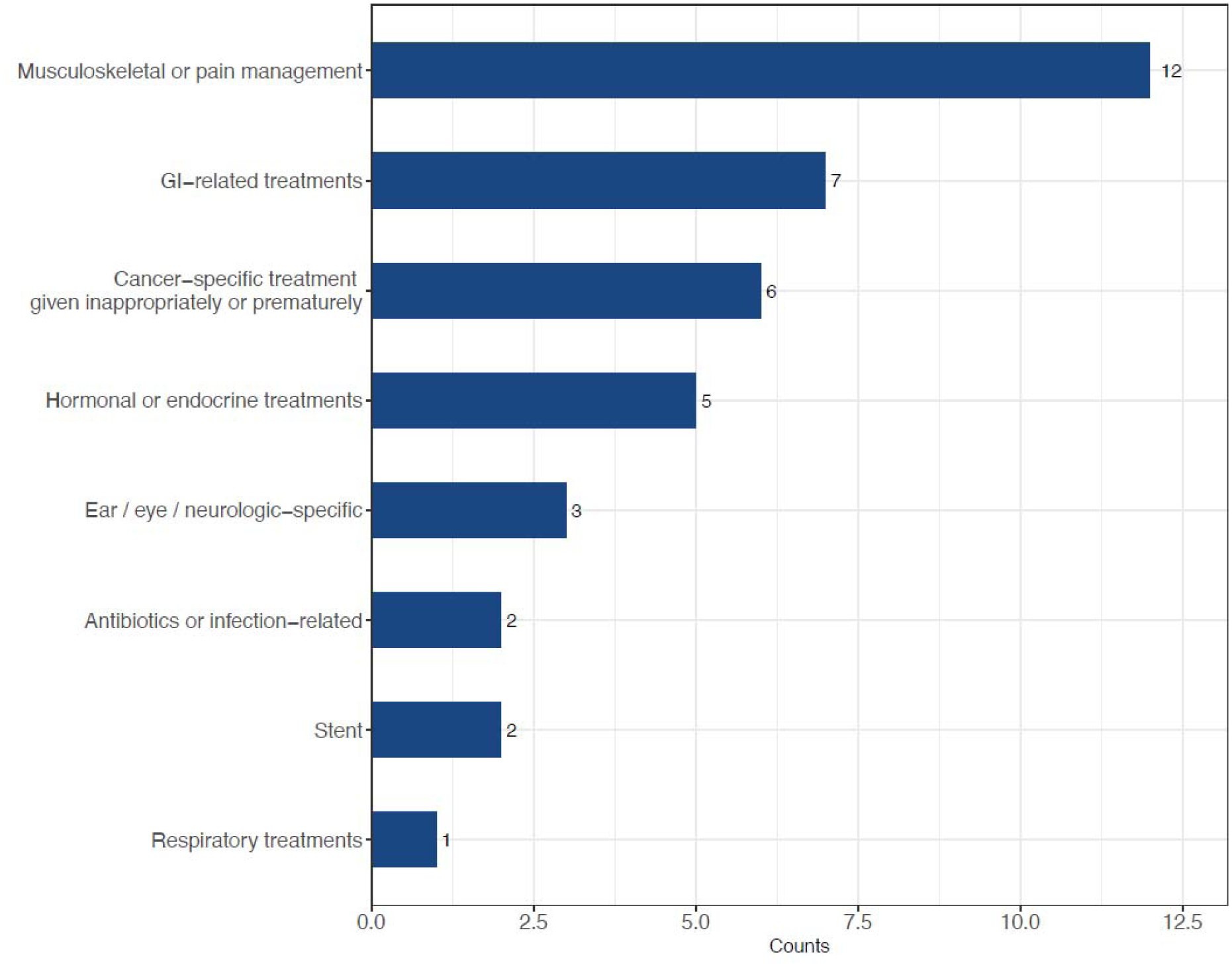
Types of Treatment Misdiagnosed Patients Received. 32 (38%) of the 84 patients with mILC who reported misdiagnoses indicated the types of treatment they were given for their misdiagnosis, including 6 (18.75%) patients who received inappropriate cancer treatments.

### Factors Contributing to Delayed or Incorrect Diagnosis

The most reported contributors to delayed or misdiagnosis were inconclusive imaging, lack of ILC knowledge by providers, and initial misdiagnosis (Figure 5).

**Figure 5.**
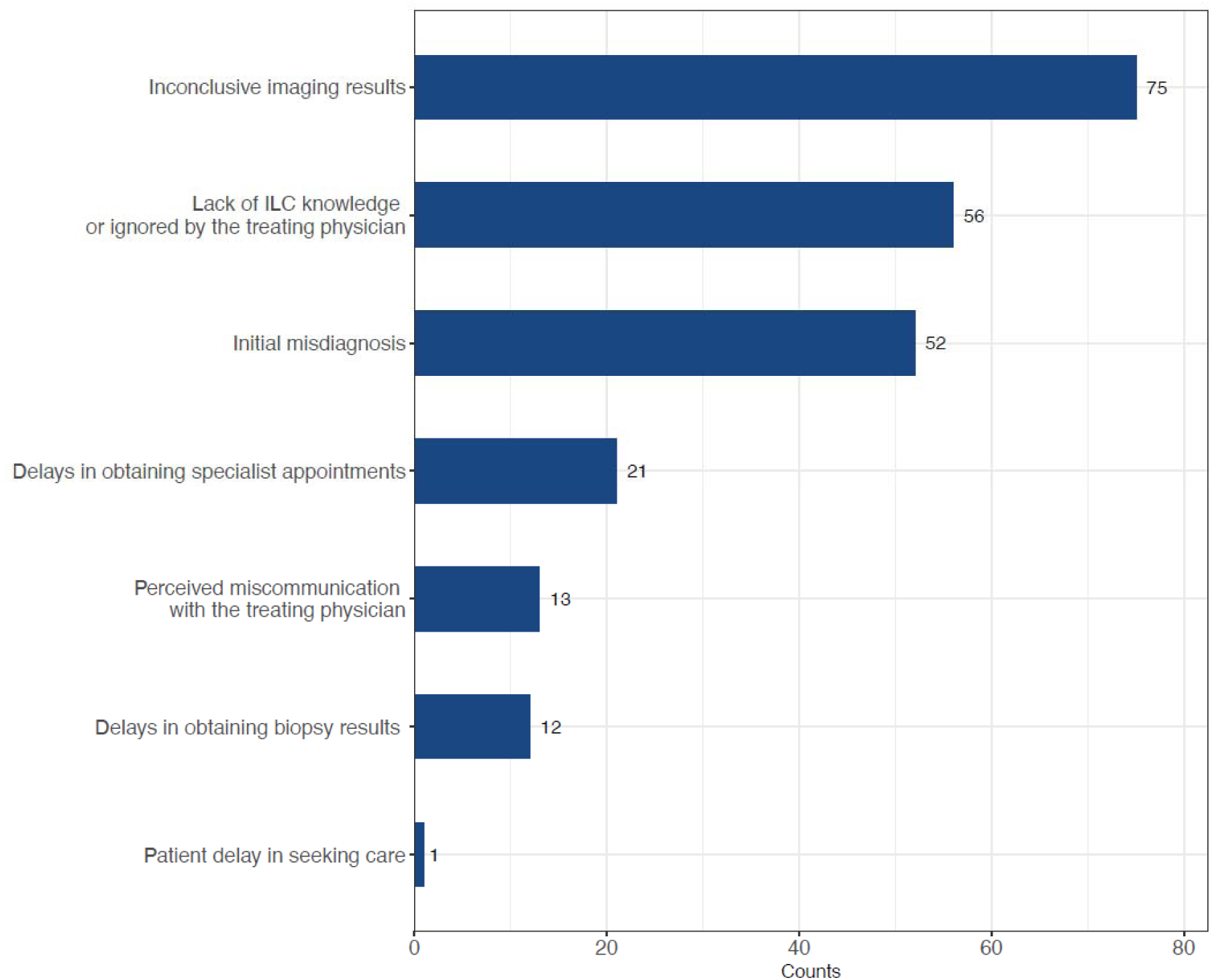
Patient-reported reasons for the delay in accurate diagnosis. Inconclusive imaging results, lack of provider knowledge, and initial misdiagnosis were the most frequently patient-reported contributors for the delay in receiving an accurate diagnosis.

Of the 321 patients with mILC, 138 (42.9%) reported experiencing symptoms prior to diagnosis, the most common being back pain (16.5%), fatigue/malaise (14.9%), GI symptoms (11.8%), bloating (8.4%), and weight loss (8.1%) (Figure 6).

**Figure 6.**
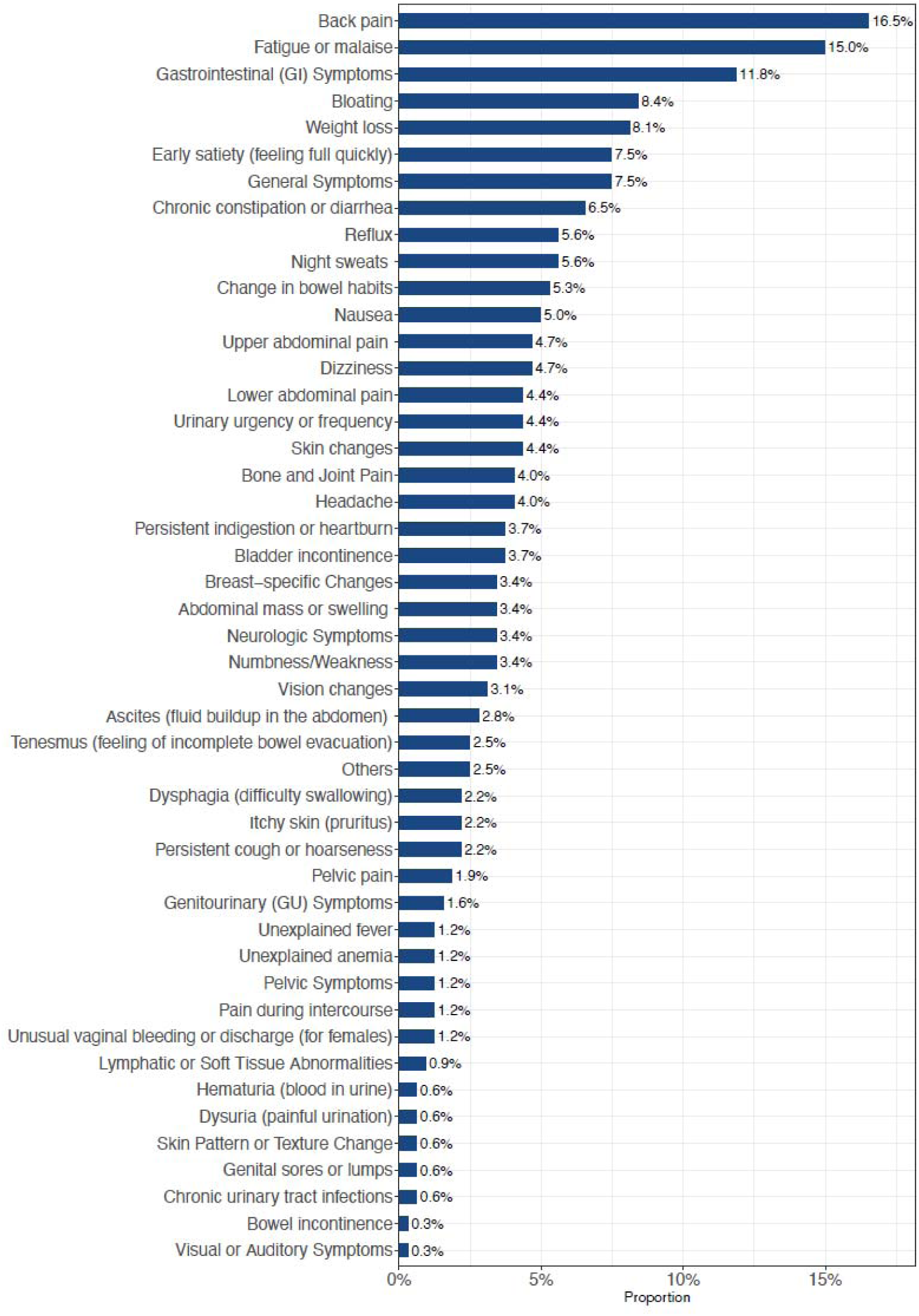
Patient-reported symptoms prior to diagnosis. 138 (42.9%) of the 321 patients with mILC reported experiencing symptoms prior to diagnosis. Proportionally, back pain (16.5%), fatigue or malaise (15%), and GI symptoms (11.8%) were the most common symptoms reported.

### Imaging Performance and Surveillance Limitations

Although 40% of patients reported having received a mammogram at the time of their initial misdiagnosis, ILC was detected in only 20.5% of these cases (24/116), and mammography detected only 5 (25%) of the 20 de novo mILC cases (Figure 7).

**Figure 7.**
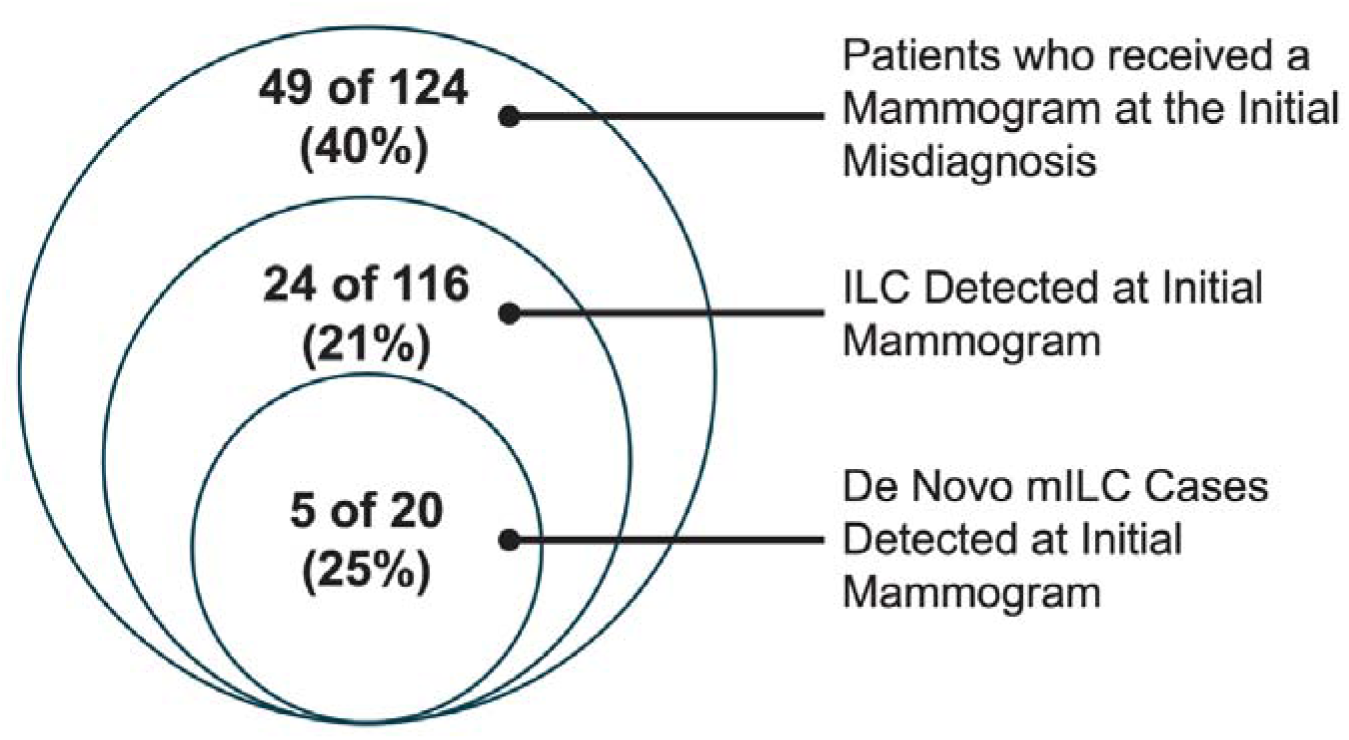
Mammographic Findings for Patients Initially Misdiagnosed. 40% (49 of 124) of patients received a mammogram at the time of their initial misdiagnosis; ILC was detected in only 20.5% (24 of 116) of cases, and mammography detected only 5 of the 20 (25%) de novo mILC cases.

Patients reported receiving additional diagnostic testing within 1-3 months of their initial mammogram, including biopsy, ultrasound, and MRI. Close to half (47.6%) of patients were in active BC surveillance at the time of their mILC diagnosis; however, no statistical difference was seen in time to diagnosis compared to those not under surveillance.

## Discussion

To our knowledge, this is the first study to systematically investigate the patient-reported prevalence of misdiagnosis in mILC. Our findings reveal that more than one in four patients with mILC experienced at least one misdiagnosis, and nearly half waited over a year for an accurate diagnosis. These results point to four key contributors—atypical metastatic sites, nonspecific symptoms, imaging limitations, and gaps in provider knowledge—all of which represent actionable targets for improving outcomes.

### De Novo mILC Presentation

Of the 525 patient surveys completed, 450 patients were diagnosed with ILC, and 321 were diagnosed with mILC, with 33.3% (n=107) reported diagnosis with *de novo* mILC at initial presentation, which is higher than de novo mILC reported in the literature. In a recent retrospective cohort analysis from Ontario, approximately 4–6% of patients with newly diagnosed lobular breast carcinoma were found to have distant metastases^28^. This is consistent with findings from other studies that reported 4-6.5% de novo mILC rates in newly diagnosed ILC patients^29,30,31^. We believe that although we defined de novo mILC at initial diagnosis in our patient questionnaire, some patients may have confused de novo mILC with recurrent mILC due to the lengthy time lapse between their initial ILC diagnosis and their recurrent mILC diagnosis. Additionally, the higher proportion of patients reporting de novo mILC in our study may reflect a survey response bias, with patients more likely to complete the survey in the hope of improving future diagnostic evaluation and workup. Raghavendra et al reported that the median progression-free survival (PFS) was shorter in patients with de novo mILC (0.57 years) than in those with recurrent mILC metastases (0.67 years), however, the overall survival (OS) of the de novo mILC cohort was longer (3.87 years) than the recurrent mILC cohort (2.6 years)^32^. Given the reported impact of de novo mILC to survival rates, early detection of ILC before metastatic spread is of critical importance in managing this breast cancer subtype.

### Atypical Metastatic Sites and Patterns of Misdiagnosis

In our study, 26.2% (n=84) of the mILC patients reported misdiagnosis, with 31% (n=26) reporting ≥2 misdiagnoses before receiving an accurate one. Diagnostic work-up for most patients included CT, biopsy, MRI, labs, and ultrasound; however, 44.5% of patients waited ≥1 year for an accurate diagnosis. A subset of those misdiagnosed (32 of 84) described their treatment with musculoskeletal, pain, or GI therapies, and 6 patients indicated they received unnecessary cancer treatments (Figure 4). These findings align with our literature review, which revealed no prior studies systematically assessing misdiagnosis prevalence in mILC, highlighting the importance of our patient-reported survey as an approach to address this gap.

ILC has a distinct metastatic pattern with a more frequent metastatic spread to the gastrointestinal and reproductive tract compared to NST^33^. Of patients with mILC in our study, primary GI cancer misdiagnosis was reported at 3.9%, and gynecological cancer misdiagnosis was reported at 2.6% (Figure 2). Consistent with the histopathologic growth pattern of primary ILC in the breast, mILC typically infiltrates target organs in a diffuse, single-file pattern rather than forming a discrete mass lesion, further complicating radiologic detection and tissue sampling^2,13,28^. In alignment with the clinical literature, patients with mILC in our study reported misdiagnosis due to diagnostic delays at 7.8% (Figure2). Additional text comment analysis identified inaccurate/inconclusive imaging results or lack of imaging result follow-up as the primary reasons for diagnostic delay. Recognition of these unique biologic and clinical features is essential to improving diagnostic accuracy, staging strategies, and therapeutic management for patients with ILC.

For example, mILC spread to the gastrointestinal tract, reported in the range of 0.8%-18% in case series, poses a challenge in clinical diagnosis and patient management^34^. Although rare, primary stomach cancer can also metastasize to the breast^35,36^, therefore, it is critical to avoid misdiagnosis by distinguishing gastric metastasis from breast cancer to select the optimal initial treatment for metastatic spread of breast cancer^37^.

In a retrospective analysis of 78 cases of gastric metastasis from breast cancer, Xu L. et al found that the majority of the patients initially presented with stage II breast cancer (35.9%), abdominal pain was the most common symptom of gastric metastases (75.6%), and 51/78 patients (65.4%) had a history of ILC^38^. Additionally, another retrospective analysis of 481 patients with ILC found 74 patients had metastases to atypical sites at a median follow-up of 46 months, with five of the 74 patients (6.76%) diagnosed with histologically confirmed gastric metastases of ILC^39^. In another study, of the patients with gastric metastases, the prevalent primary sites were breast cancer (27.9%), followed by lung cancer (23.8%), esophageal cancer (19.1%), renal cell carcinoma (7.6%), and malignant melanoma (7.0%)^40^. These examples all highlight the importance of using tools like immunohistochemistry and imaging analysis, i.e., endoscopy for GI symptoms, to arrive at an accurate diagnosis, followed by effective treatment.

### Non-Specific Symptoms

In our study, 43% (138 of 321) of patients with mILC reported experiencing symptoms prior to diagnosis, with GI symptoms and bloating being frequent complaints (Figure 6). Symptoms like persistent abdominal pain, bloating, or changes in bowel habits should prompt investigation for mILC, especially in patients with a history of early-stage ILC. Based on patient comments in our survey, metastatic breast cancer was not always considered on the initial differential diagnosis for these complaints.

At the primary care level, nonspecific symptoms associated with mILC may not be recognized as breast cancer–related for several reasons. Due to the unique metastatic spread of ILC, resulting symptoms like abdominal bloating, altered bowel habits, early satiety, pelvic discomfort, urinary frequency, or vague back pain, which are highly prevalent in the general population, may be attributed to benign or primary organ-specific conditions (e.g., irritable bowel syndrome, diverticular disease, ovarian pathology, musculoskeletal pain). Consequently, symptoms arising from gastrointestinal or gynecologic involvement may prompt referral to gastroenterology or gynecology rather than immediate oncologic evaluation.

ILC is characterized by a prolonged natural history and a propensity for late recurrence, often more than 5–10 years after initial treatment. When patients present after a long disease-free interval, both primary care clinicians and patients may cognitively “de-link” new symptoms from the prior breast cancer diagnosis. In this scenario, the longer the remission period, the lower the perceived pre-test probability of recurrence may be, even though ILC retains meaningful long-term relapse risk. Survivorship transitions—where oncology follow-up becomes less frequent and patient care is primarily managed in primary care—can further attenuate oncologic vigilance.

### Imaging Challenges in ILC Detection

#### Mammography Limitations

Due to the unique ILC biology and growth, mammography has been found to demonstrate lower sensitivity compared to NST (79 % vs 57%) for detection, with up to a 30% false negative rate^41^. In a study by Krecke and Gisvold, 54% of mammograms that showed no evidence of ILC malignancy were subsequently found to be suggestive of a tumor^42^.

Patients in our survey indicated inconclusive imaging results as the top reason for delay in receiving an accurate diagnosis (Figure 5). Although 40% of patients in our survey indicated having received a mammogram at the time of their initial misdiagnosis, ILC was detected in only 20.5% (24/116) of these cases, and mammography detected only 5 of 20 (25%) de novo mILC cases (Figure 7). Although close to half (47.6%) of patients were in active breast cancer surveillance after curative intent therapy at the time of their mILC diagnosis, no statistical difference was seen in time to mILC diagnosis compared to those not under surveillance. Patient respondents commented that having dense breast tissue was a factor in obtaining accurate mammographic detection, which is consistent with reported clinical literature findings. For example, a study by Berg. et al found that in patients with dense breasts, mammographic sensitivity dropped from 81% to 60% for IDC, and from 34% to 11% for ILC^43^.

#### PET Imaging Considerations

In addition to ILC’s single-file linear cell pattern, ILC has a low proliferation rate and lower expression of glucose transporter 1 (GLUT1) protein^44^. Because of its low metabolic rate, metastatic ILC often shows poor glucose uptake, making the tumors difficult to distinguish from normal background tissue, and less sensitive to fluorine-18 (18F)-fluorodeoxyglucose-positron emission tomography (FDG-PET) imaging^45^. This is consistent with patients in our survey indicating in the free-text comments they received false-negative FDG-PET imaging scans.

18F-Fluoroestradiol-PET (FES-PET), a relatively new modality, uses an estradiol analogue radiotracer that targets functional estrogen receptors (ER). This makes it useful when assessing ILC, as the majority of ILC cases are ER-positive^46^. FES-PET as a promising but imperfect alternative to FDG-PET, however, it is not effective in evaluating the uterus, liver, gallbladder, small bowel, and urinary system, which are common sites of mILC ^47^. The NCCN guidelines note FES-PET can be useful for specific scenarios in treating ER-positive mILC and/or recurrent ILC, including 1) helping to select appropriate patients for endocrine therapy 2) evaluating lesions that are equivocal or suspicious on other imaging tests, and 3) evaluating lesions that are difficult to biopsy, or when biopsy is non-diagnostic^48^. PET/CT imaging with novel radiolabeled molecules to accurately assess hormone receptors and HER2 expression offers a promising non-invasive approach to guide patient management without the need for repeat biopsy^49^.

#### Ultrasound as an Adjunctive Tool

Ultrasound (US) is not used as a breast cancer screening tool; however, diagnostic breast US has proved useful as an adjunctive modality in evaluating suspicious mammographic findings or physical exam findings such as a palpable lump, palpable thickening, palpable ipsilateral axillary lymph nodes, and nipple inversion. US has been reported to have sensitivity for the detection of ILC between 68% and 98% ^50,51^.

#### Advanced Imaging Modalities

More advanced imaging techniques, such as MRI and contrast-enhanced spectral mammography (CESM), are more effective in detecting ILC and assessing tumor extent, especially in dense breast tissue; however, they are not routinely used in initial screenings^52,53^. Additionally, the NCCN guidelines state that MRI may be useful when ILC is poorly or inadequately defined on mammography, ultrasound, or physical examination or to stage disease^54,55^. The NICE guidelines include a specific recommendation to offer an MRI scan before surgery to assess tumor size, where breast-conserving surgery is planned^56^.

#### Limitations of Conventional Imaging for Micro Metastases

Additionally, micro metastases often cannot be detected with conventional imaging. Our group recently published results of our rapid autopsy program that revealed numerous discrepancies in which peritoneal organs often had metastases despite ‘consistently’ normal CTs. For example, a case of ILC where the patient was deemed to have repeatedly had a ‘normal liver’ on CT up until death, only at autopsy to reveal multiple metastases that were not visible clinically^57^. Similar findings have been reported by Desmedt et al in the UPTIDER rapid autopsy study^58^.

#### Provider Knowledge and Patient Empowerment

Patients surveyed indicated a lack of ILC knowledge by providers as one of the top two reasons for delays in accurate diagnosis (Figure 5). In the analysis of the free-text responses of our survey, patients described reporting their symptoms to primary care physicians but noted that these symptoms were not initially recognized as potentially indicative of metastatic spread.

These patient-reported findings align with a recent international survey of breast cancer physicians (N=416), laboratory-based researchers (N=376), and patients (N=1126) from 66 countries which revealed gaps in ILC knowledge and communication^59^. Most physicians reported being very/extremely (41%) to moderately (42%) confident in describing the differences between ILC and NST. However, most patients (52%) thought that their healthcare providers did not sufficiently explain the unique features of ILC. These differences in respondent replies suggest an opportunity for enhanced communication and education that can influence diagnosis and treatment choices for ILC patients.

The combination of nonspecific symptoms, atypical metastatic distribution, diffuse growth pattern, and extended disease-free intervals creates a diagnostic environment in which mILC may not be immediately considered. Increased awareness of ILC’s unique recurrence profile—particularly its potential for late, extranodal, and gastrointestinal presentations—may help reduce diagnostic delay and improve time to appropriate imaging, biopsy, and treatment initiation.

ILC’s unique biology makes symptom recognition more complex, however, informed patients can reduce the risk of prolonged diagnostic delay. Patients can take steps to better recognize and contextualize nonspecific symptoms that may warrant evaluation. Specifically, patients can review education materials from providers like the Lobular Breast Cancer Alliance, which provides ILC-specific educational content that addresses atypical metastatic patterns, the

American Society of Clinical Oncology (Cancer.Net patient materials), National Comprehensive Cancer Network (NCCN Guidelines for Patients®), and the American Cancer Society. These materials will help educate patients to recognize which ‘non-specific’ symptoms that are persistent, progressive, or unexplained deserve attention by their primary care physician, especially given ILC’s higher propensity for late recurrence.

Central to this patient survey was the participation of advocacy partners and non-profit lobular breast cancer groups who helped ensure the survey addressed issues that were clinically meaningful to patients—specifically misdiagnosis, delayed recognition of metastatic disease, and the impact of nonspecific symptoms. Their participation strengthened the real-world relevance of the findings in addressing the diagnostic challenges associated with the atypical and often nonspecific presentations of ILC and mILC.

#### Study Limitations

There are limitations to this patient survey. We relied on patient memory to document their diagnostic journey and therefore, recall bias may impact patient responses. Symptoms, misdiagnosis, and sites of metastatic spread were not analyzed based on histologic subtypes, as this information was not collected. As with all patient surveys, survey respondents are highly motivated to complete the survey in the hopes of impacting future directions. In this case, patients provided anecdotal information (free-text comments) on their diagnostic journey and challenges, which may yield interesting case reports for future insights.

#### Future Directions and Clinical Implications

Our patient survey results underscore the urgent need to improve diagnostic strategies for mILC. The free-text fields in this survey also indicated that patients want improved early detection through enhanced imaging modalities and diagnostic protocols combined with improved provider knowledge of ILC. This patient viewpoint aligns with clinical literature documenting the challenges of early detection of ILC and the need for enhanced education on specific ILC diagnosis and treatment guidelines.

Our ultimate objective is to develop an artificial intelligence (AI)-based model that utilizes health records for ILC-related symptoms and nudges physicians to consider metastatic disease in patients with a history of ILC. We believe an AI model with machine learning algorithms that can identify and transform clinical data points and patterns with speed, accuracy, and reliability into actionable patient-specific diagnostic insights will reduce misdiagnosis and delay in mILC. Additionally, such an AI-based tool can be expanded to aid patient prognosis and treatment selection.

## Conclusion

Addressing delays and diagnostic errors in mILC, along with improving provider education and knowledge of the disease, is critical to optimizing treatment strategies and improving patient outcomes. Prior cancer diagnoses may be embedded within medical records and are frequently overlooked, particularly in patients who have remained disease-free or in remission for prolonged periods, thereby contributing to diagnostic delays. An AI-assisted tool capable of systematically identifying such historical diagnoses would represent a valuable addition to the screening and surveillance armamentarium for metastatic invasive lobular carcinoma (mILC).

## Supporting information

Supplemental Table 1, ILC misdiagnosis copy of patient survey

Supplemental Table 2, ILC misdiagnosis patient age and gender

Supplemental Table 3, ILC misdiagnosis Supplemental patient survey data

Supplemental Table 4, ILC misdiagnosis patient survey free text examples

## Acknowledgements

We thank all respondents to the survey who made this work possible. This work was performed as part of the University of Pittsburgh Medical School Longitudinal Research Program (MC). We thank the many patient advocates representing the different ILC advocacy groups who contributed to editing the original survey questions. This work was made possible in part by the UPMC Hillman Cancer Center with support from NIH grant award P30CA047904. This survey and its outcome are dedicated to Julia Levine, who participated in its design and execution, but sadly passed away from mILC during this study.

## Author Information

### Deceased

Julia Levine

## Author Contributions

M.C. developed the study concept, survey methodology, and survey questions; set up the on-line survey, interpreted the data, wrote the main manuscript; provided direction for development of the figures; annotated all figures (1-7); and provided project administration.

Critical input on survey development, including questions from S.O. and A.L. Overall project supervision provided by S.O. and A.L.

Final refinement of survey questions provided by J.F., C.D., R.J., M.B., and our patient advocates T.C., C.D, S.F., J.L., L.P., N.R., K.S., and C.T.

Collection and analysis of survey data by M.C and R.C. Images for figures 1-7 were generated by R.C.

Supervision of data analysis by G.T.

Primary contributors providing editorial review of the manuscript were S.O., A.L., J.F., C.D. All authors read and approved the final manuscript except patient advocate J.L., who passed away before finishing the final draft of the manuscript.

## Ethics declarations Competing interests

The other authors indicated no potential conflicts of interest.

## Data availability

The study does not contain any sequencing or structural data. The original survey document has been uploaded into Supplementary Data Files. Additional raw data are available upon reasonable request from the corresponding author.

