## Supplemental Table 1, ILC misdiagnosis copy of patient survey for "Quantitative and qualitative patient-reported analysis of misdiagnosis and/or late diagnosis of metastatic lobular cancer"

### ILC Misdiagnosis Survey

Start of Block: ILC SURVEY Start of Block: Eligibility

Dear Participant,
We invite you to participate in a voluntary survey focused on Invasive Lobular Breast Cancer (ILC), developed by patients, physicians, and researchers. The survey aims to gather insights into patients' diagnostic journeys, including experiences of misdiagnosis and delays. The survey includes 40 questions and is approved by the University of Pittsburgh Institutional Review Board. Results will be summarized in a manuscript, guiding future collaborative research efforts. Please complete the survey by May 31, 2025.
For context-related questions, contact Drs. Adrian Lee at and Steffi Oesterreich at. Breast cancer patients and advocates with questions can reach out to Tracy Cushing at. For technical issues with the survey, please contact Morgan Cody at.
You received this survey because of your participation in prior ILC Symposia, involvement with patient advocacy groups, or through social media and support groups related to ILC. We are not using any commercially obtained email lists, but you may receive multiple invitations or see it shared across various platforms. Please only complete this survey once, and do not share it with individuals who are not ILC patients. This ensures we gather responses from the correct patient population and avoid duplicate responses.
**If you have had multiple misdiagnoses**, you can complete this survey more than once—**once for each *separate* misdiagnosis**. On the final screen of the survey, after the optional contact information section, there will be an option that allows you to retake the survey and add additional misdiagnoses. You will also find a text box at the end where you can share more details about your experiences or conditions, especially if something was missed or doesn't apply to you.
The survey is anonymous, but you may choose to provide your name, affiliation, and contact information if you wish to be recognized in the publication of survey results. Your responses will remain anonymous, even if you choose to share your contact details.
Thank you for your time and participation.

End of Block: ILC SURVEY Start of Block: Eligibility

Start of Block: Personal and Medical History

X→

Q1 What is your age?

- a. Under 40 (1)
- b. 40-49 (2)
- c. 50-59 (3)
- d. 60-69 (4)
- e. 70-79 (5)
- f. 80 or older (6)

Q2 What is your gender?

- a. Female (1)
- b. Male (2)
- c. Other (3) ________________________________________________

Q3 Have you been diagnosed with Invasive Lobular Carcinoma (ILC)?

- No (1)
- Yes (2)

Q4 If yes, when were you first diagnosed with ILC? (Year)

- ○ a. 2020 or later (1)
- ○ b. 2015-2019 (2)
- ○ c. 2010-2014 (3)
- ○ d. 2005-2009 (4)
- ○ e. 2000-2004 (5)
- ○ f. 1995-1999 (6)
- ○ g. Before 1995, please specify. (7)

________________________________________________

---

Q5 Have you been diagnosed with metastatic ILC?

- ○ Yes (1)
- ○ No (2)

---

Q6 Did you have any other medical conditions at the time of your metastatic ILC diagnosis?

- ○ No (1)
- ○ Yes, please specify (2) ________________________________________________

---

Q7 Were you diagnosed with de novo metastatic ILC (meaning metastatic cancer at the time of your initial diagnosis)?

- ○ No (1)
- ○ Yes (2)

**End of Block: Personal and Medical History**

---

**Start of Block: Misdiagnosis and Delays**

Q8 How much time passed between your initial diagnosis of metastatic ILC and the confirmation of metastatic disease?

- ○ a. Less than 1 month (1)
- ○ b. 1-3 months (2)
- ○ c. 3-6 months (3)
- ○ d. 6-12 months (4)
- ○ e. If more than 1 year, please specify (5) ________________________________________________

---

Q9 Was your metastatic ILC ever misdiagnosed as another condition throughout the history of your disease?

- ○ a. Yes (1)
- ○ b. No (2)

---

Q10 How many misdiagnoses have you received? This refers to instances where, during your workup, you were diagnosed with a condition other than metastatic ILC.

- ○ 1 (1)
- ○ 2 or more (2)

---

Q11 If you have experienced **multiple misdiagnoses**, please list them in the order you received them. Then, complete this survey separately for each misdiagnosis. Start with the first misdiagnosis, complete the survey, then return to take it again for the second misdiagnosis, and so on. At the end of the survey, after the optional contact information section, you will have the option to retake the survey to submit additional misdiagnoses.

________________________________________________________________

________________________________________________________________

________________________________________________________________

________________________________________________________________

________________________________________________________________

---

Q12 What was the misdiagnosis?

- a. Primary gastrointestinal cancer (1)
- b. Gynecological cancer (2)
- c. Benign gynecological condition (3)
- d. Endometriosis (4)
- e. Benign breast condition (5)
- f. Another type of breast cancer (6)
- g. Gastritis (7)
- h. Peptic Ulcer Disease (8)
- i. Gastroesophageal Reflux Disease (GERD) (9)
- j. Irritable Bowel Syndrome (IBS) (10)
- k. Diverticulitis (11)
- l. Bone related condition (12)
- m. Bone fracture (13)
- n. Hydronephrosis (14)
- o. Anemia (16)
- p. Other (please specify) (15) ________________

Q13 How long did it take to receive the correct metastatic ILC diagnosis after the misdiagnosis?

- ○ a. Less than 1 month (1)
- ○ b. 1-3 months (2)
- ○ c. 3-6 months (3)
- ○ d. 6-12 months (4)
- ○ e. More than 1 year (5)

---

Q14 Do you feel there was a delay in diagnosing or identifying the progression of your metastatic ILC?

- ○ a. Yes (1)
- ○ b. No (2)

---

Q15 If yes, what do you believe contributed to the delay? (Select all that apply)

- ☐ a. Initial misdiagnosis (1)
- ☐ b. Inconclusive imaging results (2)
- ☐ c. Delays in obtaining specialist appointments (3)
- ☐ d. Delays in obtaining biopsy results (4)
- ☐ e. Perceived miscommunication with the treating physician (5)
- ☐ f. Lack of ILC knowledge or ignored by the treating physician (6)
- ☐ g. Other (please specify) (7) ____________________
- ☐ h. Not applicable (no delay) (8)

**End of Block: Misdiagnosis and Delays**

---

**Start of Block: Symptoms and Presentation of mILC**

Q16 Did you experience any symptoms before your metastatic ILC diagnosis or progression?

- ○ a. Yes (1)
- ○ b. No (2)

Q17 If yes, what symptoms did you experience? (Select all that apply)

- ☐ Gastrointestinal (GI) Symptoms: (3)
- ☐ a. Upper abdominal pain (4)
- ☐ b. Lower abdominal pain (5)
- ☐ c. Bloating (6)
- ☐ d. Nausea (7)
- ☐ e. Reflux (8)
- ☐ f. Change in bowel habits (9)
- ☐ g. Dysphagia (difficulty swallowing) (10)
- ☐ h. Early satiety (feeling full quickly) (11)
- ☐ i. Heme-positive stools (bloody stool) (12)
- ☐ j. Hematemesis (vomiting blood) (13)
- ☐ k. Jaundice (yellowing of the skin or eyes) (14)
- ☐ l. Ascites (fluid buildup in the abdomen) (15)
- ☐ m. Chronic constipation or diarrhea (16)
- ☐ n. Tenesmus (feeling of incomplete bowel evacuation) (17)
- ☐ o. Rectal bleeding (18)
- ☐ p. Persistent indigestion or heartburn (19)

- [ ] q. Other (please specify) (20)

---

- [ ] Genitourinary (GU) Symptoms: (21)
- [ ] r. Bladder incontinence (22)
- [ ] s. Bowel incontinence (23)
- [ ] t. Hematuria (blood in urine) (24)
- [ ] u. Urinary urgency or frequency (25)
- [ ] v. Dysuria (painful urination) (26)
- [ ] w. Other (please specify) (27)

---

- [ ] General Symptoms: (28)
- [ ] x. Weight loss (29)
- [ ] y. Unexplained fever (30)
- [ ] z. Fatigue or malaise (31)
- [ ] aa. Back pain (32)
- [ ] bb. Night sweats (33)
- [ ] cc. Itchy skin (pruritus) (34)
- [ ] dd. Skin changes (35)
- [ ] ee. Abdominal mass or swelling (36)

- ☐ ff. Unexplained anemia (37)
- ☐ gg. Persistent cough or hoarseness (38)
- ☐ hh. Tachycardia (rapid heartbeat, palpitations) (39)
- ☐ ii. Other (please specify) (40) ________________

- ☐ Pelvic Symptoms: (41)
- ☐ jj. Pelvic pain (42)
- ☐ kk. Pain during intercourse (43)
- ☐ ll. Unusual vaginal bleeding or discharge (for females) (44)
- ☐ mm. Genital sores or lumps (45)
- ☐ nn. Chronic urinary tract infections (46)
- ☐ oo. Other (please specify) (47) ________________

- ☐ Neurologic Symptoms: (48)
- ☐ a. Numbness/Weakness (49)
- ☐ b. Vision changes (50)
- ☐ c. Headache (51)
- ☐ d. Dizziness (52)
- ☐ e. Other (please specify) (53) ________________

Q18 Were your symptoms diagnosed as a primary gastrointestinal/genitourinary/hematological/neurological condition?

- a. Yes (1)
- b. No, please specify other condition (2) ________________________________________________

Q19 If yes, what was the gastrointestinal initial misdiagnosis?

- a. Gastric cancer (1)
- b. Colon cancer (2)
- c. Gastric polyps (3)
- d. Gastritis (4)
- e. Esophagitis (5)
- f. Esophageal polyps (6)
- g. Peptic Ulcer Disease (7)
- h. Inflammatory Bowel Disease (IBD) (8)
- i. Irritable Bowel Syndrome (IBS) (9)
- j. Intestinal polyps (10)
- k. Gastroesophageal Reflux Disease (GERD) (11)
- l. Gallstones (12)
- m. Hepatitis (13)
- n. Pancreatitis (14)
- o. Other, please specify (15) ________________

Q20 What was the initial genitourinary misdiagnosis?

- a. UTI (1)
- b. Bladder spasm (2)
- c. Kidney stone (3)
- d. Ureteral stricture (4)
- e. Uterine fibroids (5)
- f. Other, please specify (6) ________________

Q21 What was the initial neurologic misdiagnosis?

- a. Eye/vision/retinal problem (1)
- b. Multiple sclerosis (2)
- c. Migraine headache (3)
- d. Other, please specify (4) ________________

Q22 What was the initial hematological misdiagnosis?

- a. Multiple myeloma (1)
- b. Bone marrow suppression (2)
- c. Anemia (3)
- d. Other, please specify (4) ________________

Q23 Were you treated for the misdiagnosed condition?

- a. Yes, please specify treatment (1) ________________________________________________
- b. No (2)

Q24 Were you referred to a specialist after the initial misdiagnosis?

- a. Yes (1)
- b. No (2)

Q25 If yes, what type of specialist were you referred to?

- a. Gastroenterologist (1)
- b. Oncologist (2)
- c. Urologist (3)
- d. Gynecologist (4)
- e. Surgeon (5)
- f. Neurologist (6)
- g. Ophthalmologist (7)
- h. Other, please specify (8) ________________________________________________

Q26 What diagnostic tests were performed to identify the cause of your symptoms? (Select all that apply)

- [ ] a. Upper endoscopy (Esophagogastroduodenoscopy)  (1)
- [ ] b. Colonoscopy  (2)
- [ ] c. Biopsy  (3)
- [ ] d. CT scan  (4)
- [ ] e. Ultrasound  (5)
- [ ] f. MRI  (6)
- [ ] g. Blood work  (8)
- [ ] h. Other, please specify  (7) ________________________________________________

Q27 Were any of these tests positive for cancer?

- ○ a. Yes  (1)
- ○ b. No  (2)

Q28 If yes, which test(s) indicated cancer? (Select all that apply)

- ☐ a. Upper endoscopy (Esophagogastroduodenoscopy) (1)
- ☐ b. Colonoscopy (2)
- ☐ c. Biopsy (3)
- ☐ d. CT scan (4)
- ☐ e. PET scan (5)
- ☐ f. MRI (6)
- ☐ g. Other, please specify (7) ________________________________________________

Q29 If yes, which type of cancer were you told that you had?

- a. Esophageal adenocarcinoma (1)
- b. Esophageal squamous cell carcinoma (2)
- c. Gastric adenocarcinoma (3)
- d. Colorectal adenocarcinoma (4)
- e. Hepatocellular carcinoma (5)
- f. Pancreatic adenocarcinoma (6)
- g. Gallbladder adenocarcinoma (7)
- h. Bladder cancer (8)
- i. Ovarian cancer (9)
- j. Endometrial cancer (10)
- k. Cervical cancer (11)
- l. Fallopian Tube Cancer (12)
- m. Other, please specify (13) ________________________________________________

---

Q30 Were you treated for the misdiagnosed cancer?

- a. Yes, please specify (1) ________________________________________________
- b. No (2)

End of Block: Symptoms and Presentation of mILC

Start of Block: Diagnostic Process

Q31 At the time of your misdiagnosis, did you have a mammogram as part of your advanced or metastatic ILC diagnostic process? (if you had ILC in the past, this question refers to the recent misdiagnosis period, not the initial diagnosis)

- a. Yes (1)
- b. No (2)

Q32 If yes, how long after your symptoms did you have your first mammogram?

- a. Less than 1 month (1)
- b. 1-3 months (2)
- c. 3-6 months (3)
- d. 6-12 months (4)
- e. More than 1 year (5)

Q33 Was the mammogram able to detect the ILC?

- a. Yes (1)
- b. No (2)
- c. Not applicable (no mammogram) (3)

Q34 Did you undergo additional testing? (Select all that apply)

- ▢ a. Ultrasound (1)
- ▢ b. MRI (2)
- ▢ c. 3D mammogram (3)
- ▢ d. Biopsy (4)
- ▢ e. Other, please specify (5) ________________
- ▢ f. None (6)

Q35 How long after your initial mammogram did you undergo these additional tests?

- ○ a. Less than 1 month (1)
- ○ b. 1-3 months (2)
- ○ c. 3-6 months (3)
- ○ d. 6-12 months (4)
- ○ e. More than 1 year (5)
- ○ f. Not applicable (no additional tests) (6)

Q36 Were you undergoing active surveillance for breast cancer at the time of your metastatic ILC diagnosis?

○ No (1)

○ Yes (2)

---

Q37 If yes, what type of surveillance? (Select all that apply)

☐ a. CT scan (1)

☐ b. PET scan (2)

☐ c. Mammogram (3)

☐ d. Ultrasound (4)

☐ e. MRI (5)

☐ f. Lab tests (tumor markers such as CA-125, CEA) (6)

☐ g. Liquid biopsy (circulating tumor cells or DNA, such as Signatera) (7)

☐ h. Other, please specify (8)
________________________________________________

---

Q38 Do you believe your misdiagnosis affected your overall health and treatment plan? If yes, please describe.

________________________________________________________________

________________________________________________________________

________________________________________________________________

________________________________________________________________

Q39 During your treatment for metastatic ILC, did you ever experience symptoms or findings that were related to cancer progression but were not initially recognized by your clinician or imaging studies?

- a. Yes (1)
- b. No (2)

Q40 Do you believe your unrecognized progression affected your quality of life and/or treatment plan? If yes, please describe.

Q41 Is there anything else you would like to share regarding your diagnosis of metastatic ILC that is not covered in this Survey?

End of Block: Diagnostic Process

Start of Block: Closing

Q42 Would you be interested in being contacted for follow-up questions or to share your story in more detail?  If you prefer not to be contacted, please mark an "X" next to "I do not wish to be contacted and would like to remain anonymous."

○ a. Name (First, Last)  (1) ________________________________________________

○ b. Phone number  (2) ________________________________________________

○ c. Email address  (3) ________________________________________________

○ d. I do not wish to be contacted and would like to remain anonymous  (4) ________________________________________________

---

Q43 Thank you for your time in completing the survey. We would like to recognize you for your time and effort **by including your name in the acknowledgments section** of any manuscript describing the results of the survey. If you agree to having your name listed, please insert your name, affiliation, and email below.   If you prefer to remain anonymous, please put an "X" next to the option: "I am not interested and would like to remain anonymous."

○ a. Name (First, Last)  (1) ________________________________________________

○ b. Affiliation  (2) ________________________________________________

○ c. Email address  (3) ________________________________________________

○ d. I am not interested and would like to remain anonymous  (4) ________________________________________________

**End of Block: Closing**
