## Supplemental Table 2, ILC misdiagnosis patient age and gender for "Quantitative and qualitative patient-reported analysis of misdiagnosis and/or late diagnosis of metastatic lobular cancer"

| | Completed Survey Respondent Count | | | |
|---|---|---|---|---|
| | ILC | | mILC | |
| **Age Range Years** | Female | Male | Female | Male |
| <40 yrs | 8 | 1 | 6 | 1 |
| 40-49 yrs | 55 | 0 | 30 | 0 |
| 50-59 yrs | 139 | 1 | 83 | 2 |
| 60-69 yrs | 162 | 0 | 129 | 0 |
| 70-79 yrs | 72 | 0 | 59 | 0 |
| >=80 yrs | 12 | 0 | 11 | 0 |
| **Totals** | **448** | **2** | **318** | **3** |
| | **450** | | **321** | |

| Totals | | | |
|---|---|---|---|
| **ILC** | **% by Age** | **mILC** | **% by Age** |
| 9 | 2% | 7 | 2% |
| 55 | 12% | 30 | 9% |
| 140 | 31% | 85 | 26% |
| 162 | 36% | 129 | 40% |
| 72 | 16% | 59 | 18% |
| 12 | 3% | 11 | 3% |
| **450** | **100%** | **321** | **100%** |

Please note that not all mILC patients identifited themselves as ILC patients. 10 out of 321 mILC patients answered ‘NO’ to the ILC question.

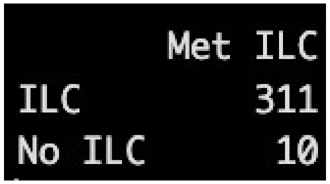

| | Met ILC |
|---|---|
| ILC | 311 |
| No ILC | 10 |
