## Supplemental Table 3, ILC misdiagnosis Supplemental patient survey data for "Quantitative and qualitative patient-reported analysis of misdiagnosis and/or late diagnosis of metastatic lobular cancer"

| Q1 | Q2 | Q3 | Q4 | Q5 | Q6 | Q7 |
|---|---|---|---|---|---|---|
| What is your age? | What is your gender? - Selected Choice | Have you been diagnosed with Invasive Lobular Carcinoma (ILC)? | If yes, when were you first diagnosed with ILC? (Year) - Selected Choice | Have you been diagnosed with metastatic ILC? | Did you have any other medical conditions at the time of your metastatic ILC diagnosis? - Selected Choice | Were you diagnosed with de novo metastatic ILC (meaning metastatic cancer at the time of your initial diagnosis)? |
| 4 | 1 | 2 | 2 | 1 | 2 | 2 |
| 3 | 1 | 2 | 1 | 1 | 2 | 1 |
| 4 | 1 | 2 | 3 | 1 | 1 | 1 |
| 3 | 1 | 2 | 1 | 2 | | |
| 3 | 1 | 2 | 1 | 1 | 2 | 2 |
| 4 | 1 | 2 | 2 | 1 | 2 | 2 |
| 3 | 1 | 2 | 2 | 1 | 1 | 1 |

| Q1 | Q2 | Q3 | Q4 | Q5 | Q6 | Q7 |
|---|---|---|---|---|---|---|
| What is your age? | What is your gender? - Selected Choice | Have you been diagnosed with Invasive Lobular Carcinoma (ILC)? | If yes, when were you first diagnosed with ILC? (Year) - Selected Choice | Have you been diagnosed with metastatic ILC? | Did you have any other medical conditions at the time of your metastatic ILC diagnosis? - Selected Choice | Were you diagnosed with de novo metastatic ILC (meaning metastatic cancer at the time of your initial diagnosis)? |
| 4 | 1 | 2 | 1 | 1 | 2 | 1 |
| 3 | 1 | 2 | 2 | 1 | 1 | 1 |
| 5 | 1 | 2 | 5 | 1 | 1 | 1 |
| 1 | 2 | 2 | 4 | 1 | 1 | 1 |
| 3 | 1 | 2 | 1 | 1 | 1 | 1 |
| 2 | 1 | 2 | 1 | 1 | 2 | 2 |
| 4 | 1 | 2 | 2 | 2 | | |
| 3 | 1 | | 3 | 2 | | |
| 4 | 1 | 2 | 5 | 1 | 1 | 1 |
| 4 | 1 | 2 | 2 | 1 | 2 | 1 |
| 4 | 1 | 2 | 1 | 1 | 1 | 2 |
| 5 | 1 | 2 | 5 | 1 | 1 | 1 |
| 5 | 1 | 1 | | 1 | 2 | 2 |

| Q1 | Q2 | Q3 | Q4 | Q5 | Q6 | Q7 |
| --- | --- | --- | --- | --- | --- | --- |
| What is your age? | What is your gender? - Selected Choice | Have you been diagnosed with Invasive Lobular Carcinoma (ILC)? | If yes, when were you first diagnosed with ILC? (Year) - Selected Choice | Have you been diagnosed with metastatic ILC? | Did you have any other medical conditions at the time of your metastatic ILC diagnosis? - Selected Choice | Were you diagnosed with de novo metastatic ILC (meaning metastatic cancer at the time of your initial diagnosis)? |
| 5 | 1 | 2 | 1 | 1 | 1 | 1 |
| 3 | 1 | 2 | 1 | 2 | | |
| 5 | 1 | 2 | 1 | 1 | 2 | 1 |
| 5 | 1 | 2 | 1 | 1 | 1 | 2 |
| 3 | 1 | 2 | 1 | 1 | 2 | 2 |
| 3 | 1 | 2 | 1 | 1 | 1 | 1 |
| 3 | 1 | 2 | 3 | 1 | 2 | 1 |
| 3 | 1 | 2 | 2 | 1 | 2 | 1 |

| Q1 | Q2 | Q3 | Q4 | Q5 | Q6 | Q7 |
|---|---|---|---|---|---|---|
| What is your age? | What is your gender? - Selected Choice | Have you been diagnosed with Invasive Lobular Carcinoma (ILC)? | If yes, when were you first diagnosed with ILC? (Year) - Selected Choice | Have you been diagnosed with metastatic ILC? | Did you have any other medical conditions at the time of your metastatic ILC diagnosis? - Selected Choice | Were you diagnosed with de novo metastatic ILC (meaning metastatic cancer at the time of your initial diagnosis)? |
| 2 | 1 | 1 | 1 | 1 | 1 | 2 |
| 4 | 1 | 2 | 4 | 1 | 1 | 1 |
| 2 | 1 | 2 | 2 | 1 | 1 | 1 |
| 4 | 1 | 2 | 2 | 1 | 2 | 2 |
| 4 | 1 | 2 | 2 | 1 | 1 | 1 |
| 2 | 1 | 2 | 1 | 2 | | |
| 4 | 1 | 2 | 3 | 1 | 2 | 1 |
| 4 | 1 | 2 | 1 | 1 | 1 | 1 |
| 4 | 1 | 2 | 2 | 1 | 1 | 1 |

| Q1 | Q2 | Q3 | Q4 | Q5 | Q6 | Q7 |
|---|---|---|---|---|---|---|
| What is your age? | What is your gender? - Selected Choice | Have you been diagnosed with Invasive Lobular Carcinoma (ILC)? | If yes, when were you first diagnosed with ILC? (Year) - Selected Choice | Have you been diagnosed with metastatic ILC? | Did you have any other medical conditions at the time of your metastatic ILC diagnosis? - Selected Choice | Were you diagnosed with de novo metastatic ILC (meaning metastatic cancer at the time of your initial diagnosis)? |
| 4 | 1 | 2 | 2 | 1 | 1 | 1 |
| 4 | 1 | 2 | 3 | 1 | 1 | 1 |
| 4 | 1 | 2 | 1 | 1 | 2 | 2 |
| 3 | 1 | 2 | 1 | 1 | 1 | 2 |
| 3 | 1 | 2 | 1 | 1 | 2 | 2 |
| 2 | 1 | 2 | 1 | 2 | | |
| 2 | 1 | 2 | 1 | 1 | 1 | 1 |
| 3 | 1 | 2 | 1 | 1 | 1 | 1 |
| 3 | 1 | 2 | 1 | 2 | | |

| Q1 | Q2 | Q3 | Q4 | Q5 | Q6 | Q7 |
|---|---|---|---|---|---|---|
| **What is your age?** | **What is your gender? - Selected Choice** | **Have you been diagnosed with Invasive Lobular Carcinoma (ILC)?** | **If yes, when were you first diagnosed with ILC? (Year) - Selected Choice** | **Have you been diagnosed with metastatic ILC?** | **Did you have any other medical conditions at the time of your metastatic ILC diagnosis? - Selected Choice** | **Were you diagnosed with de novo metastatic ILC (meaning metastatic cancer at the time of your initial diagnosis)?** |
| 4 | 1 | 2 | 1 | 2 | | |
| 3 | 1 | 2 | 1 | 2 | | |
| 3 | 1 | 2 | 1 | 2 | | |
| 4 | 1 | 2 | 3 | 1 | 1 | 1 |
| 3 | 1 | 2 | 1 | 2 | | |
| 3 | 1 | 2 | 1 | 1 | 1 | 2 |
| 3 | 1 | 2 | 1 | 2 | | |
| 4 | 1 | 2 | 1 | 1 | 1 | 1 |
| 4 | 1 | 2 | 1 | 2 | | |
| 4 | 1 | 2 | 3 | 2 | | |
| 4 | 1 | 2 | 1 | 1 | 2 | 2 |

| Q1 | Q2 | Q3 | Q4 | Q5 | Q6 | Q7 |
|---|---|---|---|---|---|---|
| **What is your age?** | **What is your gender? - Selected Choice** | **Have you been diagnosed with Invasive Lobular Carcinoma (ILC)?** | **If yes, when were you first diagnosed with ILC? (Year) - Selected Choice** | **Have you been diagnosed with metastatic ILC?** | **Did you have any other medical conditions at the time of your metastatic ILC diagnosis? - Selected Choice** | **Were you diagnosed with de novo metastatic ILC (meaning metastatic cancer at the time of your initial diagnosis)?** |
| 1 | 1 | 2 | 1 | 1 | 1 | 1 |
| 3 | 1 | 2 | 2 | 2 | | |
| 4 | 1 | 2 | 1 | 2 | | |
| 5 | 1 | 2 | 1 | 2 | | |
| 4 | 1 | 2 | 3 | 1 | 1 | 1 |
| 4 | 1 | 2 | 1 | 2 | | |
| 3 | 1 | 2 | 2 | 1 | 1 | 2 |
| 5 | 1 | 2 | 1 | 1 | 1 | 1 |
| 3 | 1 | 2 | 1 | 1 | 1 | 2 |
| 2 | 1 | 2 | 1 | 1 | 1 | 2 |
| 2 | 1 | 2 | 1 | 1 | 1 | 2 |
| 3 | 1 | 2 | 2 | 1 | 2 | 1 |
| 4 | 1 | 2 | 2 | 1 | 2 | 1 |
| 2 | 1 | 2 | 2 | 1 | 1 | 1 |
| 4 | 1 | 2 | 2 | 1 | 1 | 1 |
| 5 | 1 | 2 | 1 | 1 | 1 | 1 |
| 3 | 1 | 2 | 1 | 1 | 2 | 1 |
| 3 | 1 | 2 | 1 | 1 | 1 | 1 |

| Q1 | Q2 | Q3 | Q4 | Q5 | Q6 | Q7 |
|---|---|---|---|---|---|---|
| What is your age? | What is your gender? - Selected Choice | Have you been diagnosed with Invasive Lobular Carcinoma (ILC)? | If yes, when were you first diagnosed with ILC? (Year) - Selected Choice | Have you been diagnosed with metastatic ILC? | Did you have any other medical conditions at the time of your metastatic ILC diagnosis? - Selected Choice | Were you diagnosed with de novo metastatic ILC (meaning metastatic cancer at the time of your initial diagnosis)? |
| 5 | 1 | 2 | 5 | 1 | 2 | 1 |
| 5 | 1 | 2 | 1 | 1 | 2 | 1 |
| 4 | 1 | 2 | 1 | 1 | 2 | 1 |
| 4 | 1 | 2 | 3 | 1 | 1 | 1 |
| 4 | 1 | 2 | 2 | 1 | 1 | 1 |
| 2 | 1 | 2 | 1 | 1 | 1 | 2 |
| 6 | 1 | 2 | 1 | 1 | 2 | 2 |
| 4 | 1 | 2 | 2 | 1 | 2 | 1 |
| 4 | 1 | 2 | 1 | 1 | | |
| 4 | 1 | 2 | 2 | 1 | 1 | 2 |
| 4 | 1 | 2 | 2 | 1 | 1 | 1 |
| 5 | 1 | 2 | 3 | 1 | 1 | 1 |
| 5 | 1 | 2 | 1 | 1 | 1 | 1 |
| 6 | 1 | 2 | 4 | 1 | 1 | 1 |
| | 2 | | | | | |
| 4 | 1 | 2 | 4 | 1 | 2 | 1 |
| 5 | 1 | 2 | 1 | 1 | 2 | 2 |
| 2 | 1 | 2 | 1 | 1 | 1 | 2 |
| 3 | 1 | 2 | 2 | 1 | 1 | 1 |
| 2 | 1 | 2 | 1 | 1 | 1 | 1 |
| 1 | 1 | 2 | 1 | 1 | 1 | 2 |
| 3 | 1 | 2 | 1 | 1 | 1 | 1 |
| 4 | 1 | 2 | 2 | 1 | 1 | 1 |
| 3 | 1 | 2 | 1 | 2 | | |
| 1 | 1 | 2 | 1 | 2 | | |

| | | | | | | |
|---|---|---|---|---|---|---|
| **Q1** | **Q2** | **Q3** | **Q4** | **Q5** | **Q6** | **Q7** |
| **What is your age?** | **What is your gender? - Selected Choice** | **Have you been diagnosed with Invasive Lobular Carcinoma (ILC)?** | **If yes, when were you first diagnosed with ILC? (Year) - Selected Choice** | **Have you been diagnosed with metastatic ILC?** | **Did you have any other medical conditions at the time of your metastatic ILC diagnosis? - Selected Choice** | **Were you diagnosed with de novo metastatic ILC (meaning metastatic cancer at the time of your initial diagnosis)?** |
| 4 | 1 | 2 | 2 | 1 | 2 | 2 |
| 3 | 1 | 1 | | 2 | | |
| 3 | 1 | 2 | 2 | 2 | | |
| 5 | 1 | 2 | 3 | 1 | 2 | 1 |
| 3 | 1 | 2 | 2 | 1 | 1 | 1 |
| 4 | 1 | 2 | 1 | 1 | 1 | 1 |
| 3 | 1 | 2 | 1 | 1 | 1 | 2 |
| 2 | 1 | 2 | 1 | 1 | 2 | 1 |
| 2 | 1 | 2 | 1 | 1 | 1 | 1 |
| 2 | 1 | 2 | 2 | 1 | 1 | 1 |
| 4 | 1 | 2 | 1 | 1 | 2 | 2 |
| 4 | 1 | 2 | 3 | 1 | 1 | 1 |
| 3 | 1 | 2 | 1 | 1 | 1 | 2 |
| 3 | 1 | 2 | 1 | 1 | 1 | 1 |
| 2 | 1 | 2 | 1 | 1 | 1 | 1 |
| 3 | 1 | 2 | 1 | 1 | 1 | 1 |

| Q1 | Q2 | Q3 | Q4 | Q5 | Q6 | Q7 |
|---|---|---|---|---|---|---|
| **What is your age?** | **What is your gender? - Selected Choice** | **Have you been diagnosed with Invasive Lobular Carcinoma (ILC)?** | **If yes, when were you first diagnosed with ILC? (Year) - Selected Choice** | **Have you been diagnosed with metastatic ILC?** | **Did you have any other medical conditions at the time of your metastatic ILC diagnosis? - Selected Choice** | **Were you diagnosed with de novo metastatic ILC (meaning metastatic cancer at the time of your initial diagnosis)?** |
| 1 | 1 | 2 | 1 | 1 | 1 | 1 |
| 3 | 1 | 2 | 1 | 1 | 1 | 1 |
| 4 | 1 | 2 | 3 | 1 | 1 | 2 |
| 2 | 1 | 2 | 1 | 2 | | |
| 3 | 1 | 2 | 1 | 2 | | |
| 4 | 1 | 2 | 3 | 2 | | |
| 3 | 1 | 2 | 2 | 2 | | |
| 3 | 1 | 1 | | 2 | | |
| 3 | 1 | 2 | 1 | 2 | | |
| 4 | 1 | 2 | 1 | 1 | 1 | 2 |
| 4 | 1 | 2 | 3 | 2 | | |
| 5 | 1 | 2 | 1 | 1 | 2 | 1 |
| 4 | 1 | 2 | 2 | 1 | 1 | 1 |
| 4 | 1 | 2 | 2 | 1 | 1 | 1 |

| Q1 | Q2 | Q3 | Q4 | Q5 | Q6 | Q7 |
|---|---|---|---|---|---|---|
| **What is your age?** | **What is your gender? - Selected Choice** | **Have you been diagnosed with Invasive Lobular Carcinoma (ILC)?** | **If yes, when were you first diagnosed with ILC? (Year) - Selected Choice** | **Have you been diagnosed with metastatic ILC?** | **Did you have any other medical conditions at the time of your metastatic ILC diagnosis? - Selected Choice** | **Were you diagnosed with de novo metastatic ILC (meaning metastatic cancer at the time of your initial diagnosis)?** |
| 4 | 1 | 2 | 1 | 1 | 1 | 1 |
| 4 | 1 | 2 | 1 | 1 | 2 | 1 |
| 4 | 1 | 2 | 1 | 1 | 1 | 1 |
| 3 | 1 | 2 | 1 | 1 | 1 | 2 |
| 6 | 1 | 2 | 4 | 1 | 1 | 1 |
| 2 | 1 | 2 | 1 | 2 | | |
| 1 | 1 | 2 | 1 | 1 | 1 | 1 |
| 4 | 1 | 2 | 2 | 1 | 1 | 1 |
| 3 | 1 | 2 | 2 | 2 | | |
| 3 | 1 | 2 | 2 | 2 | | |
| 3 | 1 | 2 | 1 | 1 | 2 | 1 |
| 6 | 1 | 2 | 1 | 2 | | |

| Q1 | Q2 | Q3 | Q4 | Q5 | Q6 | Q7 |
|---|---|---|---|---|---|---|
| What is your age? | What is your gender? - Selected Choice | Have you been diagnosed with Invasive Lobular Carcinoma (ILC)? | If yes, when were you first diagnosed with ILC? (Year) - Selected Choice | Have you been diagnosed with metastatic ILC? | Did you have any other medical conditions at the time of your metastatic ILC diagnosis? - Selected Choice | Were you diagnosed with de novo metastatic ILC (meaning metastatic cancer at the time of your initial diagnosis)? |
| 4 | 1 | 2 | 2 | 1 | 1 | 1 |
| 4 | 1 | 2 | 1 | 2 | | |
| 5 | 1 | 2 | 5 | 1 | 1 | 1 |
| 4 | 1 | 2 | 1 | 1 | 1 | 2 |
| 5 | 1 | 2 | 6 | 1 | 2 | 1 |
| 5 | 1 | 2 | 3 | 2 | | |
| 4 | 1 | 2 | 2 | 1 | 1 | 1 |
| 4 | 1 | 2 | 3 | 1 | 1 | 1 |

| Q1 | Q2 | Q3 | Q4 | Q5 | Q6 | Q7 |
| --- | --- | --- | --- | --- | --- | --- |
| What is your age? | What is your gender? - Selected Choice | Have you been diagnosed with Invasive Lobular Carcinoma (ILC)? | If yes, when were you first diagnosed with ILC? (Year) - Selected Choice | Have you been diagnosed with metastatic ILC? | Did you have any other medical conditions at the time of your metastatic ILC diagnosis? - Selected Choice | Were you diagnosed with de novo metastatic ILC (meaning metastatic cancer at the time of your initial diagnosis)? |
| 4 | 1 | 2 | 1 | 1 | 1 | 1 |
| 3 | 1 | 2 | 1 | 2 | | |
| 5 | 1 | 2 | 3 | 2 | | |
| 4 | 1 | 2 | 1 | 1 | 1 | 1 |
| 5 | 1 | 2 | 1 | 2 | | |
| 3 | 1 | 2 | 1 | 2 | | |
| 2 | 1 | 2 | 1 | 1 | 1 | 2 |
| 4 | 1 | 2 | 1 | 1 | 1 | 1 |
| 4 | 1 | 2 | 2 | 2 | | |
| 4 | 1 | 2 | 1 | 2 | | |
| 3 | 1 | 2 | 3 | 1 | 2 | 1 |

| Q1 | Q2 | Q3 | Q4 | Q5 | Q6 | Q7 |
|---|---|---|---|---|---|---|
| What is your age? | What is your gender? - Selected Choice | Have you been diagnosed with Invasive Lobular Carcinoma (ILC)? | If yes, when were you first diagnosed with ILC? (Year) - Selected Choice | Have you been diagnosed with metastatic ILC? | Did you have any other medical conditions at the time of your metastatic ILC diagnosis? - Selected Choice | Were you diagnosed with de novo metastatic ILC (meaning metastatic cancer at the time of your initial diagnosis)? |
| 5 | 1 | 2 | 5 | 1 | 1 | 2 |
| 5 | 1 | 2 | 1 | 1 | 2 | 1 |
| 3 | 1 | 2 | 2 | 1 | 1 | 2 |
| 4 | 1 | 2 | 3 | 1 | 1 | 1 |
| 5 | 1 | 2 | 2 | 2 | | |
| 5 | 1 | 2 | 2 | 2 | | |
| 4 | 1 | 2 | 2 | 2 | | |
| 4 | 1 | 2 | 3 | 1 | 2 | 1 |
| 3 | 1 | 2 | 2 | 1 | 1 | 2 |
| 3 | 1 | 2 | 1 | 1 | 1 | 1 |

| Q1 | Q2 | Q3 | Q4 | Q5 | Q6 | Q7 |
|---|---|---|---|---|---|---|
| What is your age? | What is your gender? - Selected Choice | Have you been diagnosed with Invasive Lobular Carcinoma (ILC)? | If yes, when were you first diagnosed with ILC? (Year) - Selected Choice | Have you been diagnosed with metastatic ILC? | Did you have any other medical conditions at the time of your metastatic ILC diagnosis? - Selected Choice | Were you diagnosed with de novo metastatic ILC (meaning metastatic cancer at the time of your initial diagnosis)? |
| 5 | 1 | 2 | 4 | 1 | 1 | 1 |
| 4 | 1 | 2 | 1 | 2 | | |
| 4 | 1 | 2 | 1 | 2 | | |
| 3 | 1 | 2 | 1 | 1 | 2 | 2 |
| 5 | 1 | 1 | 1 | 1 | 1 | 2 |
| 5 | 1 | 2 | 3 | 1 | 1 | 1 |
| 3 | 1 | 2 | 2 | 1 | 2 | 1 |

| Q1 | Q2 | Q3 | Q4 | Q5 | Q6 | Q7 |
|---|---|---|---|---|---|---|
| What is your age? | What is your gender? - Selected Choice | Have you been diagnosed with Invasive Lobular Carcinoma (ILC)? | If yes, when were you first diagnosed with ILC? (Year) - Selected Choice | Have you been diagnosed with metastatic ILC? | Did you have any other medical conditions at the time of your metastatic ILC diagnosis? - Selected Choice | Were you diagnosed with de novo metastatic ILC (meaning metastatic cancer at the time of your initial diagnosis)? |
| 3 | 2 | 1 | 1 | 1 | 1 | 1 |
| 2 | 1 | 2 | 1 | 1 | 2 | 2 |
| 5 | 1 | 2 | 1 | 1 | | 1 |
| 5 | 1 | 2 | 1 | 2 | | |
| 2 | 1 | 2 | 1 | 2 | | |
| 4 | 1 | 2 | 5 | 1 | 1 | 1 |
| 5 | 1 | 2 | 1 | 1 | 1 | 2 |
| 3 | 1 | 2 | 1 | 2 | | |
| 3 | 1 | 2 | 1 | 1 | 1 | 2 |
| 4 | 1 | 2 | 1 | 2 | | |
| 5 | 1 | 2 | 1 | 2 | | |
| 4 | 1 | 2 | 2 | 1 | 1 | 1 |
| 4 | 1 | 2 | 2 | 1 | 2 | 1 |
| 3 | 1 | 2 | 2 | 1 | 2 | 1 |
| 4 | 1 | 2 | 1 | 2 | | |

| Q1 | Q2 | Q3 | Q4 | Q5 | Q6 | Q7 |
| --- | --- | --- | --- | --- | --- | --- |
| What is your age? | What is your gender? - Selected Choice | Have you been diagnosed with Invasive Lobular Carcinoma (ILC)? | If yes, when were you first diagnosed with ILC? (Year) - Selected Choice | Have you been diagnosed with metastatic ILC? | Did you have any other medical conditions at the time of your metastatic ILC diagnosis? - Selected Choice | Were you diagnosed with de novo metastatic ILC (meaning metastatic cancer at the time of your initial diagnosis)? |
| 3 | 1 | 2 | 2 | 1 | 2 | 1 |
| 3 | 1 | 2 | 2 | 2 | | |
| 2 | 1 | 1 | | 2 | | |
| 3 | 1 | 2 | 1 | 2 | | |
| 4 | 1 | 2 | 1 | 1 | 2 | 1 |
| 3 | 1 | 2 | 1 | 1 | 2 | 2 |
| 4 | 1 | 2 | 2 | 1 | 2 | 2 |

| Q1 | Q2 | Q3 | Q4 | Q5 | Q6 | Q7 |
|---|---|---|---|---|---|---|
| **What is your age?** | **What is your gender? - Selected Choice** | **Have you been diagnosed with Invasive Lobular Carcinoma (ILC)?** | **If yes, when were you first diagnosed with ILC? (Year) - Selected Choice** | **Have you been diagnosed with metastatic ILC?** | **Did you have any other medical conditions at the time of your metastatic ILC diagnosis? - Selected Choice** | **Were you diagnosed with de novo metastatic ILC (meaning metastatic cancer at the time of your initial diagnosis)?** |
| 2 | 1 | 2 | 1 | 1 | 1 | 1 |
| 4 | 1 | 2 | 1 | 1 | 1 | 1 |
| 3 | 1 | 2 | 1 | 2 | | |
| 4 | 1 | 2 | 2 | 1 | 1 | 2 |
| 4 | 1 | 2 | 4 | 1 | 1 | 1 |
| 2 | 1 | 2 | 1 | 1 | 1 | 2 |
| 4 | 1 | 2 | 1 | 1 | 1 | 2 |
| 4 | 1 | 2 | 1 | 1 | 1 | 2 |
| 2 | 1 | 2 | 1 | 1 | 1 | 2 |

| Q1 | Q2 | Q3 | Q4 | Q5 | Q6 | Q7 |
|---|---|---|---|---|---|---|
| What is your age? | What is your gender? - Selected Choice | Have you been diagnosed with Invasive Lobular Carcinoma (ILC)? | If yes, when were you first diagnosed with ILC? (Year) - Selected Choice | Have you been diagnosed with metastatic ILC? | Did you have any other medical conditions at the time of your metastatic ILC diagnosis? - Selected Choice | Were you diagnosed with de novo metastatic ILC (meaning metastatic cancer at the time of your initial diagnosis)? |
| 4 | 1 | 2 | 2 | 1 | 1 | 1 |
| 5 | 1 | 2 | 2 | 1 | 2 | 2 |
| 4 | 1 | 2 | 1 | 2 | | |
| 4 | 1 | 2 | 1 | 1 | 1 | 2 |
| 3 | 1 | 2 | 1 | 1 | 2 | 1 |

| Q1 | Q2 | Q3 | Q4 | Q5 | Q6 | Q7 |
|---|---|---|---|---|---|---|
| What is your age? | What is your gender? - Selected Choice | Have you been diagnosed with Invasive Lobular Carcinoma (ILC)? | If yes, when were you first diagnosed with ILC? (Year) - Selected Choice | Have you been diagnosed with metastatic ILC? | Did you have any other medical conditions at the time of your metastatic ILC diagnosis? - Selected Choice | Were you diagnosed with de novo metastatic ILC (meaning metastatic cancer at the time of your initial diagnosis)? |
| 4 | 1 | 2 | 1 | 1 | 1 | 2 |
| 5 | 1 | 2 | 2 | 1 | 1 | 2 |
| 4 | 1 | 2 | 2 | 1 | 2 | 1 |
| 5 | 1 | 2 | 2 | 1 | 2 | 1 |
| 4 | 1 | 2 | 1 | 1 | 1 | 2 |
| 4 | 1 | 2 | 1 | 2 | | |
| 5 | 1 | 2 | 2 | 2 | | |
| 5 | 1 | 2 | 4 | 1 | 1 | 2 |
| 3 | 1 | | 3 | 1 | 1 | 1 |
| 4 | 1 | 2 | 3 | 1 | 1 | 1 |
| 5 | 1 | 2 | 4 | 1 | 1 | 1 |
| 5 | 1 | 2 | 2 | 1 | 2 | 1 |

| Q1 | Q2 | Q3 | Q4 | Q5 | Q6 | Q7 |
|---|---|---|---|---|---|---|
| What is your age? | What is your gender? - Selected Choice | Have you been diagnosed with Invasive Lobular Carcinoma (ILC)? | If yes, when were you first diagnosed with ILC? (Year) - Selected Choice | Have you been diagnosed with metastatic ILC? | Did you have any other medical conditions at the time of your metastatic ILC diagnosis? - Selected Choice | Were you diagnosed with de novo metastatic ILC (meaning metastatic cancer at the time of your initial diagnosis)? |
| 4 | 1 | 2 | 1 | 2 | | |
| 5 | 1 | 2 | 2 | 2 | | |
| 4 | 1 | 2 | 1 | 1 | 2 | 2 |
| 4 | 1 | 2 | 2 | 1 | 1 | 1 |
| 5 | 1 | 2 | 1 | 2 | | |
| 5 | 1 | 2 | 2 | 1 | 1 | 2 |
| 4 | 1 | 2 | 6 | 1 | 1 | 1 |
| 4 | 1 | 2 | 1 | 1 | 1 | 1 |
| 4 | 1 | 2 | 1 | 1 | 1 | 1 |

| Q1 | Q2 | Q3 | Q4 | Q5 | Q6 | Q7 |
|---|---|---|---|---|---|---|
| What is your age? | What is your gender? - Selected Choice | Have you been diagnosed with Invasive Lobular Carcinoma (ILC)? | If yes, when were you first diagnosed with ILC? (Year) - Selected Choice | Have you been diagnosed with metastatic ILC? | Did you have any other medical conditions at the time of your metastatic ILC diagnosis? - Selected Choice | Were you diagnosed with de novo metastatic ILC (meaning metastatic cancer at the time of your initial diagnosis)? |
| 2 | 1 | 2 | 1 | 1 | 1 | 1 |
| 3 | 1 | 2 | 2 | 1 | 1 | 1 |
| 3 | 1 | 2 | 1 | 1 | 1 | 1 |

| Q1 | Q2 | Q3 | Q4 | Q5 | Q6 | Q7 |
|---|---|---|---|---|---|---|
| What is your age? | What is your gender? - Selected Choice | Have you been diagnosed with Invasive Lobular Carcinoma (ILC)? | If yes, when were you first diagnosed with ILC? (Year) - Selected Choice | Have you been diagnosed with metastatic ILC? | Did you have any other medical conditions at the time of your metastatic ILC diagnosis? - Selected Choice | Were you diagnosed with de novo metastatic ILC (meaning metastatic cancer at the time of your initial diagnosis)? |
| 4 | 1 | 2 | 1 | 1 | 2 | 1 |
| 6 | 1 | 1 | 4 | 1 | 2 | 1 |
| 3 | 1 | 2 | 1 | 1 | 1 | 1 |
| 4 | 1 | 2 | 2 | 1 | 2 | 1 |
| 4 | 1 | 2 | 3 | 1 | 1 | 1 |
| 5 | 1 | 2 | 1 | 2 | | |
| 6 | 1 | 2 | 1 | 2 | | |
| 4 | 1 | 2 | 1 | 1 | 1 | 2 |
| 4 | 1 | 2 | 1 | 1 | 1 | 2 |
| 4 | 1 | 2 | 1 | 1 | 1 | 2 |

| Q1 What is your age? | Q2 What is your gender? - Selected Choice | Q3 Have you been diagnosed with Invasive Lobular Carcinoma (ILC)? | Q4 If yes, when were you first diagnosed with ILC? (Year) - Selected Choice | Q5 Have you been diagnosed with metastatic ILC? | Q6 Did you have any other medical conditions at the time of your metastatic ILC diagnosis? - Selected Choice | Q7 Were you diagnosed with de novo metastatic ILC (meaning metastatic cancer at the time of your initial diagnosis)? |
|---|---|---|---|---|---|---|
| 3 | 1 | 2 | 1 | 1 | 1 | 2 |
| 6 | 1 | 2 | 1 | 1 | 1 | 2 |
| 4 | 1 | 2 | 3 | 1 | 1 | 1 |
| 4 | 1 | 2 | 1 | 1 | 1 | 1 |
| 4 | 1 | 2 | 2 | 2 | | |
| 2 | 1 | 2 | 1 | 2 | | |
| 1 | 1 | 2 | 1 | 1 | 1 | 2 |
| 4 | 1 | 2 | 2 | 1 | 1 | 2 |
| 5 | 1 | 2 | 5 | 1 | 1 | 2 |
| 3 | 1 | 2 | 2 | | 1 | 1 |
| 3 | 1 | 2 | 1 | 1 | 2 | 1 |
| 2 | 1 | 2 | 1 | 1 | | |
| 4 | 1 | 2 | 1 | 2 | | |

| Q1 | Q2 | Q3 | Q4 | Q5 | Q6 | Q7 |
|---|---|---|---|---|---|---|
| What is your age? | What is your gender? - Selected Choice | Have you been diagnosed with Invasive Lobular Carcinoma (ILC)? | If yes, when were you first diagnosed with ILC? (Year) - Selected Choice | Have you been diagnosed with metastatic ILC? | Did you have any other medical conditions at the time of your metastatic ILC diagnosis? - Selected Choice | Were you diagnosed with de novo metastatic ILC (meaning metastatic cancer at the time of your initial diagnosis)? |
| 4 | 1 | 2 | 1 | 1 | 2 | 2 |
| 4 | 1 | 2 | 3 | 1 | 1 | 2 |
| 3 | 1 | 2 | 1 | 1 | 1 | 2 |
| 5 | 1 | 2 | 5 | 1 | 2 | 1 |
| 4 | 1 | 2 | 5 | 1 | 2 | 1 |
| 2 | 1 | 2 | 2 | 1 | 2 | 2 |
| 4 | 1 | 2 | 1 | 1 | 1 | 1 |
| 3 | 1 | 1 | | 2 | | |

| Q1 | Q2 | Q3 | Q4 | Q5 | Q6 | Q7 |
|---|---|---|---|---|---|---|
| What is your age? | What is your gender? - Selected Choice | Have you been diagnosed with Invasive Lobular Carcinoma (ILC)? | If yes, when were you first diagnosed with ILC? (Year) - Selected Choice | Have you been diagnosed with metastatic ILC? | Did you have any other medical conditions at the time of your metastatic ILC diagnosis? - Selected Choice | Were you diagnosed with de novo metastatic ILC (meaning metastatic cancer at the time of your initial diagnosis)? |
| 3 | 1 | 2 | 1 | 1 | 1 | 2 |
| 3 | 1 | 2 | 2 | 1 | 1 | 1 |
| 4 | 1 | 2 | 2 | 1 | 1 | 1 |
| 5 | 1 | 1 | | 2 | | |
| 5 | 1 | 2 | 4 | 1 | 2 | 1 |
| 4 | 1 | 2 | 1 | 1 | 2 | 2 |
| 3 | 1 | 2 | 1 | 1 | 1 | 2 |
| 5 | 1 | 2 | 2 | 1 | 1 | 2 |

| Q1 | Q2 | Q3 | Q4 | Q5 | Q6 | Q7 |
|---|---|---|---|---|---|---|
| What is your age? | What is your gender? - Selected Choice | Have you been diagnosed with Invasive Lobular Carcinoma (ILC)? | If yes, when were you first diagnosed with ILC? (Year) - Selected Choice | Have you been diagnosed with metastatic ILC? | Did you have any other medical conditions at the time of your metastatic ILC diagnosis? - Selected Choice | Were you diagnosed with de novo metastatic ILC (meaning metastatic cancer at the time of your initial diagnosis)? |
| 5 | 1 | 2 | 2 | 1 | 1 | 1 |
| 4 | 1 | 2 | 1 | 1 | 1 | 1 |
| 5 | 1 | 2 | 1 | 1 | 2 | 2 |
| 4 | 1 | 2 | 1 | 1 | 1 | 1 |
| 6 | 1 | 2 | | 1 | 1 | 1 |
| 3 | 1 | 2 | 1 | 1 | | 1 |
| 6 | 1 | 2 | 1 | 1 | 1 | 2 |
| 4 | 1 | 2 | 1 | 1 | 1 | 1 |
| 4 | 1 | 2 | 2 | 1 | 1 | 1 |
| 4 | 1 | 2 | 2 | 2 | | |
| 6 | 1 | 2 | 1 | 1 | 1 | 2 |
| 1 | 1 | 2 | 1 | 1 | 2 | 2 |
| 3 | 1 | 2 | 1 | 1 | 1 | 1 |
| 3 | 1 | 2 | 2 | 1 | 1 | 1 |
| 6 | 1 | 2 | 1 | 1 | 2 | 1 |
| 5 | 1 | 2 | 3 | 1 | 1 | 2 |
| 5 | 1 | 1 | 1 | 1 | 1 | 2 |
| 5 | 1 | 2 | 2 | 1 | 1 | 1 |
| 3 | 1 | 2 | 1 | 1 | 1 | 2 |

| Q1 | Q2 | Q3 | Q4 | Q5 | Q6 | Q7 |
|---|---|---|---|---|---|---|
| What is your age? | What is your gender? - Selected Choice | Have you been diagnosed with Invasive Lobular Carcinoma (ILC)? | If yes, when were you first diagnosed with ILC? (Year) - Selected Choice | Have you been diagnosed with metastatic ILC? | Did you have any other medical conditions at the time of your metastatic ILC diagnosis? - Selected Choice | Were you diagnosed with de novo metastatic ILC (meaning metastatic cancer at the time of your initial diagnosis)? |
| 4 | 1 | 2 | 4 | 1 | 1 | 1 |
| 5 | 1 | 2 | 2 | 1 | 1 | 1 |
| 5 | 1 | 2 | 1 | 1 | 1 | 1 |
| 3 | 1 | 2 | 3 | 1 | 1 | 1 |
| 5 | 1 | 2 | 1 | 1 | 2 | 2 |
| 5 | 1 | 2 | 2 | 1 | 1 | 1 |
| 5 | 1 | 2 | 6 | 1 | | |
| 5 | 1 | 2 | 4 | 1 | 1 | 1 |
| 4 | 1 | 2 | 4 | 1 | 1 | 1 |
| 4 | 1 | 2 | 1 | 1 | 2 | 2 |
| 4 | 1 | 2 | 1 | 1 | 1 | 1 |
| 5 | 1 | 2 | 1 | 1 | 2 | 1 |
| 4 | 1 | 2 | 3 | 1 | 1 | 1 |
| 4 | 1 | 2 | 1 | 1 | 2 | 2 |

| Q1 | Q2 | Q3 | Q4 | Q5 | Q6 | Q7 |
|---|---|---|---|---|---|---|
| What is your age? | What is your gender? - Selected Choice | Have you been diagnosed with Invasive Lobular Carcinoma (ILC)? | If yes, when were you first diagnosed with ILC? (Year) - Selected Choice | Have you been diagnosed with metastatic ILC? | Did you have any other medical conditions at the time of your metastatic ILC diagnosis? - Selected Choice | Were you diagnosed with de novo metastatic ILC (meaning metastatic cancer at the time of your initial diagnosis)? |
| 3 | 1 | 1 | 1 | 1 | 2 | 2 |
| 4 | 1 | 2 | 3 | 1 | 1 | 1 |
| 2 | 1 | 2 | 1 | 1 | 1 | 1 |
| 4 | 1 | 2 | 1 | 1 | 1 | 1 |
| 3 | 1 | 2 | 3 | 1 | 1 | 1 |
| 2 | 1 | 2 | 1 | 2 | | |
| 3 | 1 | 2 | 1 | 1 | 1 | 1 |
| 3 | 1 | 2 | 1 | 1 | 1 | 2 |
| 3 | 1 | 2 | 1 | 1 | 1 | 2 |
| 5 | 1 | 2 | 1 | 1 | 1 | 2 |
| 3 | 1 | 2 | 1 | 1 | 2 | 2 |
| 3 | 1 | 2 | 6 | 1 | 1 | 2 |
| 4 | 1 | 1 | 1 | 1 | 2 | 2 |
| 4 | 1 | 2 | 3 | 1 | 1 | 1 |
| 5 | 1 | 2 | 5 | 1 | 1 | 1 |
| 3 | 2 | 2 | 1 | 1 | 1 | 1 |
| 4 | 1 | 2 | 1 | 1 | 1 | 1 |
| 3 | 1 | 2 | 3 | 1 | 1 | 1 |

| Q1 | Q2 | Q3 | Q4 | Q5 | Q6 | Q7 |
|---|---|---|---|---|---|---|
| What is your age? | What is your gender? - Selected Choice | Have you been diagnosed with Invasive Lobular Carcinoma (ILC)? | If yes, when were you first diagnosed with ILC? (Year) - Selected Choice | Have you been diagnosed with metastatic ILC? | Did you have any other medical conditions at the time of your metastatic ILC diagnosis? - Selected Choice | Were you diagnosed with de novo metastatic ILC (meaning metastatic cancer at the time of your initial diagnosis)? |
| 4 | 1 | 2 | 2 | 1 | 2 | 2 |
| 3 | 1 | 2 | 4 | 1 | 1 | 1 |
| 3 | 1 | 2 | 2 | 1 | 1 | 1 |
| 4 | 1 | 2 | 4 | 1 | 2 | 1 |
| 3 | 1 | 2 | 1 | 1 | 1 | 1 |
| 5 | 1 | 2 | 1 | 1 | 1 | 1 |
| 5 | 1 | 1 | 2 | 1 | 1 | 2 |
| 3 | 1 | 2 | 3 | 1 | 1 | 1 |
| 4 | 1 | 2 | 1 | 1 | 1 | 1 |
| 4 | 1 | 2 | 1 | 1 | 2 | 1 |
| 4 | 1 | 2 | 2 | 2 | | |
| 6 | 1 | 2 | 2 | 1 | 2 | 1 |
| 4 | 1 | 2 | 1 | 1 | 1 | 1 |
| 4 | 1 | 2 | 3 | 1 | 2 | 1 |
| 5 | 1 | 2 | 1 | 1 | 1 | 1 |

| Q1 | Q2 | Q3 | Q4 | Q5 | Q6 | Q7 |
|---|---|---|---|---|---|---|
| What is your age? | What is your gender? - Selected Choice | Have you been diagnosed with Invasive Lobular Carcinoma (ILC)? | If yes, when were you first diagnosed with ILC? (Year) - Selected Choice | Have you been diagnosed with metastatic ILC? | Did you have any other medical conditions at the time of your metastatic ILC diagnosis? - Selected Choice | Were you diagnosed with de novo metastatic ILC (meaning metastatic cancer at the time of your initial diagnosis)? |
| 4 | 1 | 1 | 2 | 1 | 1 | 2 |
| 5 | 1 | 2 | 6 | 1 | 2 | 2 |
| 4 | 1 | 2 | 5 | 1 | 2 | 1 |
| 2 | 1 | 2 | 1 | 2 | | |
| 4 | 1 | 2 | 1 | 1 | 2 | 2 |
| 4 | 1 | 2 | 1 | 1 | 1 | 2 |
| 4 | 1 | 2 | 2 | 1 | 2 | 1 |
| 4 | 1 | 2 | 3 | 1 | 1 | 1 |
| 4 | 1 | 2 | 2 | 1 | 1 | 2 |
| 2 | 1 | 2 | 1 | 1 | 1 | 1 |
| 3 | 1 | 2 | 1 | 1 | 2 | 1 |
| 3 | 1 | 2 | 1 | 1 | 2 | 1 |
| 3 | 1 | 2 | 2 | 2 | | |
| 2 | 1 | 2 | 1 | 1 | 1 | 1 |

| Q1 | Q2 | Q3 | Q4 | Q5 | Q6 | Q7 |
|---|---|---|---|---|---|---|
| What is your age? | What is your gender? - Selected Choice | Have you been diagnosed with Invasive Lobular Carcinoma (ILC)? | If yes, when were you first diagnosed with ILC? (Year) - Selected Choice | Have you been diagnosed with metastatic ILC? | Did you have any other medical conditions at the time of your metastatic ILC diagnosis? - Selected Choice | Were you diagnosed with de novo metastatic ILC (meaning metastatic cancer at the time of your initial diagnosis)? |
| 4 | 1 | 2 | 2 | 1 | 1 | 2 |
| 5 | 1 | 2 | 1 | 2 | | |
| 5 | 1 | 2 | 1 | 2 | | |
| 3 | 1 | 2 | 1 | 1 | 2 | 2 |
| 3 | 1 | 2 | 2 | 1 | 1 | 1 |
| 5 | 1 | 2 | 1 | 1 | 2 | 2 |
| 3 | 1 | 2 | 2 | 1 | 2 | 1 |
| 4 | 1 | 2 | 3 | 1 | 1 | 1 |
| 5 | 1 | 2 | 1 | 1 | 2 | 1 |
| 5 | 1 | 2 | 2 | 1 | 1 | 1 |
| 4 | 1 | 2 | 1 | 1 | 2 | 1 |
| 4 | 1 | 2 | 2 | 1 | 1 | 1 |
| 5 | 1 | 2 | 1 | 1 | 1 | 1 |
| 4 | 1 | 2 | 1 | 1 | 2 | 1 |
| 3 | 1 | 2 | 1 | 1 | 2 | 1 |

| Q1 | Q2 | Q3 | Q4 | Q5 | Q6 | Q7 |
|---|---|---|---|---|---|---|
| What is your age? | What is your gender? - Selected Choice | Have you been diagnosed with Invasive Lobular Carcinoma (ILC)? | If yes, when were you first diagnosed with ILC? (Year) - Selected Choice | Have you been diagnosed with metastatic ILC? | Did you have any other medical conditions at the time of your metastatic ILC diagnosis? - Selected Choice | Were you diagnosed with de novo metastatic ILC (meaning metastatic cancer at the time of your initial diagnosis)? |
| 2 | 1 | 2 | 1 | 1 | 1 | 1 |
| 2 | 1 | 2 | 1 | 1 | 1 | 2 |
| 2 | 1 | 2 | 2 | 1 | 2 | 1 |
| 4 | 1 | 2 | 2 | 1 | 2 | 1 |
| 5 | 1 | 2 | 4 | 1 | 2 | 1 |
| 4 | 1 | 2 | 2 | 1 | 2 | 1 |
| 2 | 1 | 2 | 2 | 1 | 2 | 1 |
| 4 | 1 | 2 | 1 | 1 | 1 | 1 |
| 3 | 1 | 2 | 3 | 2 | | |
| 3 | 1 | 2 | 1 | 1 | 1 | 2 |
| 5 | 1 | 2 | 4 | 1 | 1 | 1 |
| 3 | 1 | 2 | 1 | 2 | | |
| 4 | 1 | 2 | 1 | 2 | | |
| 3 | | 2 | | 2 | | |
| 3 | 1 | 2 | 1 | 2 | | |

| Q1 | Q2 | Q3 | Q4 | Q5 | Q6 | Q7 |
|---|---|---|---|---|---|---|
| What is your age? | What is your gender? - Selected Choice | Have you been diagnosed with Invasive Lobular Carcinoma (ILC)? | If yes, when were you first diagnosed with ILC? (Year) - Selected Choice | Have you been diagnosed with metastatic ILC? | Did you have any other medical conditions at the time of your metastatic ILC diagnosis? - Selected Choice | Were you diagnosed with de novo metastatic ILC (meaning metastatic cancer at the time of your initial diagnosis)? |
| 3 | 1 | 2 | 1 | 2 | | |
| 3 | 1 | 2 | 1 | 2 | | |
| 3 | 1 | 2 | 1 | 1 | 1 | 1 |
| 3 | 1 | 2 | 1 | 2 | | |
| 3 | 1 | 2 | 1 | 2 | | |
| 2 | 1 | 2 | 1 | 2 | | |
| 2 | 1 | 2 | 1 | 2 | | |
| 4 | 1 | 2 | 1 | 2 | | |
| 3 | 1 | 2 | 1 | 2 | | |
| 1 | 1 | 2 | 1 | 2 | | |
| 3 | 1 | 2 | 1 | 2 | | |
| 2 | 1 | 2 | 1 | 2 | | |
| 5 | 1 | 2 | 1 | 2 | | |
| 3 | 1 | 2 | 1 | 2 | | |
| 3 | 1 | 2 | 1 | 1 | 2 | 2 |
| 3 | 1 | 2 | 1 | 2 | | |
| 3 | 1 | 2 | 1 | 2 | | |
| 2 | 1 | 2 | 1 | 2 | | |
| 4 | 1 | 2 | 1 | 2 | | |
| 3 | 1 | 2 | 2 | 2 | | |
| 3 | 1 | 2 | 1 | 2 | | |
| 3 | 1 | 2 | 1 | 2 | | |
| 3 | 1 | 2 | 1 | 2 | | |
| 4 | 1 | 2 | 1 | 2 | | |
| 3 | 1 | 2 | 1 | 2 | | |
| 3 | 1 | 2 | 1 | 2 | | |
| 3 | 1 | 2 | 2 | 2 | | |

| Q1 | Q2 | Q3 | Q4 | Q5 | Q6 | Q7 |
|---|---|---|---|---|---|---|
| What is your age? | What is your gender? - Selected Choice | Have you been diagnosed with Invasive Lobular Carcinoma (ILC)? | If yes, when were you first diagnosed with ILC? (Year) - Selected Choice | Have you been diagnosed with metastatic ILC? | Did you have any other medical conditions at the time of your metastatic ILC diagnosis? - Selected Choice | Were you diagnosed with de novo metastatic ILC (meaning metastatic cancer at the time of your initial diagnosis)? |
| 2 | 1 | 2 | 1 | 2 | | |
| 3 | 1 | 2 | 1 | 2 | | |
| 4 | 1 | 2 | 4 | 2 | | |
| 4 | 1 | 2 | 4 | 2 | | |
| 5 | 1 | 2 | 7 | 2 | | |
| 4 | 1 | 2 | 3 | 2 | | |
| 2 | 1 | 2 | 1 | 2 | | |
| 2 | 1 | 2 | 1 | 2 | | |
| 2 | 1 | 2 | 1 | 2 | | |
| 4 | 1 | 2 | 1 | 2 | | |
| 3 | 1 | 2 | 1 | 1 | 1 | 2 |
| 3 | 1 | 2 | 1 | 2 | | |
| 5 | 1 | 2 | 1 | 2 | | |
| 2 | 1 | 2 | 1 | 2 | | |
| 4 | 1 | 2 | 1 | 1 | 1 | 2 |
| 3 | 1 | 2 | 1 | 2 | | |
| 2 | 1 | 2 | 1 | 2 | | |
| 2 | 1 | 2 | 1 | 2 | | |
| 3 | 1 | 2 | 1 | 2 | | |
| 3 | 1 | 2 | 1 | 2 | | |

| Q1 | Q2 | Q3 | Q4 | Q5 | Q6 | Q7 |
|---|---|---|---|---|---|---|
| What is your age? | What is your gender? - Selected Choice | Have you been diagnosed with Invasive Lobular Carcinoma (ILC)? | If yes, when were you first diagnosed with ILC? (Year) - Selected Choice | Have you been diagnosed with metastatic ILC? | Did you have any other medical conditions at the time of your metastatic ILC diagnosis? - Selected Choice | Were you diagnosed with de novo metastatic ILC (meaning metastatic cancer at the time of your initial diagnosis)? |
| 4 | 1 | 2 | 3 | 1 | 1 | 2 |
| 3 | 1 | 2 | 2 | 1 | 2 | 1 |
| 4 | 1 | 2 | 1 | 1 | 2 | 1 |
| 6 | 1 | 1 | 1 | 2 | | |
| 2 | 1 | 2 | 1 | 2 | | |
| 2 | 1 | 2 | 1 | 2 | | |
| 2 | 1 | 2 | 1 | 1 | 1 | 2 |

| Q1 | Q2 | Q3 | Q4 | Q5 | Q6 | Q7 |
|---|---|---|---|---|---|---|
| What is your age? | What is your gender? - Selected Choice | Have you been diagnosed with Invasive Lobular Carcinoma (ILC)? | If yes, when were you first diagnosed with ILC? (Year) - Selected Choice | Have you been diagnosed with metastatic ILC? | Did you have any other medical conditions at the time of your metastatic ILC diagnosis? - Selected Choice | Were you diagnosed with de novo metastatic ILC (meaning metastatic cancer at the time of your initial diagnosis)? |
| 4 | 1 | 2 | 1 | 1 | 2 | 1 |
| 6 | 1 | 2 | 5 | 1 | 1 | 1 |
| 2 | 1 | 2 | 1 | 2 | | |
| 2 | 1 | 2 | 1 | 2 | | |
| 3 | 1 | 2 | 3 | 2 | | |
| 3 | 1 | 2 | 1 | 2 | | |
| 4 | 1 | 2 | 1 | 2 | | |
| 2 | 1 | 2 | 1 | 2 | | |
| 3 | 1 | 2 | 1 | 2 | | |
| 2 | 1 | 2 | 1 | 2 | | |
| 3 | 1 | 2 | 1 | 2 | | |
| 4 | 1 | 2 | 1 | 2 | | |

| Q1 | Q2 | Q3 | Q4 | Q5 | Q6 | Q7 |
|---|---|---|---|---|---|---|
| What is your age? | What is your gender? - Selected Choice | Have you been diagnosed with Invasive Lobular Carcinoma (ILC)? | If yes, when were you first diagnosed with ILC? (Year) - Selected Choice | Have you been diagnosed with metastatic ILC? | Did you have any other medical conditions at the time of your metastatic ILC diagnosis? - Selected Choice | Were you diagnosed with de novo metastatic ILC (meaning metastatic cancer at the time of your initial diagnosis)? |
| 4 | 1 | 2 | 1 | 1 | 2 | 1 |
| 3 | 1 | 2 | 1 | 2 | | |
| 2 | 1 | 2 | 1 | 2 | | |
| 3 | 1 | 2 | 1 | 2 | | |
| 3 | 1 | 2 | 1 | 2 | | |
| 3 | 1 | 2 | 1 | 2 | | |
| 3 | 1 | 2 | 1 | 2 | | |
| 3 | 1 | 2 | 2 | 1 | 2 | 1 |
| 3 | 1 | 2 | 3 | 1 | 2 | 1 |
| 3 | 1 | 2 | 2 | 1 | 1 | 1 |
| 4 | 1 | 2 | 1 | 2 | | |
| 3 | 1 | 2 | 1 | 2 | | |
| 3 | 1 | 2 | 2 | 1 | 1 | 1 |

| Q1 | Q2 | Q3 | Q4 | Q5 | Q6 | Q7 |
|---|---|---|---|---|---|---|
| What is your age? | What is your gender? - Selected Choice | Have you been diagnosed with Invasive Lobular Carcinoma (ILC)? | If yes, when were you first diagnosed with ILC? (Year) - Selected Choice | Have you been diagnosed with metastatic ILC? | Did you have any other medical conditions at the time of your metastatic ILC diagnosis? - Selected Choice | Were you diagnosed with de novo metastatic ILC (meaning metastatic cancer at the time of your initial diagnosis)? |
| 4 | 1 | 2 | 2 | | | |
| 3 | 1 | 2 | 3 | 1 | 1 | 1 |
| 3 | 1 | 2 | 3 | 1 | 1 | 1 |

| Q8 | Q9 | Q10 | Q12 | Q12_15_TEXT_chatGPT category |
|---|---|---|---|---|
| How much time passed between your initial diagnosis of metastatic ILC and the confirmation of metastatic disease? - Selected Choice | Was your metastatic ILC ever misdiagnosed as another condition throughout the history of your disease? | How many misdiagnoses have you received? This refers to instances where, during your workup, you were diagnosed with a condition other than metastatic ILC. | What was the misdiagnosis? - Selected Choice | What was the misdiagnosis? - p. Other (please specify) - Text |
| 1 | 1 | | 15 | Hematologic Condition |
| 2 | | | | |
| 2 | 1 | | 15 | Skin-related |
| 2 | 2 | | | |
| 2 | 1 | | 15 | Menopause-related |
| 1 | 1 | | 15 | Gastrointestinal-related |

| Q8 | Q9 | Q10 | Q12 | Q12_15_TEXT_chatGPT category |
|---|---|---|---|---|
| How much time passed between your initial diagnosis of metastatic ILC and the confirmation of metastatic disease? - Selected Choice | Was your metastatic ILC ever misdiagnosed as another condition throughout the history of your disease? | How many misdiagnoses have you received? This refers to instances where, during your workup, you were diagnosed with a condition other than metastatic ILC. | What was the misdiagnosis? - Selected Choice | What was the misdiagnosis? - p. Other (please specify) - Text |
| 1 | 1 | | 15 | Skin-related |
| 1 | 2 | | | |
| 1 | 2 | | | |
| 1 | 1 | 1 | 15 | Others |
| 1 | 2 | | | |
| 1 | 2 | | | |
| 5 | 2 | 1 | 4 | |
| 1 | 2 | | | |
| 1 | 1 | 2 | 13 | |
| 2 | 2 | | | |

| Q8 | Q9 | Q10 | Q12 | Q12_15_TEXT_chatGPT category |
|---|---|---|---|---|
| How much time passed between your initial diagnosis of metastatic ILC and the confirmation of metastatic disease? - Selected Choice | Was your metastatic ILC ever misdiagnosed as another condition throughout the history of your disease? | How many misdiagnoses have you received? This refers to instances where, during your workup, you were diagnosed with a condition other than metastatic ILC. | What was the misdiagnosis? - Selected Choice | What was the misdiagnosis? - p. Other (please specify) - Text |
| 5 | 1 | 1 | 5 | |
| 1 | 2 | 1 | 15 | Lymphatic system |
| 1 | 1 | 2 | 9 | |
| 1 | 2 | | | |
| 5 | 2 | | | |
| 1 | 2 | | | |
| 2 | 2 | | | |

| Q8 | Q9 | Q10 | Q12 | Q12_15_TEXT_chatGPT category |
|---|---|---|---|---|
| How much time passed between your initial diagnosis of metastatic ILC and the confirmation of metastatic disease? - Selected Choice | Was your metastatic ILC ever misdiagnosed as another condition throughout the history of your disease? | How many misdiagnoses have you received? This refers to instances where, during your workup, you were diagnosed with a condition other than metastatic ILC. | What was the misdiagnosis? - Selected Choice | What was the misdiagnosis? - p. Other (please specify) - Text |
| 1 | 2 | | | |
| 5 | 1 | 1 | 15 | Bone related condition |
| 1 | 2 | | | |
| 1 | 1 | 2 | 15 | Menopause-related |
| 5 | 2 | | | |
| 2 | 2 | | | |
| 4 | 1 | 2 | 15 | Skin-related |
| 5 | 2 | | | |

| Q8 | Q9 | Q10 | Q12 | Q12_15_TEXT_chatGPT category |
| --- | --- | --- | --- | --- |
| How much time passed between your initial diagnosis of metastatic ILC and the confirmation of metastatic disease? - Selected Choice | Was your metastatic ILC ever misdiagnosed as another condition throughout the history of your disease? | How many misdiagnoses have you received? This refers to instances where, during your workup, you were diagnosed with a condition other than metastatic ILC. | What was the misdiagnosis? - Selected Choice | What was the misdiagnosis? - p. Other (please specify) - Text |
| 1 | 2 | 1 | 15 | Benign breast condition |
| 2 | 1 | 1 | 1 | |
| 1 | 2 | | | |
| 2 | 2 | | | |
| 1 | 1 | 1 | 15 | Bone related condition |
| 1 | 2 | 1 | 9 | |
| 5 | 1 | 1 | 15 | diagnostic delay |

| Q8 | Q9 | Q10 | Q12 | Q12_15_TEXT_chatGPT category |
|---|---|---|---|---|
| How much time passed between your initial diagnosis of metastatic ILC and the confirmation of metastatic disease? - Selected Choice | Was your metastatic ILC ever misdiagnosed as another condition throughout the history of your disease? | How many misdiagnoses have you received? This refers to instances where, during your workup, you were diagnosed with a condition other than metastatic ILC. | What was the misdiagnosis? - Selected Choice | What was the misdiagnosis? - p. Other (please specify) - Text |
| 5 | 2 | | | |
| 2 | 2 | | | |
| 4 | 2 | | | |
| 1 | 2 | | | |

| Q8 | Q9 | Q10 | Q12 | Q12_15_TEXT_chatGPT category |
|---|---|---|---|---|
| How much time passed between your initial diagnosis of metastatic ILC and the confirmation of metastatic disease? - Selected Choice | Was your metastatic ILC ever misdiagnosed as another condition throughout the history of your disease? | How many misdiagnoses have you received? This refers to instances where, during your workup, you were diagnosed with a condition other than metastatic ILC. | What was the misdiagnosis? - Selected Choice | What was the misdiagnosis? - p. Other (please specify) - Text |
| 4 | 2 | | | |
| 5 | 2 | | 7 | |
| 1 | 1 | 1 | 15 | Menopause-related |
| | 2 | 2 | | |
| 1 | 2 | | | |
| 1 | 2 | | 2 | |
| 1 | 2 | | | |
| 5 | 2 | | | |
| 5 | 2 | | | |
| 1 | 2 | | | |
| 5 | 1 | 2 | | |
| 1 | 2 | 1 | | |
| 1 | 1 | 1 | | |
| 5 | 1 | 2 | 6 | |

| Q8 | Q9 | Q10 | Q12 | Q12_15_TEXT_chatGPT category |
| --- | --- | --- | --- | --- |
| How much time passed between your initial diagnosis of metastatic ILC and the confirmation of metastatic disease? - Selected Choice | Was your metastatic ILC ever misdiagnosed as another condition throughout the history of your disease? | How many misdiagnoses have you received? This refers to instances where, during your workup, you were diagnosed with a condition other than metastatic ILC. | What was the misdiagnosis? - Selected Choice | What was the misdiagnosis? - p. Other (please specify) - Text |
| 4 | 2 | 1 | 15 | |
| 1 | 2 | 1 | 15 | Another type of breast cancer |
| 5 | 2 | 1 | 15 | Bone related condition |
| 5 | 2 | 1 | 15 | Hematologic Condition |
| 1 | 2 | | | |
| 1 | 1 | 1 | 5 | |
| 1 | 2 | | | |
| 5 | 2 | | | |
| 1 | 2 | 1 | | |
| 5 | 2 | | | |
| 1 | 2 | | | |
| 5 | 2 | | | |
| 2 | 2 | | | |
| 1 | 2 | | | |
| 1 | 2 | 1 | | |
| 5 | 2 | | | |
| 1 | 1 | 1 | 15 | |
| 1 | 2 | | | |
| 1 | 1 | 1 | 12 | |
| 5 | 2 | 1 | 15 | Bone related condition |

| Q8 | Q9 | Q10 | Q12 | Q12_15_TEXT_chatGPT category |
|---|---|---|---|---|
| How much time passed between your initial diagnosis of metastatic ILC and the confirmation of metastatic disease? - Selected Choice | Was your metastatic ILC ever misdiagnosed as another condition throughout the history of your disease? | How many misdiagnoses have you received? This refers to instances where, during your workup, you were diagnosed with a condition other than metastatic ILC. | What was the misdiagnosis? - Selected Choice | What was the misdiagnosis? - p. Other (please specify) - Text |
| 5 | 1 | 1 | 15 | Another type of breast cancer |
| 1 | 2 | | | |
| 1 | 2 | | | |
| 2 | 2 | | | |
| 5 | 2 | 1 | 15 | diagnostic delay |
| 5 | 2 | | | |
| 5 | 2 | | | |
| 2 | 2 | | | |
| 5 | 2 | | | |
| 1 | 2 | | | |
| 1 | 2 | | | |
| 4 | 2 | | | |
| 2 | 2 | | | |

| Q8 | Q9 | Q10 | Q12 | Q12_15_TEXT_chatGPT category |
|---|---|---|---|---|
| How much time passed between your initial diagnosis of metastatic ILC and the confirmation of metastatic disease? - Selected Choice | Was your metastatic ILC ever misdiagnosed as another condition throughout the history of your disease? | How many misdiagnoses have you received? This refers to instances where, during your workup, you were diagnosed with a condition other than metastatic ILC. | What was the misdiagnosis? - Selected Choice | What was the misdiagnosis? - p. Other (please specify) - Text |
| 2 | 2 | | | |
| 1 | 2 | | | |
| 1 | 2 | | | |
| 2 | 2 | | | |
| 2 | 2 | | | |
| 5 | 2 | | | |

| Q8 | Q9 | Q10 | Q12 | Q12_15_TEXT_chatGPT category |
|---|---|---|---|---|
| How much time passed between your initial diagnosis of metastatic ILC and the confirmation of metastatic disease? - Selected Choice | Was your metastatic ILC ever misdiagnosed as another condition throughout the history of your disease? | How many misdiagnoses have you received? This refers to instances where, during your workup, you were diagnosed with a condition other than metastatic ILC. | What was the misdiagnosis? - Selected Choice | What was the misdiagnosis? - p. Other (please specify) - Text |
| 5 | 2 | | | |
| 1 | 2 | | | |
| 1 | 2 | 1 | 6 | |
| 1 | 1 | 2 | 5 | |
| 5 | 2 | | | |
| 5 | 2 | | | |
| 4 | 1 | 1 | 15 | Others |
| 1 | 2 | | | |

| Q8 | Q9 | Q10 | Q12 | Q12_15_TEXT_chatGPT category |
|---|---|---|---|---|
| How much time passed between your initial diagnosis of metastatic ILC and the confirmation of metastatic disease? - Selected Choice | Was your metastatic ILC ever misdiagnosed as another condition throughout the history of your disease? | How many misdiagnoses have you received? This refers to instances where, during your workup, you were diagnosed with a condition other than metastatic ILC. | What was the misdiagnosis? - Selected Choice | What was the misdiagnosis? - p. Other (please specify) - Text |
| 2 | 2 | | | |
| 1 | 1 | 1 | 12 | |
| 1 | 1 | 1 | 16 | |
| 5 | 2 | | | |
| 5 | 1 | 1 | 15 | Benign gynecological condition |
| 5 | 1 | 1 | 12 | |

| Q8 | Q9 | Q10 | Q12 | Q12_15_TEXT_chatGPT category |
|---|---|---|---|---|
| How much time passed between your initial diagnosis of metastatic ILC and the confirmation of metastatic disease? - Selected Choice | Was your metastatic ILC ever misdiagnosed as another condition throughout the history of your disease? | How many misdiagnoses have you received? This refers to instances where, during your workup, you were diagnosed with a condition other than metastatic ILC. | What was the misdiagnosis? - Selected Choice | What was the misdiagnosis? - p. Other (please specify) - Text |
| 2 | 1 | 1 | 15 | |
| 1 | 2 | | 5 | |
| 1 | 2 | | | |
| 5 | 1 | 1 | 5 | |
| 1 | 1 | 1 | 15 | Bone related condition |

| Q8 | Q9 | Q10 | Q12 | Q12_15_TEXT_chatGPT category |
|---|---|---|---|---|
| How much time passed between your initial diagnosis of metastatic ILC and the confirmation of metastatic disease? - Selected Choice | Was your metastatic ILC ever misdiagnosed as another condition throughout the history of your disease? | How many misdiagnoses have you received? This refers to instances where, during your workup, you were diagnosed with a condition other than metastatic ILC. | What was the misdiagnosis? - Selected Choice | What was the misdiagnosis? - p. Other (please specify) - Text |
| 1 | 1 | 1 | 5 | |
| 1 | 2 | | | |
| 5 | 1 | 2 | 15 | |
| 2 | 2 | | | |
| 1 | 1 | 2 | 15 | Bone related condition |
| 4 | 2 | | | |
| 4 | 2 | 1 | 14 | |

| Q8 | Q9 | Q10 | Q12 | Q12_15_TEXT_chatGPT category |
|---|---|---|---|---|
| How much time passed between your initial diagnosis of metastatic ILC and the confirmation of metastatic disease? - Selected Choice | Was your metastatic ILC ever misdiagnosed as another condition throughout the history of your disease? | How many misdiagnoses have you received? This refers to instances where, during your workup, you were diagnosed with a condition other than metastatic ILC. | What was the misdiagnosis? - Selected Choice | What was the misdiagnosis? - p. Other (please specify) - Text |
| 1 | 2 | | | |
| 1 | 2 | | | |
| 2 | 2 | | | |
| 5 | 2 | | | |
| 5 | 2 | | 5 | |

| Q8 | Q9 | Q10 | Q12 | Q12_15_TEXT_chatGPT category |
| --- | --- | --- | --- | --- |
| How much time passed between your initial diagnosis of metastatic ILC and the confirmation of metastatic disease? - Selected Choice | Was your metastatic ILC ever misdiagnosed as another condition throughout the history of your disease? | How many misdiagnoses have you received? This refers to instances where, during your workup, you were diagnosed with a condition other than metastatic ILC. | What was the misdiagnosis? - Selected Choice | What was the misdiagnosis? - p. Other (please specify) - Text |
| 1 | 2 | | 15 | diagnostic delay |
| 3 | 1 | 1 | 15 | Benign breast condition |
| 2 | 2 | | | |
| 5 | 2 | 1 | 2 | |
| 1 | 2 | 1 | 6 | |
| 1 | 2 | | | |
| 1 | 2 | 1 | 5 | |
| 2 | 2 | 1 | 6 | |
| 5 | 1 | 2 | 5 | |

| Q8 | Q9 | Q10 | Q12 | Q12_15_TEXT_chatGPT category |
|---|---|---|---|---|
| How much time passed between your initial diagnosis of metastatic ILC and the confirmation of metastatic disease? - Selected Choice | Was your metastatic ILC ever misdiagnosed as another condition throughout the history of your disease? | How many misdiagnoses have you received? This refers to instances where, during your workup, you were diagnosed with a condition other than metastatic ILC. | What was the misdiagnosis? - Selected Choice | What was the misdiagnosis? - p. Other (please specify) - Text |
| 1 | 1 | 2 | 15 | Bone related condition |
| 4 | 1 | 2 | 5 | |
| 1 | 1 | 1 | 15 | Others |
| 2 | 2 | | 15 | Others |

| Q8 | Q9 | Q10 | Q12 | Q12_15_TEXT_chatGPT category |
|---|---|---|---|---|
| How much time passed between your initial diagnosis of metastatic ILC and the confirmation of metastatic disease? - Selected Choice | Was your metastatic ILC ever misdiagnosed as another condition throughout the history of your disease? | How many misdiagnoses have you received? This refers to instances where, during your workup, you were diagnosed with a condition other than metastatic ILC. | What was the misdiagnosis? - Selected Choice | What was the misdiagnosis? - p. Other (please specify) - Text |
| 1 | 1 | 1 | 5 | |
| 3 | 2 | 1 | 12 | |
| 2 | 1 | 1 | 6 | |
| 5 | 2 | | 2 | |
| 2 | 2 | 1 | 15 | |
| 2 | 2 | | | |
| 1 | 1 | 1 | 10 | |
| 1 | 1 | 1 | 15 | Benign breast condition |

| Q8 | Q9 | Q10 | Q12 | Q12_15_TEXT_chatGPT category |
|---|---|---|---|---|
| How much time passed between your initial diagnosis of metastatic ILC and the confirmation of metastatic disease? - Selected Choice | Was your metastatic ILC ever misdiagnosed as another condition throughout the history of your disease? | How many misdiagnoses have you received? This refers to instances where, during your workup, you were diagnosed with a condition other than metastatic ILC. | What was the misdiagnosis? - Selected Choice | What was the misdiagnosis? - p. Other (please specify) - Text |
| 1 | 2 | | | |
| 1 | 2 | | 15 | diagnostic delay |
| 1 | 2 | | | |
| 1 | 1 | 1 | 15 | diagnostic delay |

| Q8 | Q9 | Q10 | Q12 | Q12_15_TEXT_chatGPT category |
|---|---|---|---|---|
| How much time passed between your initial diagnosis of metastatic ILC and the confirmation of metastatic disease? - Selected Choice | Was your metastatic ILC ever misdiagnosed as another condition throughout the history of your disease? | How many misdiagnoses have you received? This refers to instances where, during your workup, you were diagnosed with a condition other than metastatic ILC. | What was the misdiagnosis? - Selected Choice | What was the misdiagnosis? - p. Other (please specify) - Text |
| 1 | 2 | | | |
| 1 | 2 | 1 | | |
| 2 | 2 | | | |
| 2 | 2 | | | |
| 5 | 2 | | | |
| 1 | 2 | | | |
| 5 | 1 | 1 | 12 | |
| 5 | 2 | | | |
| 5 | 2 | 1 | 15 | |
| 1 | 2 | | | |

| Q8 | Q9 | Q10 | Q12 | Q12_15_TEXT_chatGPT category |
|---|---|---|---|---|
| How much time passed between your initial diagnosis of metastatic ILC and the confirmation of metastatic disease? - Selected Choice | Was your metastatic ILC ever misdiagnosed as another condition throughout the history of your disease? | How many misdiagnoses have you received? This refers to instances where, during your workup, you were diagnosed with a condition other than metastatic ILC. | What was the misdiagnosis? - Selected Choice | What was the misdiagnosis? - p. Other (please specify) - Text |
| 1 | 2 | | | |
| 1 | 1 | 1 | 15 | Bone related condition |
| 1 | 2 | | | |
| 5 | 1 | 1 | 15 | Bone related condition |
| 1 | 2 | 1 | 6 | |
| 5 | 2 | | | |

| Q8 | Q9 | Q10 | Q12 | Q12_15_TEXT_chatGPT category |
|---|---|---|---|---|
| How much time passed between your initial diagnosis of metastatic ILC and the confirmation of metastatic disease? - Selected Choice | Was your metastatic ILC ever misdiagnosed as another condition throughout the history of your disease? | How many misdiagnoses have you received? This refers to instances where, during your workup, you were diagnosed with a condition other than metastatic ILC. | What was the misdiagnosis? - Selected Choice | What was the misdiagnosis? - p. Other (please specify) - Text |
| 2 | 1 | 1 | 15 | Benign gynecological condition |
| 5 | 1 | 2 | 15 | Lymphatic system |
| 2 | 2 | 1 | 15 | Another type of breast cancer |

| Q8 | Q9 | Q10 | Q12 | Q12_15_TEXT_chatGPT category |
|---|---|---|---|---|
| How much time passed between your initial diagnosis of metastatic ILC and the confirmation of metastatic disease? - Selected Choice | Was your metastatic ILC ever misdiagnosed as another condition throughout the history of your disease? | How many misdiagnoses have you received? This refers to instances where, during your workup, you were diagnosed with a condition other than metastatic ILC. | What was the misdiagnosis? - Selected Choice | What was the misdiagnosis? - p. Other (please specify) - Text |
| | 1 | 1 | 6 | |
| 2 | 2 | | | |
| 1 | 2 | 1 | 15 | Hydronephrosis |
| 5 | 1 | 1 | 12 | |
| 5 | 1 | 1 | 15 | Gastrointestinal-related |
| 1 | 2 | | | |
| 1 | 2 | | | |
| 1 | 2 | | | |

| Q8 | Q9 | Q10 | Q12 | Q12_15_TEXT_chatGPT category |
|---|---|---|---|---|
| How much time passed between your initial diagnosis of metastatic ILC and the confirmation of metastatic disease? - Selected Choice | Was your metastatic ILC ever misdiagnosed as another condition throughout the history of your disease? | How many misdiagnoses have you received? This refers to instances where, during your workup, you were diagnosed with a condition other than metastatic ILC. | What was the misdiagnosis? - Selected Choice | What was the misdiagnosis? - p. Other (please specify) - Text |
| 1 | 2 | | | |
| 1 | 2 | 1 | 15 | |
| 1 | 1 | 2 | 15 | Benign breast condition |
| 1 | 1 | 1 | 12 | |
| 1 | 1 | 2 | 5 | |
| 1 | 2 | | | |
| 2 | 2 | 1 | | |

| Q8 | Q9 | Q10 | Q12 | Q12_15_TEXT_chatGPT category |
|---|---|---|---|---|
| How much time passed between your initial diagnosis of metastatic ILC and the confirmation of metastatic disease? - Selected Choice | Was your metastatic ILC ever misdiagnosed as another condition throughout the history of your disease? | How many misdiagnoses have you received? This refers to instances where, during your workup, you were diagnosed with a condition other than metastatic ILC. | What was the misdiagnosis? - Selected Choice | What was the misdiagnosis? - p. Other (please specify) - Text |
| 1 | 1 | 1 | 15 | |
| 2 | 2 | 1 | 15 | |
| 2 | 2 | | | |
| 5 | 2 | | | |
| 5 | 1 | 2 | 10 | |
| 2 | 1 | 1 | 6 | |
| 5 | 2 | 1 | 15 | |

| Q8 | Q9 | Q10 | Q12 | Q12_15_TEXT_chatGPT category |
|---|---|---|---|---|
| How much time passed between your initial diagnosis of metastatic ILC and the confirmation of metastatic disease? - Selected Choice | Was your metastatic ILC ever misdiagnosed as another condition throughout the history of your disease? | How many misdiagnoses have you received? This refers to instances where, during your workup, you were diagnosed with a condition other than metastatic ILC. | What was the misdiagnosis? - Selected Choice | What was the misdiagnosis? - p. Other (please specify) - Text |
| 1 | 1 | 2 | 15 | Bone related condition |
| 5 | 1 | 1 | 15 | Bone related condition |
| 5 | 2 | 1 | 7 | |
| 2 | 2 | | | |
| 1 | 2 | 1 | 6 | |
| 2 | 2 | | | |
| 1 | 2 | | 15 | |

| Q8 | Q9 | Q10 | Q12 | Q12_15_TEXT_chatGPT category |
|---|---|---|---|---|
| How much time passed between your initial diagnosis of metastatic ILC and the confirmation of metastatic disease? - Selected Choice | Was your metastatic ILC ever misdiagnosed as another condition throughout the history of your disease? | How many misdiagnoses have you received? This refers to instances where, during your workup, you were diagnosed with a condition other than metastatic ILC. | What was the misdiagnosis? - Selected Choice | What was the misdiagnosis? - p. Other (please specify) - Text |
| 5 | 2 | | | |
| 2 | 2 | | | |
| 1 | 2 | | | |
| 2 | 2 | | | |
| 5 | 2 | | | |
| 5 | 2 | 1 | | |
| 2 | 2 | | | |
| 1 | 2 | | | |
| 5 | 2 | | | |
| 1 | 2 | | 15 | |
| 1 | 2 | 1 | | |
| 2 | 2 | | | |
| 1 | 2 | | | |
| | 2 | | | |
| 2 | 2 | | | |
| 5 | 2 | | 15 | |

| Q8 | Q9 | Q10 | Q12 | Q12_15_TEXT_chatGPT category |
|---|---|---|---|---|
| How much time passed between your initial diagnosis of metastatic ILC and the confirmation of metastatic disease? - Selected Choice | Was your metastatic ILC ever misdiagnosed as another condition throughout the history of your disease? | How many misdiagnoses have you received? This refers to instances where, during your workup, you were diagnosed with a condition other than metastatic ILC. | What was the misdiagnosis? - Selected Choice | What was the misdiagnosis? - p. Other (please specify) - Text |
| 1 | 1 | 1 | 15 | Bone related condition |
| 1 | 2 | | | |
| 2 | 2 | | 5 | |
| 1 | 2 | | | |
| 1 | 2 | | | |
| 1 | 2 | 1 | | |
| 5 | 2 | | | |
| 5 | 2 | 1 | | |
| 1 | 2 | 1 | | |
| 2 | 1 | 1 | 5 | |
| 1 | 2 | | | |
| 2 | 2 | | 9 | |

| Q8 | Q9 | Q10 | Q12 | Q12_15_TEXT_chatGPT category |
| --- | --- | --- | --- | --- |
| How much time passed between your initial diagnosis of metastatic ILC and the confirmation of metastatic disease? - Selected Choice | Was your metastatic ILC ever misdiagnosed as another condition throughout the history of your disease? | How many misdiagnoses have you received? This refers to instances where, during your workup, you were diagnosed with a condition other than metastatic ILC. | What was the misdiagnosis? - Selected Choice | What was the misdiagnosis? - p. Other (please specify) - Text |
| 5 | 2 | | | |
| 1 | 2 | 1 | 5 | |
| 5 | 2 | | | |
| 5 | 1 | 1 | 15 | Others |
| 4 | 2 | 1 | 12 | |
| 3 | 2 | 1 | | |
| 5 | 2 | | | |
| 2 | 2 | | | |
| 4 | 1 | 2 | 15 | Primary gastrointestinal cancer |
| 4 | 2 | | | |
| 1 | 2 | | | |
| 5 | 2 | 1 | 11 | |
| 5 | 2 | | | |
| 1 | 2 | | | |
| 5 | 2 | 1 | | |
| 5 | 2 | | | |

| Q8 | Q9 | Q10 | Q12 | Q12_15_TEXT_chatGPT category |
|---|---|---|---|---|
| How much time passed between your initial diagnosis of metastatic ILC and the confirmation of metastatic disease? - Selected Choice | Was your metastatic ILC ever misdiagnosed as another condition throughout the history of your disease? | How many misdiagnoses have you received? This refers to instances where, during your workup, you were diagnosed with a condition other than metastatic ILC. | What was the misdiagnosis? - Selected Choice | What was the misdiagnosis? - p. Other (please specify) - Text |
| 5 | 1 | 1 | 15 | Bone related condition |
| 5 | 2 | 1 | | |
| 2 | 2 | | | |
| 1 | 2 | | | |
| 1 | 2 | 1 | 5 | |
| 1 | 1 | 2 | | |
| 1 | 2 | | | |
| 1 | 2 | | | |
| 3 | 1 | 1 | 2 | |
| 4 | 2 | | | |
| 5 | 2 | | | |
| 5 | 2 | | | |

| Q8 | Q9 | Q10 | Q12 | Q12_15_TEXT_chatGPT category |
|---|---|---|---|---|
| How much time passed between your initial diagnosis of metastatic ILC and the confirmation of metastatic disease? - Selected Choice | Was your metastatic ILC ever misdiagnosed as another condition throughout the history of your disease? | How many misdiagnoses have you received? This refers to instances where, during your workup, you were diagnosed with a condition other than metastatic ILC. | What was the misdiagnosis? - Selected Choice | What was the misdiagnosis? - p. Other (please specify) - Text |
| 1 | 2 | | | |
| 1 | 2 | 1 | 12 | |
| 2 | 2 | | | |
| 1 | 2 | | | |
| 1 | 1 | 1 | 9 | |
| 3 | 1 | 1 | 5 | |
| 1 | 2 | 1 | 5 | |
| 1 | 2 | | | |
| 1 | 2 | | | |
| 1 | 2 | | | |

| Q8 | Q9 | Q10 | Q12 | Q12_15_TEXT_chatGPT category |
|---|---|---|---|---|
| How much time passed between your initial diagnosis of metastatic ILC and the confirmation of metastatic disease? - Selected Choice | Was your metastatic ILC ever misdiagnosed as another condition throughout the history of your disease? | How many misdiagnoses have you received? This refers to instances where, during your workup, you were diagnosed with a condition other than metastatic ILC. | What was the misdiagnosis? - Selected Choice | What was the misdiagnosis? - p. Other (please specify) - Text |
| 2 | 2 | | | |
| 5 | 2 | | | |
| 2 | 1 | 2 | 5 | |
| 1 | 1 | 2 | 15 | Benign breast condition |
| 1 | 1 | 2 | | |
| 2 | 2 | | | |
| 5 | 2 | | | |
| 5 | 2 | | | |
| 1 | 2 | 1 | | |
| 1 | 2 | | | |
| 3 | 2 | | | |
| 5 | 2 | 1 | | |

| Q8 | Q9 | Q10 | Q12 | Q12_15_TEXT_chatGPT category |
|---|---|---|---|---|
| How much time passed between your initial diagnosis of metastatic ILC and the confirmation of metastatic disease? - Selected Choice | Was your metastatic ILC ever misdiagnosed as another condition throughout the history of your disease? | How many misdiagnoses have you received? This refers to instances where, during your workup, you were diagnosed with a condition other than metastatic ILC. | What was the misdiagnosis? - Selected Choice | What was the misdiagnosis? - p. Other (please specify) - Text |
| 5 | 1 | 2 | | |
| 1 | 1 | 1 | 5 | |
| 5 | 2 | | | |
| 1 | 2 | | | |
| 5 | 2 | 1 | | |
| 1 | 1 | 1 | 9 | |
| 1 | 2 | | | |
| 2 | 2 | | | |

| Q8 | Q9 | Q10 | Q12 | Q12_15_TEXT_chatGPT category |
|---|---|---|---|---|
| How much time passed between your initial diagnosis of metastatic ILC and the confirmation of metastatic disease? - Selected Choice | Was your metastatic ILC ever misdiagnosed as another condition throughout the history of your disease? | How many misdiagnoses have you received? This refers to instances where, during your workup, you were diagnosed with a condition other than metastatic ILC. | What was the misdiagnosis? - Selected Choice | What was the misdiagnosis? - p. Other (please specify) - Text |
| 5 | 2 | | | |
| 1 | 1 | 1 | 15 | Bone related condition |

| Q8 | Q9 | Q10 | Q12 | Q12_15_TEXT_chatGPT category |
|---|---|---|---|---|
| How much time passed between your initial diagnosis of metastatic ILC and the confirmation of metastatic disease? - Selected Choice | Was your metastatic ILC ever misdiagnosed as another condition throughout the history of your disease? | How many misdiagnoses have you received? This refers to instances where, during your workup, you were diagnosed with a condition other than metastatic ILC. | What was the misdiagnosis? - Selected Choice | What was the misdiagnosis? - p. Other (please specify) - Text |
| 2 | 2 | | | |
| 1 | 1 | 1 | 5 | |

| Q8 | Q9 | Q10 | Q12 | Q12_15_TEXT_chatGPT category |
|---|---|---|---|---|
| How much time passed between your initial diagnosis of metastatic ILC and the confirmation of metastatic disease? - Selected Choice | Was your metastatic ILC ever misdiagnosed as another condition throughout the history of your disease? | How many misdiagnoses have you received? This refers to instances where, during your workup, you were diagnosed with a condition other than metastatic ILC. | What was the misdiagnosis? - Selected Choice | What was the misdiagnosis? - p. Other (please specify) - Text |
| 1 | 1 | 1 | 2 | |
| 5 | 2 | | | |
| 2 | 1 | 1 | 15 | Primary gastrointestinal cancer |
| 2 | 2 | | | |

| Q8 | Q9 | Q10 | Q12 | Q12_15_TEXT_chatGPT category |
| --- | --- | --- | --- | --- |
| How much time passed between your initial diagnosis of metastatic ILC and the confirmation of metastatic disease? - Selected Choice | Was your metastatic ILC ever misdiagnosed as another condition throughout the history of your disease? | How many misdiagnoses have you received? This refers to instances where, during your workup, you were diagnosed with a condition other than metastatic ILC. | What was the misdiagnosis? - Selected Choice | What was the misdiagnosis? - p. Other (please specify) - Text |
| 2 | 2 | 2 | | |
| 1 | 2 | | | |

| Q8 | Q9 | Q10 | Q12 | Q12_15_TEXT_chatGPT category |
|---|---|---|---|---|
| How much time passed between your initial diagnosis of metastatic ILC and the confirmation of metastatic disease? - Selected Choice | Was your metastatic ILC ever misdiagnosed as another condition throughout the history of your disease? | How many misdiagnoses have you received? This refers to instances where, during your workup, you were diagnosed with a condition other than metastatic ILC. | What was the misdiagnosis? - Selected Choice | What was the misdiagnosis? - p. Other (please specify) - Text |
| 5 | 1 | 1 | 15 | Another type of breast cancer |
| 3 | 2 | | | |
| 2 | 1 | 2 | 15 | Menopause-related |
| 1 | 2 | | | |
| 1 | 2 | | | |

| Q8 | Q9 | Q10 | Q12 | Q12_15_TEXT_chatGPT category |
|---|---|---|---|---|
| How much time passed between your initial diagnosis of metastatic ILC and the confirmation of metastatic disease? - Selected Choice | Was your metastatic ILC ever misdiagnosed as another condition throughout the history of your disease? | How many misdiagnoses have you received? This refers to instances where, during your workup, you were diagnosed with a condition other than metastatic ILC. | What was the misdiagnosis? - Selected Choice | What was the misdiagnosis? - p. Other (please specify) - Text |
| 5 | 2 | | | |
| 1 | 1 | 2 | 12 | |
| 1 | 1 | 2 | 15 | Bone related condition |

| Q13 | Q14 | Q15 | Q16 |
| --- | --- | --- | --- |
| How long did it take to receive the correct metastatic ILC diagnosis after the misdiagnosis? | Do you feel there was a delay in diagnosing or identifying the progression of your metastatic ILC? | If yes, what do you believe contributed to the delay? (Select all that apply) - Selected Choice | Did you experience any symptoms before your metastatic ILC diagnosis or progression? |
| 5 | 1 | 7 | 1 |
| 2 | 1 | 1,2,6 | |
| 2 | 2 | 8 | 2 |
| 2 | 1 | 3,7 | 1 |
| 5 | 1 | 1 | 1 |
| 5 | 1 | 1,2,6 | 1 |

| Q13 | Q14 | Q15 | Q16 |
| --- | --- | --- | --- |
| How long did it take to receive the correct metastatic ILC diagnosis after the misdiagnosis? | Do you feel there was a delay in diagnosing or identifying the progression of your metastatic ILC? | If yes, what do you believe contributed to the delay? (Select all that apply) - Selected Choice | Did you experience any symptoms before your metastatic ILC diagnosis or progression? |
| 5 | 1 | 1,2,6,7 | 1 |
| 1 | 2 | 3 | |
| 1 | 2 | 8 | |
| 2 | 1 | 1,2 | 1 |
| | 1 | 2,3,7 | 2 |
| | | 8 | 1 |
| 1 | 2 | | 1 |
| | 2 | | 1 |
| 5 | 1 | 1 | 1 |
| | 2 | 8 | 1 |

| Q13 | Q14 | Q15 | Q16 |
| --- | --- | --- | --- |
| **How long did it take to receive the correct metastatic ILC diagnosis after the misdiagnosis?** | **Do you feel there was a delay in diagnosing or identifying the progression of your metastatic ILC?** | **If yes, what do you believe contributed to the delay? (Select all that apply) - Selected Choice** | **Did you experience any symptoms before your metastatic ILC diagnosis or progression?** |
| 3 | 1 | 6 | 1 |
| 1 | 2 | | 2 |
| 4 | 1 | 1 | 1 |
| | | | 2 |
| | | | 2 |
| | 2 | | 2 |
| | 2 | | 1 |

| Q13 | Q14 | Q15 | Q16 |
| --- | --- | --- | --- |
| How long did it take to receive the correct metastatic ILC diagnosis after the misdiagnosis? | Do you feel there was a delay in diagnosing or identifying the progression of your metastatic ILC? | If yes, what do you believe contributed to the delay? (Select all that apply) - Selected Choice | Did you experience any symptoms before your metastatic ILC diagnosis or progression? |
| | 1 | 3,6 | 1 |
| 4 | 2 | | 1 |
| | 1 | 7 | 1 |
| 5 | 1 | 1,3,5,6 | 1 |
| 2 | 1 | 5 | 1 |
| | 2 | | 1 |
| 4 | 1 | 6 | 1 |
| | 2 | 8 | 2 |

| Q13 | Q14 | Q15 | Q16 |
| --- | --- | --- | --- |
| How long did it take to receive the correct metastatic ILC diagnosis after the misdiagnosis? | Do you feel there was a delay in diagnosing or identifying the progression of your metastatic ILC? | If yes, what do you believe contributed to the delay? (Select all that apply) - Selected Choice | Did you experience any symptoms before your metastatic ILC diagnosis or progression? |
| 1 | 1 | 2,6 | 1 |
| 2 | 1 | 6 | 1 |
| 1 | 2 | 8 | 2 |
|  | 2 |  | 2 |
| 1 | 2 | 8 | 1 |
| 3 | 1 | 6,7 | 1 |
| 4 | 1 | 6 | 2 |

| Q13 | Q14 | Q15 | Q16 |
|---|---|---|---|
| How long did it take to receive the correct metastatic ILC diagnosis after the misdiagnosis? | Do you feel there was a delay in diagnosing or identifying the progression of your metastatic ILC? | If yes, what do you believe contributed to the delay? (Select all that apply) - Selected Choice | Did you experience any symptoms before your metastatic ILC diagnosis or progression? |
| | 1 | 7 | 1 |
| | 2 | 8 | 1 |
| | 2 | 2,7 | 2 |
| | 1 | 7 | 1 |

| Q13 | Q14 | Q15 | Q16 |
|---|---|---|---|
| How long did it take to receive the correct metastatic ILC diagnosis after the misdiagnosis? | Do you feel there was a delay in diagnosing or identifying the progression of your metastatic ILC? | If yes, what do you believe contributed to the delay? (Select all that apply) - Selected Choice | Did you experience any symptoms before your metastatic ILC diagnosis or progression? |
| | 1 | 7 | 2 |
| 1 | 2 | 8 | 1 |
| 5 | 1 | 1,6 | 1 |
| | | | 1 |
| | 2 | | 1 |
| | 2 | | 1 |
| | 2 | 8 | 2 |
| 5 | 1 | 1,2,6 | 2 |

| Q13 | Q14 | Q15 | Q16 |
|---|---|---|---|
| How long did it take to receive the correct metastatic ILC diagnosis after the misdiagnosis? | Do you feel there was a delay in diagnosing or identifying the progression of your metastatic ILC? | If yes, what do you believe contributed to the delay? (Select all that apply) - Selected Choice | Did you experience any symptoms before your metastatic ILC diagnosis or progression? |
| 5 | 1 | 8 | |
| 1 | 2 | | 2 |
| 5 | 1 | 7 | |
| 2 | 1 | 4 | |
| 2 | 1 | 1 | |
| | | | 2 |
| | 2 | | 1 |
| | 1 | 5 | 1 |
| | 1 | 3 | |
| | 2 | 8 | 1 |
| | | 8 | 2 |
| | 1 | 6 | |
| 5 | 1 | 1,2 | |
| 1 | 2 | 8 | |

| Q13 | Q14 | Q15 | Q16 |
|---|---|---|---|
| How long did it take to receive the correct metastatic ILC diagnosis after the misdiagnosis? | Do you feel there was a delay in diagnosing or identifying the progression of your metastatic ILC? | If yes, what do you believe contributed to the delay? (Select all that apply) - Selected Choice | Did you experience any symptoms before your metastatic ILC diagnosis or progression? |
| 5 | 1 | 1,2,7 | 1 |
| | | | 1 |
| | | 8 | 2 |
| 5 | 1 | 7 | |
| | 1 | 2 | 1 |
| | 1 | 7 | 2 |
| | 2 | | |
| | 1 | 2 | |
| | 2 | | |

| Q13 | Q14 | Q15 | Q16 |
|---|---|---|---|
| How long did it take to receive the correct metastatic ILC diagnosis after the misdiagnosis? | Do you feel there was a delay in diagnosing or identifying the progression of your metastatic ILC? | If yes, what do you believe contributed to the delay? (Select all that apply) - Selected Choice | Did you experience any symptoms before your metastatic ILC diagnosis or progression? |
| 2 | 2 | | |
| 1 | 2 | | 2 |
| | | | 2 |
| | 2 | 8 | 2 |
| | 1 | 2,4,7 | 1 |
| | 2 | 8 | 1 |

| Q13 | Q14 | Q15 | Q16 |
|---|---|---|---|
| How long did it take to receive the correct metastatic ILC diagnosis after the misdiagnosis? | Do you feel there was a delay in diagnosing or identifying the progression of your metastatic ILC? | If yes, what do you believe contributed to the delay? (Select all that apply) - Selected Choice | Did you experience any symptoms before your metastatic ILC diagnosis or progression? |
| | 1 | 6 | 2 |
| 2 | 2 | | 1 |
| 5 | 1 | 1 | 2 |
| | | | 2 |
| | 1 | 2 | 2 |
| 4 | 1 | 7 | 1 |
| | 2 | 8 | 1 |

| Q13 | Q14 | Q15 | Q16 |
| --- | --- | --- | --- |
| How long did it take to receive the correct metastatic ILC diagnosis after the misdiagnosis? | Do you feel there was a delay in diagnosing or identifying the progression of your metastatic ILC? | If yes, what do you believe contributed to the delay? (Select all that apply) - Selected Choice | Did you experience any symptoms before your metastatic ILC diagnosis or progression? |
|  | 2 | 8 | 2 |
| 5 | 1 | 6,7 | 1 |
| 3 | 1 | 6 | 1 |
|  | 2 |  | 1 |
| 2 | 1 | 7 | 1 |
| 4 | 1 | 6 | 1 |

| Q13 | Q14 | Q15 | Q16 |
|---|---|---|---|
| How long did it take to receive the correct metastatic ILC diagnosis after the misdiagnosis? | Do you feel there was a delay in diagnosing or identifying the progression of your metastatic ILC? | If yes, what do you believe contributed to the delay? (Select all that apply) - Selected Choice | Did you experience any symptoms before your metastatic ILC diagnosis or progression? |
| 2 | 1 | 1,3,5,6,7 | 2 |
| 3 | 1 | 1,2,7 | 1 |
|  | 1 | 3 | 2 |
| 5 | 1 | 2 | 2 |
| 2 | 1 | 1 | 1 |

| Q13 | Q14 | Q15 | Q16 |
|---|---|---|---|
| **How long did it take to receive the correct metastatic ILC diagnosis after the misdiagnosis?** | **Do you feel there was a delay in diagnosing or identifying the progression of your metastatic ILC?** | **If yes, what do you believe contributed to the delay? (Select all that apply) - Selected Choice** | **Did you experience any symptoms before your metastatic ILC diagnosis or progression?** |
| 4 | 1 | 1,2,3,6 | 1 |
| 1 | 2 | 8 | 1 |
| 5 | 1 | 1,2,6 | 1 |
| | 1 | 2 | 1 |
| 4 | 1 | 6 | 1 |
| | 2 | 8 | 2 |
| 3 | 1 | 2 | 1 |

| Q13 | Q14 | Q15 | Q16 |
|---|---|---|---|
| How long did it take to receive the correct metastatic ILC diagnosis after the misdiagnosis? | Do you feel there was a delay in diagnosing or identifying the progression of your metastatic ILC? | If yes, what do you believe contributed to the delay? (Select all that apply) - Selected Choice | Did you experience any symptoms before your metastatic ILC diagnosis or progression? |
| | | | 1 |
| | 2 | 8 | 1 |
| | | | 1 |
| | 2 | 8 | 2 |
| 5 | 1 | 3,6,7 | 1 |

| Q13 | Q14 | Q15 | Q16 |
|---|---|---|---|
| How long did it take to receive the correct metastatic ILC diagnosis after the misdiagnosis? | Do you feel there was a delay in diagnosing or identifying the progression of your metastatic ILC? | If yes, what do you believe contributed to the delay? (Select all that apply) - Selected Choice | Did you experience any symptoms before your metastatic ILC diagnosis or progression? |
| 4 | 1 | 1,4,7 | 1 |
| 3 | 1 | 2,3,5,6 | 2 |
| | 1 | 3,6 | 2 |
| 3 | 2 | | 2 |
| 2 | 1 | 1,7 | 2 |
| | 2 | 8 | 1 |
| 1 | 2 | 2 | 2 |
| 1 | 2 | 8 | 1 |
| 5 | 1 | 1,6 | 1 |

| Q13 | Q14 | Q15 | Q16 |
| --- | --- | --- | --- |
| How long did it take to receive the correct metastatic ILC diagnosis after the misdiagnosis? | Do you feel there was a delay in diagnosing or identifying the progression of your metastatic ILC? | If yes, what do you believe contributed to the delay? (Select all that apply) - Selected Choice | Did you experience any symptoms before your metastatic ILC diagnosis or progression? |
| 5 | 1 | 1,7 | 1 |
| 2 | 1 | 1,2,6 | 2 |
| 5 | 1 | 1,2 | 1 |
| 5 | 2 | 8 | 1 |

| Q13 | Q14 | Q15 | Q16 |
| --- | --- | --- | --- |
| **How long did it take to receive the correct metastatic ILC diagnosis after the misdiagnosis?** | **Do you feel there was a delay in diagnosing or identifying the progression of your metastatic ILC?** | **If yes, what do you believe contributed to the delay? (Select all that apply) - Selected Choice** | **Did you experience any symptoms before your metastatic ILC diagnosis or progression?** |
| 4 | 1 | 1,7 | 1 |
| 3 | 1 | 2 | 1 |
| 4 | 1 | 6 | 1 |
| 1 | 2 | | 1 |
| 2 | 2 | 2,8 | 1 |
| | | | 2 |
| 5 | 1 | 2 | 1 |
| 5 | 2 | 6 | 1 |

| Q13 | Q14 | Q15 | Q16 |
| --- | --- | --- | --- |
| How long did it take to receive the correct metastatic ILC diagnosis after the misdiagnosis? | Do you feel there was a delay in diagnosing or identifying the progression of your metastatic ILC? | If yes, what do you believe contributed to the delay? (Select all that apply) - Selected Choice | Did you experience any symptoms before your metastatic ILC diagnosis or progression? |
| | | | 1 |
| 1 | 2 | 8 | 2 |
| 1 | 2 | 8 | 2 |
| 4 | 1 | 1,2 | 1 |

| Q13 | Q14 | Q15 | Q16 |
| --- | --- | --- | --- |
| **How long did it take to receive the correct metastatic ILC diagnosis after the misdiagnosis?** | **Do you feel there was a delay in diagnosing or identifying the progression of your metastatic ILC?** | **If yes, what do you believe contributed to the delay? (Select all that apply) - Selected Choice** | **Did you experience any symptoms before your metastatic ILC diagnosis or progression?** |
| | 2 | | 2 |
| 1 | 2 | 8 | 2 |
| | | | 1 |
| | 2 | 8 | 2 |
| | 2 | | 2 |
| | | | 1 |
| 4 | 1 | 1 | 1 |
| | 2 | | 1 |
| 2 | 1 | 2,6 | 1 |
| | 1 | 2 | 1 |

| Q13 | Q14 | Q15 | Q16 |
| --- | --- | --- | --- |
| How long did it take to receive the correct metastatic ILC diagnosis after the misdiagnosis? | Do you feel there was a delay in diagnosing or identifying the progression of your metastatic ILC? | If yes, what do you believe contributed to the delay? (Select all that apply) - Selected Choice | Did you experience any symptoms before your metastatic ILC diagnosis or progression? |
| | 2 | | 1 |
| 2 | 1 | 3,7 | 1 |
| | 2 | | 1 |
| 5 | 1 | 1,2,6 | 1 |
| 2 | 1 | 6 | 2 |
| | 2 | | 1 |

| Q13 | Q14 | Q15 | Q16 |
| --- | --- | --- | --- |
| How long did it take to receive the correct metastatic ILC diagnosis after the misdiagnosis? | Do you feel there was a delay in diagnosing or identifying the progression of your metastatic ILC? | If yes, what do you believe contributed to the delay? (Select all that apply) - Selected Choice | Did you experience any symptoms before your metastatic ILC diagnosis or progression? |
| 3 | 1 | 2,6 | 1 |
| 5 | 1 | 1,2,5,7 | 1 |
| 2 | 1 | 6,7 | 1 |

| Q13 | Q14 | Q15 | Q16 |
| --- | --- | --- | --- |
| How long did it take to receive the correct metastatic ILC diagnosis after the misdiagnosis? | Do you feel there was a delay in diagnosing or identifying the progression of your metastatic ILC? | If yes, what do you believe contributed to the delay? (Select all that apply) - Selected Choice | Did you experience any symptoms before your metastatic ILC diagnosis or progression? |
| 5 | 1 | 2 | 1 |
|  | 2 |  | 1 |
| 1 | 2 | 8 | 1 |
| 5 | 1 | 1,2,4,6,7 | 2 |
| 2 | 1 | 4,5 | 1 |
| 1 | 2 | 8 | 2 |
| 1 | 1 | 2,7 | 2 |
|  | 2 | 8 | 2 |

| Q13 | Q14 | Q15 | Q16 |
| --- | --- | --- | --- |
| How long did it take to receive the correct metastatic ILC diagnosis after the misdiagnosis? | Do you feel there was a delay in diagnosing or identifying the progression of your metastatic ILC? | If yes, what do you believe contributed to the delay? (Select all that apply) - Selected Choice | Did you experience any symptoms before your metastatic ILC diagnosis or progression? |
| | 2 | | 1 |
| 1 | 2 | | 2 |
| 5 | 1 | 1 | 2 |
| 5 | 1 | 1,2,6 | 2 |
| 2 | 1 | 1 | 1 |

| Q13 | Q14 | Q15 | Q16 |
|---|---|---|---|
| How long did it take to receive the correct metastatic ILC diagnosis after the misdiagnosis? | Do you feel there was a delay in diagnosing or identifying the progression of your metastatic ILC? | If yes, what do you believe contributed to the delay? (Select all that apply) - Selected Choice | Did you experience any symptoms before your metastatic ILC diagnosis or progression? |
| 5 | 1 | 1 | 1 |
| 3 | 1 | 1,2,3,5,6,7 | 1 |
| | 1 | 4,7 | 2 |
| | 1 | 2 | 1 |
| 4 | 1 | 1,2,6 | 1 |
| 2 | 1 | 1,2,6 | 1 |
| 5 | 1 | 2 | 1 |

| Q13 | Q14 | Q15 | Q16 |
| --- | --- | --- | --- |
| How long did it take to receive the correct metastatic ILC diagnosis after the misdiagnosis? | Do you feel there was a delay in diagnosing or identifying the progression of your metastatic ILC? | If yes, what do you believe contributed to the delay? (Select all that apply) - Selected Choice | Did you experience any symptoms before your metastatic ILC diagnosis or progression? |
| 2 | 1 | 2,3,6 | 1 |
| 4 | 1 | 2,6 | 2 |
| 5 | 1 | 2 | 1 |
|  | 1 | 7 | 1 |
| 1 | 2 |  |  |
|  | 2 |  | 1 |
|  | 2 | 8 | 2 |

| Q13 | Q14 | Q15 | Q16 |
|---|---|---|---|
| How long did it take to receive the correct metastatic ILC diagnosis after the misdiagnosis? | Do you feel there was a delay in diagnosing or identifying the progression of your metastatic ILC? | If yes, what do you believe contributed to the delay? (Select all that apply) - Selected Choice | Did you experience any symptoms before your metastatic ILC diagnosis or progression? |
| | 2 | | |
| | | | 2 |
| | 2 | 8 | |
| | 2 | | 2 |
| | 2 | | |
| | 2 | | |
| 1 | 2 | 8 | 2 |
| | 2 | | |
| | | 3 | 2 |
| 5 | 1 | 2 | |

| Q13 | Q14 | Q15 | Q16 |
|---|---|---|---|
| **How long did it take to receive the correct metastatic ILC diagnosis after the misdiagnosis?** | **Do you feel there was a delay in diagnosing or identifying the progression of your metastatic ILC?** | **If yes, what do you believe contributed to the delay? (Select all that apply) - Selected Choice** | **Did you experience any symptoms before your metastatic ILC diagnosis or progression?** |
| 3 | 1 | 1,6 | 1 |
| 5 | 1 | 2,7 | 1 |
| 2 | 1 | 3,6 | 1 |
| | | 8 | |
| | 2 | | 2 |
| 1 | 2 | | 1 |
| 1 | 1 | 2 | |
| 5 | 1 | 2 | 1 |
| 1 | 2 | 8 | |

| Q13 | Q14 | Q15 | Q16 |
| --- | --- | --- | --- |
| How long did it take to receive the correct metastatic ILC diagnosis after the misdiagnosis? | Do you feel there was a delay in diagnosing or identifying the progression of your metastatic ILC? | If yes, what do you believe contributed to the delay? (Select all that apply) - Selected Choice | Did you experience any symptoms before your metastatic ILC diagnosis or progression? |
| 2 | 1 | 2,4 | 1 |
| 5 | 1 | 2 | |
| 1 | 1 | 7 | |
| 3 | 1 | 1,2 | |
| | 2 | | 1 |
| 4 | 1 | 1,2,6 | |
| | | 2,6 | |
| 4 | 2 | 8 | |
| | 1 | 7 | |
| | 1 | 7 | |

| Q13 | Q14 | Q15 | Q16 |
|---|---|---|---|
| **How long did it take to receive the correct metastatic ILC diagnosis after the misdiagnosis?** | **Do you feel there was a delay in diagnosing or identifying the progression of your metastatic ILC?** | **If yes, what do you believe contributed to the delay? (Select all that apply) - Selected Choice** | **Did you experience any symptoms before your metastatic ILC diagnosis or progression?** |
| 5 | 1 | 1,6,7 | 1 |
| | | | 1 |
| 3 | 2 | | 1 |
| | 2 | | |
| 4 | 1 | 1,2,3,6 | |

| Q13 | Q14 | Q15 | Q16 |
|---|---|---|---|
| How long did it take to receive the correct metastatic ILC diagnosis after the misdiagnosis? | Do you feel there was a delay in diagnosing or identifying the progression of your metastatic ILC? | If yes, what do you believe contributed to the delay? (Select all that apply) - Selected Choice | Did you experience any symptoms before your metastatic ILC diagnosis or progression? |
| 2 | 1 | 2 | |
| | 2 | 8 | 1 |
| 1 | 2 | | |
| 2 | 2 | 8 | 1 |
| 1 | 1 | | |
| 4 | 1 | 1 | |
| | 2 | | |
| | 1 | 5,6 | |
| | 1 | 2 | 2 |

| Q13 | Q14 | Q15 | Q16 |
|---|---|---|---|
| How long did it take to receive the correct metastatic ILC diagnosis after the misdiagnosis? | Do you feel there was a delay in diagnosing or identifying the progression of your metastatic ILC? | If yes, what do you believe contributed to the delay? (Select all that apply) - Selected Choice | Did you experience any symptoms before your metastatic ILC diagnosis or progression? |
|  | 2 |  | 1 |
| 5 | 1 | 1,2,3,4,5,6,7 | 1 |
| 4 | 1 | 1,2,3,5,6 | 1 |
|  | 2 |  | 2 |
|  |  |  | 1 |
| 1 | 2 | 8 | 1 |

| Q13 | Q14 | Q15 | Q16 |
| --- | --- | --- | --- |
| **How long did it take to receive the correct metastatic ILC diagnosis after the misdiagnosis?** | **Do you feel there was a delay in diagnosing or identifying the progression of your metastatic ILC?** | **If yes, what do you believe contributed to the delay? (Select all that apply) - Selected Choice** | **Did you experience any symptoms before your metastatic ILC diagnosis or progression?** |
| | | 2,6 | |
| 2 | 1 | 1 | |
| | 1 | 3,6 | 1 |
| 2 | 2 | 8 | |
| | 1 | 7 | 1 |

| Q13 | Q14 | Q15 | Q16 |
| --- | --- | --- | --- |
| How long did it take to receive the correct metastatic ILC diagnosis after the misdiagnosis? | Do you feel there was a delay in diagnosing or identifying the progression of your metastatic ILC? | If yes, what do you believe contributed to the delay? (Select all that apply) - Selected Choice | Did you experience any symptoms before your metastatic ILC diagnosis or progression? |
| | 1 | 2 | 1 |
| 4 | 1 | 1,6,7 | 1 |

| Q13 | Q14 | Q15 | Q16 |
|---|---|---|---|
| How long did it take to receive the correct metastatic ILC diagnosis after the misdiagnosis? | Do you feel there was a delay in diagnosing or identifying the progression of your metastatic ILC? | If yes, what do you believe contributed to the delay? (Select all that apply) - Selected Choice | Did you experience any symptoms before your metastatic ILC diagnosis or progression? |
| 5 | 1 | 6 | 2 |
| 5 | 1 | 1,2,6 | 2 |

| Q13 | Q14 | Q15 | Q16 |
| --- | --- | --- | --- |
| How long did it take to receive the correct metastatic ILC diagnosis after the misdiagnosis? | Do you feel there was a delay in diagnosing or identifying the progression of your metastatic ILC? | If yes, what do you believe contributed to the delay? (Select all that apply) - Selected Choice | Did you experience any symptoms before your metastatic ILC diagnosis or progression? |
| 1 | 2 | | 1 |
| | 2 | 8 | 2 |
| 2 | 1 | 4 | 1 |
| | 2 | | 2 |

| Q13 | Q14 | Q15 | Q16 |
| --- | --- | --- | --- |
| **How long did it take to receive the correct metastatic ILC diagnosis after the misdiagnosis?** | **Do you feel there was a delay in diagnosing or identifying the progression of your metastatic ILC?** | **If yes, what do you believe contributed to the delay? (Select all that apply) - Selected Choice** | **Did you experience any symptoms before your metastatic ILC diagnosis or progression?** |
| 3 | 1 | 1,2 | 1 |
| | | 8 | 2 |

| Q13 | Q14 | Q15 | Q16 |
| --- | --- | --- | --- |
| How long did it take to receive the correct metastatic ILC diagnosis after the misdiagnosis? | Do you feel there was a delay in diagnosing or identifying the progression of your metastatic ILC? | If yes, what do you believe contributed to the delay? (Select all that apply) - Selected Choice | Did you experience any symptoms before your metastatic ILC diagnosis or progression? |
| 5 | 1 | 1,2,7 | 1 |
| | 2 | 8 | 2 |
| 5 | 1 | 6 | 1 |
| | 2 | 8 | 1 |
| 4 | 1 | 2,7 | 1 |

| Q13 | Q14 | Q15 | Q16 |
| --- | --- | --- | --- |
| How long did it take to receive the correct metastatic ILC diagnosis after the misdiagnosis? | Do you feel there was a delay in diagnosing or identifying the progression of your metastatic ILC? | If yes, what do you believe contributed to the delay? (Select all that apply) - Selected Choice | Did you experience any symptoms before your metastatic ILC diagnosis or progression? |
| 1 | 1 | 2,3 | 1 |
| 1 | 2 | 2 | 1 |
| 3 | 1 | 7 | 1 |

| Q17 | Q18 | Q19 | Q20 | Q21 | Q22 |
|---|---|---|---|---|---|
| If yes, what symptoms did you experience? (Select all that apply) - Selected Choice | Were your symptoms diagnosed as a primary gastrointestinal/genitourinary/hematological/neurological condition? - Selected Choice | If yes, what was the gastrointestinal initial misdiagnosis? - Selected Choice | What was the initial genitourinary misdiagnosis? - Selected Choice | What was the initial neurologic misdiagnosis? - Selected Choice | What was the initial hematological misdiagnosis? - Selected Choice |
| 3,7,8,10,11,29,31,32,33 | 2 | 15 | | 4 | 1 |
| 35 | 2 | | | | |
| 31 | 2 | | | | |
| 4,5,6,7,9,11,31,42 | 2 | | | | |
| 3,5,6,8,9,16,25,31,40 | 1 | 15 | 6 | | 4 |

| Q17 | Q18 | Q19 | Q20 | Q21 | Q22 |
|---|---|---|---|---|---|
| If yes, what symptoms did you experience? (Select all that apply) - Selected Choice | Were your symptoms diagnosed as a primary gastrointestinal/genitourinary/hematological/neurological condition? - Selected Choice | If yes, what was the gastrointestinal initial misdiagnosis? - Selected Choice | What was the initial genitourinary misdiagnosis? - Selected Choice | What was the initial neurologic misdiagnosis? - Selected Choice | What was the initial hematological misdiagnosis? - Selected Choice |
| 3,5,6,7,8,10,11,19 | 2 | | | | |
| 20,38,51 | 2 | | | 3 | |
| 32 | 2 | | | | |
| 50,52 | 2 | | | 1 | |
| 31,32,36 | 1 | 10 | 5 | 3 | |
| 20 | | | | | |
| 29,40 | 2 | | | | |
| 39 | 2 | 11 | | | |

| Q17 | Q18 | Q19 | Q20 | Q21 | Q22 |
|---|---|---|---|---|---|
| If yes, what symptoms did you experience? (Select all that apply) - Selected Choice | Were your symptoms diagnosed as a primary gastrointestinal/genitourinary/hematological/neurological condition? - Selected Choice | If yes, what was the gastrointestinal initial misdiagnosis? - Selected Choice | What was the initial genitourinary misdiagnosis? - Selected Choice | What was the initial neurologic misdiagnosis? - Selected Choice | What was the initial hematological misdiagnosis? - Selected Choice |
| 17,32 | 2 | | | | |
| | 2 | | 6 | | |
| 3,6,8,11,19,22,25,29 | 1 | 11 | 1 | | |
| | 2 | | | | |
| | 2 | | | | |
| 11,20 | 2 | | | | |

| Q17 | Q18 | Q19 | Q20 | Q21 | Q22 |
|---|---|---|---|---|---|
| If yes, what symptoms did you experience? (Select all that apply) - Selected Choice | Were your symptoms diagnosed as a primary gastrointestinal/genitourinary/hematological/neurological condition? - Selected Choice | If yes, what was the gastrointestinal initial misdiagnosis? - Selected Choice | What was the initial genitourinary misdiagnosis? - Selected Choice | What was the initial neurologic misdiagnosis? - Selected Choice | What was the initial hematological misdiagnosis? - Selected Choice |
| 35 | | | | | |
| | 2 | | | | |
| 7,9,22,32 | 2 | | | | |
| 20,22,25,31,32,33 | 2 | | | | |
| 11 | | | | | |
| 40 | 2 | | | | |
| 3,4,5,8,11,16,19,31,35 | 1 | 11 | | | |

| Q17 | Q18 | Q19 | Q20 | Q21 | Q22 |
| --- | --- | --- | --- | --- | --- |
| **If yes, what symptoms did you experience? (Select all that apply) - Selected Choice** | **Were your symptoms diagnosed as a primary gastrointestinal/genitourinary/hematological/neurological condition? - Selected Choice** | **If yes, what was the gastrointestinal initial misdiagnosis? - Selected Choice** | **What was the initial genitourinary misdiagnosis? - Selected Choice** | **What was the initial neurologic misdiagnosis? - Selected Choice** | **What was the initial hematological misdiagnosis? - Selected Choice** |
| 31,33,40,47 | 2 | | | | |
| 3,4,5,6,31,33,49,52 | 1 | 1 | | | |
| 29 | 2 | | | | |
| | 2 | | | | |
| 32 | 2 | | | | |
| [illegible],7,8,9,10,11,15,16,19,22,25,[illegible] | 1 | 15 | 6 | | |
| | 2 | | | | |

| Q17 | Q18 | Q19 | Q20 | Q21 | Q22 |
|---|---|---|---|---|---|
| If yes, what symptoms did you experience? (Select all that apply) - Selected Choice | Were your symptoms diagnosed as a primary gastrointestinal/genitourinary/hematological/neurological condition? - Selected Choice | If yes, what was the gastrointestinal initial misdiagnosis? - Selected Choice | What was the initial genitourinary misdiagnosis? - Selected Choice | What was the initial neurologic misdiagnosis? - Selected Choice | What was the initial hematological misdiagnosis? - Selected Choice |
| 20,35 | 2 | | | | |
| 16,17,31,32,37,51 | 2 | | | | 3 |
| 3,6,7,8,16,30,32,43,45 | 1 | 9 | | | |

| Q17 | Q18 | Q19 | Q20 | Q21 | Q22 |
|---|---|---|---|---|---|
| If yes, what symptoms did you experience? (Select all that apply) - Selected Choice | Were your symptoms diagnosed as a primary gastrointestinal/genitourinary/hematological/neurological condition? - Selected Choice | If yes, what was the gastrointestinal initial misdiagnosis? - Selected Choice | What was the initial genitourinary misdiagnosis? - Selected Choice | What was the initial neurologic misdiagnosis? - Selected Choice | What was the initial hematological misdiagnosis? - Selected Choice |
| 20 | 1 | 4 | | | |
| 5,6,8,9,16,31 | 2 | | | | |
| 32,53 | 2 | | | | |
| 3,4,5,6,7,15,16,28,29,36 | 1 | 2 | | | |
| 31 | 2 | | | | |
| | 2 | | | | |

| Q17 | Q18 | Q19 | Q20 | Q21 | Q22 |
|---|---|---|---|---|---|
| If yes, what symptoms did you experience? (Select all that apply) - Selected Choice | Were your symptoms diagnosed as a primary gastrointestinal/genitourinary/hematological/neurological condition? - Selected Choice | If yes, what was the gastrointestinal initial misdiagnosis? - Selected Choice | What was the initial genitourinary misdiagnosis? - Selected Choice | What was the initial neurologic misdiagnosis? - Selected Choice | What was the initial hematological misdiagnosis? - Selected Choice |
| | 2 | | | | |
| 8,10,31,32 | 2 | | | | |
| 11 | 2 | | | | |
| 29,30,32 | 2 | | | | |

| Q17 | Q18 | Q19 | Q20 | Q21 | Q22 |
|---|---|---|---|---|---|
| If yes, what symptoms did you experience? (Select all that apply) - Selected Choice | Were your symptoms diagnosed as a primary gastrointestinal/genitourinary/hematological/neurological condition? - Selected Choice | If yes, what was the gastrointestinal initial misdiagnosis? - Selected Choice | What was the initial genitourinary misdiagnosis? - Selected Choice | What was the initial neurologic misdiagnosis? - Selected Choice | What was the initial hematological misdiagnosis? - Selected Choice |
| 3,7,8,10,11,29,31,33 | 2 | | | | 4 |
| 20,32 | 2 | | | | |
| | 2 | | | | |
| 28,32,36,40 | 2 | | | | |

| Q17 | Q18 | Q19 | Q20 | Q21 | Q22 |
| --- | --- | --- | --- | --- | --- |
| **If yes, what symptoms did you experience? (Select all that apply) - Selected Choice** | **Were your symptoms diagnosed as a primary gastrointestinal/genitourinary/hematological/neurological condition? - Selected Choice** | **If yes, what was the gastrointestinal initial misdiagnosis? - Selected Choice** | **What was the initial genitourinary misdiagnosis? - Selected Choice** | **What was the initial neurologic misdiagnosis? - Selected Choice** | **What was the initial hematological misdiagnosis? - Selected Choice** |
| 11,22,25,29,31,32,35 | 2 | | | | 1 |
| 20 | 2 | | | | |

| Q17 | Q18 | Q19 | Q20 | Q21 | Q22 |
|---|---|---|---|---|---|
| If yes, what symptoms did you experience? (Select all that apply) - Selected Choice | Were your symptoms diagnosed as a primary gastrointestinal/genitourinary/hematological/neurological condition? - Selected Choice | If yes, what was the gastrointestinal initial misdiagnosis? - Selected Choice | What was the initial genitourinary misdiagnosis? - Selected Choice | What was the initial neurologic misdiagnosis? - Selected Choice | What was the initial hematological misdiagnosis? - Selected Choice |
| | 2 | | | | |
| 51 | 2 | | | | |
| | 2 | | | | |
| 20 | 2 | | | | |
| 26,28,29,30,31,32,33,39,41, | 1 | 12 | 1 | 3 | 3 |

| Q17 | Q18 | Q19 | Q20 | Q21 | Q22 |
| --- | --- | --- | --- | --- | --- |
| **If yes, what symptoms did you experience? (Select all that apply) - Selected Choice** | **Were your symptoms diagnosed as a primary gastrointestinal/genitourinary/hematological/neurological condition? - Selected Choice** | **If yes, what was the gastrointestinal initial misdiagnosis? - Selected Choice** | **What was the initial genitourinary misdiagnosis? - Selected Choice** | **What was the initial neurologic misdiagnosis? - Selected Choice** | **What was the initial hematological misdiagnosis? - Selected Choice** |
| 31,32 | 2 | | | | |
| 3,10,19,29,31,37 | 2 | | | | 3 |
| 3,8,11,19,29,51 | 2 | | | 4 | |
| 20,29,31,32,35,52 | 2 | | | | |
| 11,32 | 2 | | | | |

| Q17 | Q18 | Q19 | Q20 | Q21 | Q22 |
|---|---|---|---|---|---|
| If yes, what symptoms did you experience? (Select all that apply) - Selected Choice | Were your symptoms diagnosed as a primary gastrointestinal/genitourinary/hematological/neurological condition? - Selected Choice | If yes, what was the gastrointestinal initial misdiagnosis? - Selected Choice | What was the initial genitourinary misdiagnosis? - Selected Choice | What was the initial neurologic misdiagnosis? - Selected Choice | What was the initial hematological misdiagnosis? - Selected Choice |
| 31,32,50,52 | 2 | 15 | | 4 | |
| 28,32 | 2 | | | | |

| Q17 | Q18 | Q19 | Q20 | Q21 | Q22 |
| --- | --- | --- | --- | --- | --- |
| If yes, what symptoms did you experience? (Select all that apply) - Selected Choice | Were your symptoms diagnosed as a primary gastrointestinal/genitourinary/hematological/neurological condition? - Selected Choice | If yes, what was the gastrointestinal initial misdiagnosis? - Selected Choice | What was the initial genitourinary misdiagnosis? - Selected Choice | What was the initial neurologic misdiagnosis? - Selected Choice | What was the initial hematological misdiagnosis? - Selected Choice |
| 20,29,31,34,36 | 2 | | | | |
| 50,53 | 2 | | | 4 | |
| 32,33,34,51,53 | 2 | | | | |
| 20,35 | 2 | | | | |
| 32,41,42 | 2 | | | | 2 |
| 19,21,22,25,31,33,34,37,39, | 1 | 11 | 1 | 3 | 3 |

| Q17 | Q18 | Q19 | Q20 | Q21 | Q22 |
| --- | --- | --- | --- | --- | --- |
| If yes, what symptoms did you experience? (Select all that apply) - Selected Choice | Were your symptoms diagnosed as a primary gastrointestinal/genitourinary/hematological/neurological condition? - Selected Choice | If yes, what was the gastrointestinal initial misdiagnosis? - Selected Choice | What was the initial genitourinary misdiagnosis? - Selected Choice | What was the initial neurologic misdiagnosis? - Selected Choice | What was the initial hematological misdiagnosis? - Selected Choice |
| 4,20,32,49 | 2 | | | 4 | |
| 4,20,44 | 2 | 15 | | | |
| 22,31 | 2 | | | | |
| | 2 | | | | |
| 22,31,33,34,35,38,39,40,47,4 | 2 | 15 | 3 | | |

| Q17 | Q18 | Q19 | Q20 | Q21 | Q22 |
|---|---|---|---|---|---|
| If yes, what symptoms did you experience? (Select all that apply) - Selected Choice | Were your symptoms diagnosed as a primary gastrointestinal/genitourinary/hematological/neurological condition? - Selected Choice | If yes, what was the gastrointestinal initial misdiagnosis? - Selected Choice | What was the initial genitourinary misdiagnosis? - Selected Choice | What was the initial neurologic misdiagnosis? - Selected Choice | What was the initial hematological misdiagnosis? - Selected Choice |
| 20,36 | 2 | | | | |
| | 2 | | | | |
| | 2 | | 5 | | |
| | 2 | | | | |
| 31,32 | 2 | | | | |
| 36,40,49,53 | 2 | | | | |
| 47 | 2 | | | | |

| Q17 | Q18 | Q19 | Q20 | Q21 | Q22 |
|---|---|---|---|---|---|
| If yes, what symptoms did you experience? (Select all that apply) - Selected Choice | Were your symptoms diagnosed as a primary gastrointestinal/genitourinary/hematological/neurological condition? - Selected Choice | If yes, what was the gastrointestinal initial misdiagnosis? - Selected Choice | What was the initial genitourinary misdiagnosis? - Selected Choice | What was the initial neurologic misdiagnosis? - Selected Choice | What was the initial hematological misdiagnosis? - Selected Choice |
| 3,6,8,19,25,29,31,32,33,49 | 2 | | 6 | 4 | |
| | 2 | | | | |
| 6,20,31,33,40,52 | 1 | 15 | | 4 | |
| 41,50 | 2 | | | | |

| Q17 | Q18 | Q19 | Q20 | Q21 | Q22 |
|---|---|---|---|---|---|
| If yes, what symptoms did you experience? (Select all that apply) - Selected Choice | Were your symptoms diagnosed as a primary gastrointestinal/genitourinary/hematological/neurological condition? - Selected Choice | If yes, what was the gastrointestinal initial misdiagnosis? - Selected Choice | What was the initial genitourinary misdiagnosis? - Selected Choice | What was the initial neurologic misdiagnosis? - Selected Choice | What was the initial hematological misdiagnosis? - Selected Choice |
| 25 | 2 | | 6 | 4 | 4 |
| 5,32,33,42 | 1 | 9 | 5 | 1 | 3 |
| 6,16,31,32,40 | 2 | | | | 2 |
| 6,9,16,17,20,32 | 2 | | | | |
| | 2 | | | | |
| 3 | 1 | 15 | | | |
| 4,5,6,7,15,36 | 1 | 2 | 5 | | |

| Q17 | Q18 | Q19 | Q20 | Q21 | Q22 |
|---|---|---|---|---|---|
| If yes, what symptoms did you experience? (Select all that apply) - Selected Choice | Were your symptoms diagnosed as a primary gastrointestinal/genitourinary/hematological/neurological condition? - Selected Choice | If yes, what was the gastrointestinal initial misdiagnosis? - Selected Choice | What was the initial genitourinary misdiagnosis? - Selected Choice | What was the initial neurologic misdiagnosis? - Selected Choice | What was the initial hematological misdiagnosis? - Selected Choice |
| 28,40 | | | | | |
| | 2 | 15 | 6 | 4 | 4 |
| 20,32 | 2 | | | | |

| Q17 | Q18 | Q19 | Q20 | Q21 | Q22 |
|---|---|---|---|---|---|
| If yes, what symptoms did you experience? (Select all that apply) - Selected Choice | Were your symptoms diagnosed as a primary gastrointestinal/genitourinary/hematological/neurological condition? - Selected Choice | If yes, what was the gastrointestinal initial misdiagnosis? - Selected Choice | What was the initial genitourinary misdiagnosis? - Selected Choice | What was the initial neurologic misdiagnosis? - Selected Choice | What was the initial hematological misdiagnosis? - Selected Choice |
| | 2 | | | | |
| 25,39,46 | | | 1 | | |
| 20,31,32 | 2 | | | | |
| | 2 | | | | |
| 31,32 | 2 | | | | |
| 32 | 2 | | | | |
| 3,6,7,9,11,15,16,36 | 1 | 4 | | | |
| 20,28,31,38 | 2 | | | | |
| 35,40 | 2 | | | | |

| Q17 | Q18 | Q19 | Q20 | Q21 | Q22 |
|---|---|---|---|---|---|
| If yes, what symptoms did you experience? (Select all that apply) - Selected Choice | Were your symptoms diagnosed as a primary gastrointestinal/genitourinary/hematological/neurological condition? - Selected Choice | If yes, what was the gastrointestinal initial misdiagnosis? - Selected Choice | What was the initial genitourinary misdiagnosis? - Selected Choice | What was the initial neurologic misdiagnosis? - Selected Choice | What was the initial hematological misdiagnosis? - Selected Choice |
| 32 | 2 | | | | 1 |
| 31,32,40,49 | 2 | | | 4 | |
| 3,5,7,9,16,23,29,50,53 | 1 | 15 | | 1 | |
| 40 | 2 | | | | |
| 20,52 | 2 | | | | |

| Q17 | Q18 | Q19 | Q20 | Q21 | Q22 |
|---|---|---|---|---|---|
| If yes, what symptoms did you experience? (Select all that apply) - Selected Choice | Were your symptoms diagnosed as a primary gastrointestinal/genitourinary/hematological/neurological condition? - Selected Choice | If yes, what was the gastrointestinal initial misdiagnosis? - Selected Choice | What was the initial genitourinary misdiagnosis? - Selected Choice | What was the initial neurologic misdiagnosis? - Selected Choice | What was the initial hematological misdiagnosis? - Selected Choice |
| ;,6,22,25,31,32,34,35,42,43, | 2 | 4 | 6 | 1 | 4 |
| 3,6,15,16,20,31,32,33 | 2 | 12 | 6 | | |
| 20,32,39 | 2 | | | | |

| Q17 | Q18 | Q19 | Q20 | Q21 | Q22 |
| --- | --- | --- | --- | --- | --- |
| **If yes, what symptoms did you experience? (Select all that apply) - Selected Choice** | **Were your symptoms diagnosed as a primary gastrointestinal/genitourinary/hematological/neurological condition? - Selected Choice** | **If yes, what was the gastrointestinal initial misdiagnosis? - Selected Choice** | **What was the initial genitourinary misdiagnosis? - Selected Choice** | **What was the initial neurologic misdiagnosis? - Selected Choice** | **What was the initial hematological misdiagnosis? - Selected Choice** |
| 3,4,5,6,11,29,31,36 | 2 | 4 | | | |
| 3,8,9,11,15,17,25,29,33 | 1 | | 6 | | |
| 27 | 1 | | 4 | | |
| | 2 | | | | |
| 3,4,6,9,11,16,29 | 2 | 12 | | | |
| | 2 | | | | |

| Q17 | Q18 | Q19 | Q20 | Q21 | Q22 |
| --- | --- | --- | --- | --- | --- |
| If yes, what symptoms did you experience? (Select all that apply) - Selected Choice | Were your symptoms diagnosed as a primary gastrointestinal/genitourinary/hematological/neurological condition? - Selected Choice | If yes, what was the gastrointestinal initial misdiagnosis? - Selected Choice | What was the initial genitourinary misdiagnosis? - Selected Choice | What was the initial neurologic misdiagnosis? - Selected Choice | What was the initial hematological misdiagnosis? - Selected Choice |
| 32,35 | 1 | | | | |
| | 1 | 9 | | | |
| 20 | 2 | | | | |

| Q17 | Q18 | Q19 | Q20 | Q21 | Q22 |
|---|---|---|---|---|---|
| If yes, what symptoms did you experience? (Select all that apply) - Selected Choice | Were your symptoms diagnosed as a primary gastrointestinal/genitourinary/hematological/neurological condition? - Selected Choice | If yes, what was the gastrointestinal initial misdiagnosis? - Selected Choice | What was the initial genitourinary misdiagnosis? - Selected Choice | What was the initial neurologic misdiagnosis? - Selected Choice | What was the initial hematological misdiagnosis? - Selected Choice |
| 20,27,31,32,49,52 | 2 | | | | |
| 20,31,32,49,52 | 2 | | | | 4 |
| | 2 | | | | |
| 53 | 2 | | | | |
| 3,4,5,6,9,11,15,16,36 | 1 | 9 | 4 | | |
| 3,32,51,52 | 2 | 15 | | | |
| 7,29 | 2 | | | | |

| Q17 | Q18 | Q19 | Q20 | Q21 | Q22 |
|---|---|---|---|---|---|
| If yes, what symptoms did you experience? (Select all that apply) - Selected Choice | Were your symptoms diagnosed as a primary gastrointestinal/genitourinary/hematological/neurological condition? - Selected Choice | If yes, what was the gastrointestinal initial misdiagnosis? - Selected Choice | What was the initial genitourinary misdiagnosis? - Selected Choice | What was the initial neurologic misdiagnosis? - Selected Choice | What was the initial hematological misdiagnosis? - Selected Choice |
| 10,16,32,38,42,43,45 | 1 | 9 | 1 | 1 | |
| | 2 | | | | |
| 4,32,51 | 2 | 4 | 1 | 3 | 3 |
| 20,29,31,32 | 2 | | | | |
| 3,11,25 | 2 | | | | |
| | 2 | | | | |

| Q17 | Q18 | Q19 | Q20 | Q21 | Q22 |
| --- | --- | --- | --- | --- | --- |
| If yes, what symptoms did you experience? (Select all that apply) - Selected Choice | Were your symptoms diagnosed as a primary gastrointestinal/genitourinary/hematological/neurological condition? - Selected Choice | If yes, what was the gastrointestinal initial misdiagnosis? - Selected Choice | What was the initial genitourinary misdiagnosis? - Selected Choice | What was the initial neurologic misdiagnosis? - Selected Choice | What was the initial hematological misdiagnosis? - Selected Choice |
| | 2 | | | | |
| 27,40 | 2 | 15 | 6 | 4 | 4 |

| Q17 | Q18 | Q19 | Q20 | Q21 | Q22 |
|---|---|---|---|---|---|
| If yes, what symptoms did you experience? (Select all that apply) - Selected Choice | Were your symptoms diagnosed as a primary gastrointestinal/genitourinary/hematological/neurological condition? - Selected Choice | If yes, what was the gastrointestinal initial misdiagnosis? - Selected Choice | What was the initial genitourinary misdiagnosis? - Selected Choice | What was the initial neurologic misdiagnosis? - Selected Choice | What was the initial hematological misdiagnosis? - Selected Choice |
| 29,32 | 2 | | | | |
| 3,11,20,29,31 | 1 | 11 | 6 | 4 | 3 |
| 20 | 2 | | | | |
| 33,34,51 | 2 | | | | |
| 35 | 1 | | | | |
| 31,33,34,51,52 | 2 | | | 3 | |

| Q17 | Q18 | Q19 | Q20 | Q21 | Q22 |
|---|---|---|---|---|---|
| If yes, what symptoms did you experience? (Select all that apply) - Selected Choice | Were your symptoms diagnosed as a primary gastrointestinal/genitourinary/hematological/neurological condition? - Selected Choice | If yes, what was the gastrointestinal initial misdiagnosis? - Selected Choice | What was the initial genitourinary misdiagnosis? - Selected Choice | What was the initial neurologic misdiagnosis? - Selected Choice | What was the initial hematological misdiagnosis? - Selected Choice |
| 11,29,30,31,32,49 | 2 | | 6 | | |
| 8,19 | 1 | 11 | | | |

| Q17 | Q18 | Q19 | Q20 | Q21 | Q22 |
|---|---|---|---|---|---|
| If yes, what symptoms did you experience? (Select all that apply) - Selected Choice | Were your symptoms diagnosed as a primary gastrointestinal/genitourinary/hematological/neurological condition? - Selected Choice | If yes, what was the gastrointestinal initial misdiagnosis? - Selected Choice | What was the initial genitourinary misdiagnosis? - Selected Choice | What was the initial neurologic misdiagnosis? - Selected Choice | What was the initial hematological misdiagnosis? - Selected Choice |
| 31,32,33,53 | 2 | | | | |
| 3,6,7,9,16,17,31,32,33 | 2 | 9 | | | |
| 32 | 2 | | | | |

| Q17 | Q18 | Q19 | Q20 | Q21 | Q22 |
|---|---|---|---|---|---|
| If yes, what symptoms did you experience? (Select all that apply) - Selected Choice | Were your symptoms diagnosed as a primary gastrointestinal/genitourinary/hematological/neurological condition? - Selected Choice | If yes, what was the gastrointestinal initial misdiagnosis? - Selected Choice | What was the initial genitourinary misdiagnosis? - Selected Choice | What was the initial neurologic misdiagnosis? - Selected Choice | What was the initial hematological misdiagnosis? - Selected Choice |
| 40 | 2 | | | | |
| 8,11,19,29,38,52 | 1 | 11 | | | |
| 29,31 | 2 | | | | |

| Q17 | Q18 | Q19 | Q20 | Q21 | Q22 |
|---|---|---|---|---|---|
| If yes, what symptoms did you experience? (Select all that apply) - Selected Choice | Were your symptoms diagnosed as a primary gastrointestinal/genitourinary/hematological/neurological condition? - Selected Choice | If yes, what was the gastrointestinal initial misdiagnosis? - Selected Choice | What was the initial genitourinary misdiagnosis? - Selected Choice | What was the initial neurologic misdiagnosis? - Selected Choice | What was the initial hematological misdiagnosis? - Selected Choice |
| 3,29,33 | 2 | | | | |
| 28,35,40 | | | | | |
| 43 | | 15 | | | |
| 21,27,40,44 | 2 | | 6 | | 4 |
| | 2 | | | | |
| 3,4,5,6,29,31,36 | 2 | | | | |
| 20,31,32 | 2 | | 6 | 4 | 4 |

| Q17 | Q18 | Q19 | Q20 | Q21 | Q22 |
|---|---|---|---|---|---|
| If yes, what symptoms did you experience? (Select all that apply) - Selected Choice | Were your symptoms diagnosed as a primary gastrointestinal/genitourinary/hematological/neurological condition? - Selected Choice | If yes, what was the gastrointestinal initial misdiagnosis? - Selected Choice | What was the initial genitourinary misdiagnosis? - Selected Choice | What was the initial neurologic misdiagnosis? - Selected Choice | What was the initial hematological misdiagnosis? - Selected Choice |
| 3,6,9,16,19,21,24,25,46 | 1 | | 1 | | |
| 20,32 | 2 | | | | |

| Q17 | Q18 | Q19 | Q20 | Q21 | Q22 |
|---|---|---|---|---|---|
| If yes, what symptoms did you experience? (Select all that apply) - Selected Choice | Were your symptoms diagnosed as a primary gastrointestinal/genitourinary/hematological/neurological condition? - Selected Choice | If yes, what was the gastrointestinal initial misdiagnosis? - Selected Choice | What was the initial genitourinary misdiagnosis? - Selected Choice | What was the initial neurologic misdiagnosis? - Selected Choice | What was the initial hematological misdiagnosis? - Selected Choice |
| 6,22 | 1 | 15 | 5 | | |
| ,21,25,26,28,31,35,37,41,44 | 1 | 15 | 1 | 4 | 4 |

| Q17 | Q18 | Q19 | Q20 | Q21 | Q22 |
| --- | --- | --- | --- | --- | --- |
| If yes, what symptoms did you experience? (Select all that apply) - Selected Choice | Were your symptoms diagnosed as a primary gastrointestinal/genitourinary/hematological/neurological condition? - Selected Choice | If yes, what was the gastrointestinal initial misdiagnosis? - Selected Choice | What was the initial genitourinary misdiagnosis? - Selected Choice | What was the initial neurologic misdiagnosis? - Selected Choice | What was the initial hematological misdiagnosis? - Selected Choice |
| | 2 | | | | |
| | 2 | | | | |

| Q17 | Q18 | Q19 | Q20 | Q21 | Q22 |
| --- | --- | --- | --- | --- | --- |
| If yes, what symptoms did you experience? (Select all that apply) - Selected Choice | Were your symptoms diagnosed as a primary gastrointestinal/genitourinary/hematological/neurological condition? - Selected Choice | If yes, what was the gastrointestinal initial misdiagnosis? - Selected Choice | What was the initial genitourinary misdiagnosis? - Selected Choice | What was the initial neurologic misdiagnosis? - Selected Choice | What was the initial hematological misdiagnosis? - Selected Choice |
| 3,9,20 | 1 | 15 | | | |
| | 2 | 15 | 6 | 4 | |
| 3,7,8,9,15,16,20,22,29,38 | 1 | 1 | | | |

| Q17 | Q18 | Q19 | Q20 | Q21 | Q22 |
|---|---|---|---|---|---|
| If yes, what symptoms did you experience? (Select all that apply) - Selected Choice | Were your symptoms diagnosed as a primary gastrointestinal/genitourinary/hematological/neurological condition? - Selected Choice | If yes, what was the gastrointestinal initial misdiagnosis? - Selected Choice | What was the initial genitourinary misdiagnosis? - Selected Choice | What was the initial neurologic misdiagnosis? - Selected Choice | What was the initial hematological misdiagnosis? - Selected Choice |
| 20 | 2 | 15 | 6 | 4 | 4 |

| Q17 | Q18 | Q19 | Q20 | Q21 | Q22 |
|---|---|---|---|---|---|
| If yes, what symptoms did you experience? (Select all that apply) - Selected Choice | Were your symptoms diagnosed as a primary gastrointestinal/genitourinary/hematological/neurological condition? - Selected Choice | If yes, what was the gastrointestinal initial misdiagnosis? - Selected Choice | What was the initial genitourinary misdiagnosis? - Selected Choice | What was the initial neurologic misdiagnosis? - Selected Choice | What was the initial hematological misdiagnosis? - Selected Choice |
| ,15,16,19,20,22,31,34,38,43 | 1 | 15 | | | |
| 31,40 | 2 | | | | |
| 4 | 2 | | | | |
| 48,50,51,52,53 | | | | | |

| Q17 | Q18 | Q19 | Q20 | Q21 | Q22 |
|---|---|---|---|---|---|
| If yes, what symptoms did you experience? (Select all that apply) - Selected Choice | Were your symptoms diagnosed as a primary gastrointestinal/genitourinary/hematological/neurological condition? - Selected Choice | If yes, what was the gastrointestinal initial misdiagnosis? - Selected Choice | What was the initial genitourinary misdiagnosis? - Selected Choice | What was the initial neurologic misdiagnosis? - Selected Choice | What was the initial hematological misdiagnosis? - Selected Choice |
| 20,52 | | | | | |
| 32 | 2 | | | | |
| 20,27 | | | | 4 | |

| Q23 | Q23_1_TEXT_chatGPT category | Q24 | Q25 | Q26 |
| --- | --- | --- | --- | --- |
| **Were you treated for the misdiagnosed condition? - Selected Choice** | **Were you treated for the misdiagnosed condition? - a. Yes, please specify treatment - Text** | **Were you referred to a specialist after the initial misdiagnosis?** | **If yes, what type of specialist were you referred to? - Selected Choice** | **What diagnostic tests were performed to identify the cause of your symptoms? (Select all that apply) - Selected Choice** |
| 2 | | 1 | 1 | 1,2,3,4,5,6,8 |
| 1 | | 1 | 8 | 3,5 |
| 2 | | 2 | | 3,7 |
| 1 | Hormonal or endocrine treatments | 2 | | 5,8 |
| 1 | | 2 | | 5,7 |

| Q23 | Q23_1_TEXT_chatGPT category | Q24 | Q25 | Q26 |
|---|---|---|---|---|
| Were you treated for the misdiagnosed condition? - Selected Choice | Were you treated for the misdiagnosed condition? - a. Yes, please specify treatment - Text | Were you referred to a specialist after the initial misdiagnosis? | If yes, what type of specialist were you referred to? - Selected Choice | What diagnostic tests were performed to identify the cause of your symptoms? (Select all that apply) - Selected Choice |
| | | | | 1,3,4 |
| 1 | Respiratory treatments | 1 | 8 | 4,7 |
| | | | | 4,6 |
| 2 | | 1 | 7 | 6 |
| 1 | | 1 | 5 | 3,5,8 |
| | | | | 3,5,6 |
| 2 | | 2 | | 7 |

| Q23 | Q23_1_TEXT_chatGPT category | Q24 | Q25 | Q26 |
| --- | --- | --- | --- | --- |
| Were you treated for the misdiagnosed condition? - Selected Choice | Were you treated for the misdiagnosed condition? - a. Yes, please specify treatment - Text | Were you referred to a specialist after the initial misdiagnosis? | If yes, what type of specialist were you referred to? - Selected Choice | What diagnostic tests were performed to identify the cause of your symptoms? (Select all that apply) - Selected Choice |
| | | | | 4,6 |
| 1 | | 2 | | 3,5 |
| 2 | | 1 | 1 | 1,2,3 |
| | | | | 6 |
| | | 2 | | |

| Q23 | Q23_1_TEXT_chatGPT category | Q24 | Q25 | Q26 |
| --- | --- | --- | --- | --- |
| Were you treated for the misdiagnosed condition? - Selected Choice | Were you treated for the misdiagnosed condition? - a. Yes, please specify treatment - Text | Were you referred to a specialist after the initial misdiagnosis? | If yes, what type of specialist were you referred to? - Selected Choice | What diagnostic tests were performed to identify the cause of your symptoms? (Select all that apply) - Selected Choice |
| | | | | 3,4,5,6,8 |
| 2 | | 2 | | 7 |
| | | | | 4,5,8 |
| 1 | | 1 | 2 | 4,6,8 |
| 2 | | 2 | | 1,2,3,4,6,8 |
| | | | 8 | 4,7 |
| 1 | GI-related treatments | 2 | | 1,3,4 |

| Q23 | Q23_1_TEXT_chatGPT category | Q24 | Q25 | Q26 |
|---|---|---|---|---|
| **Were you treated for the misdiagnosed condition? - Selected Choice** | **Were you treated for the misdiagnosed condition? - a. Yes, please specify treatment - Text** | **Were you referred to a specialist after the initial misdiagnosis?** | **If yes, what type of specialist were you referred to? - Selected Choice** | **What diagnostic tests were performed to identify the cause of your symptoms? (Select all that apply) - Selected Choice** |
| 2 | | 2 | | |
| 2 | | 1 | 1 | 1,2,3 |
| | | | | 3,4,5,6 |
| 1 | management | 2 | | 4,6 |
| 2 | | 1 | 2 | 1,4,6,8 |
| 2 | | 2 | | 4 |

| Q23 | Q23_1_TEXT_chatGPT category | Q24 | Q25 | Q26 |
|---|---|---|---|---|
| Were you treated for the misdiagnosed condition? - Selected Choice | Were you treated for the misdiagnosed condition? - a. Yes, please specify treatment - Text | Were you referred to a specialist after the initial misdiagnosis? | If yes, what type of specialist were you referred to? - Selected Choice | What diagnostic tests were performed to identify the cause of your symptoms? (Select all that apply) - Selected Choice |
| 2 | | 2 | | 3 |
| 1 | Hormonal or endocrine treatments | 2 | | 5,8 |
| 2 | | 2 | | |

| Q23 | Q23_1_TEXT_chatGPT category | Q24 | Q25 | Q26 |
|---|---|---|---|---|
| Were you treated for the misdiagnosed condition? - Selected Choice | Were you treated for the misdiagnosed condition? - a. Yes, please specify treatment - Text | Were you referred to a specialist after the initial misdiagnosis? | If yes, what type of specialist were you referred to? - Selected Choice | What diagnostic tests were performed to identify the cause of your symptoms? (Select all that apply) - Selected Choice |
| 1 | GI-related treatments | 2 | | 1 |
| 2 | | 2 | | 7 |
| 1 | GI-related treatments | 2 | 4 | 1,2,4<br>3,5,7 |
| 1 | Cancer-specific treatment given inappropriately or prematurely | 2 | | |

| Q23 | Q23_1_TEXT_chatGPT category | Q24 | Q25 | Q26 |
| --- | --- | --- | --- | --- |
| Were you treated for the misdiagnosed condition? - Selected Choice | Were you treated for the misdiagnosed condition? - a. Yes, please specify treatment - Text | Were you referred to a specialist after the initial misdiagnosis? | If yes, what type of specialist were you referred to? - Selected Choice | What diagnostic tests were performed to identify the cause of your symptoms? (Select all that apply) - Selected Choice |
| 1 | Cancer-specific treatment given inappropriately or prematurely | 1 | 5 | 3,5 |
| 2 | | 2 | | 1,2,7 |
| | | | | 1,2,3 |
| | | | | 7 |
| | | | | 4 |

| Q23 | Q23_1_TEXT_chatGPT category | Q24 | Q25 | Q26 |
|---|---|---|---|---|
| Were you treated for the misdiagnosed condition? - Selected Choice | Were you treated for the misdiagnosed condition? - a. Yes, please specify treatment - Text | Were you referred to a specialist after the initial misdiagnosis? | If yes, what type of specialist were you referred to? - Selected Choice | What diagnostic tests were performed to identify the cause of your symptoms? (Select all that apply) - Selected Choice |
| 2 | | 2 | | 1,3 |
| | | | | 4,6 |
| 2 | | 2 | | 4,5,6 |

| Q23 | Q23_1_TEXT_chatGPT category | Q24 | Q25 | Q26 |
|---|---|---|---|---|
| Were you treated for the misdiagnosed condition? - Selected Choice | Were you treated for the misdiagnosed condition? - a. Yes, please specify treatment - Text | Were you referred to a specialist after the initial misdiagnosis? | If yes, what type of specialist were you referred to? - Selected Choice | What diagnostic tests were performed to identify the cause of your symptoms? (Select all that apply) - Selected Choice |
| 2 | | | | |
| 2 | | | | 6,8 |
| | | | | 3,4 |

| Q23 | Q23_1_TEXT_chatGPT category | Q24 | Q25 | Q26 |
|---|---|---|---|---|
| Were you treated for the misdiagnosed condition? - Selected Choice | Were you treated for the misdiagnosed condition? - a. Yes, please specify treatment - Text | Were you referred to a specialist after the initial misdiagnosis? | If yes, what type of specialist were you referred to? - Selected Choice | What diagnostic tests were performed to identify the cause of your symptoms? (Select all that apply) - Selected Choice |
| 2 | | 2 | | 6 |
| 2 | | 2 | | |
| 2 | | | | |
| 1 | Stent | 1 | 3 | 3,4,8 |
| 1 | | 2 | | 1,2,3 |

| Q23 | Q23_1_TEXT_chatGPT category | Q24 | Q25 | Q26 |
|---|---|---|---|---|
| Were you treated for the misdiagnosed condition? - Selected Choice | Were you treated for the misdiagnosed condition? - a. Yes, please specify treatment - Text | Were you referred to a specialist after the initial misdiagnosis? | If yes, what type of specialist were you referred to? - Selected Choice | What diagnostic tests were performed to identify the cause of your symptoms? (Select all that apply) - Selected Choice |
| 1 | Musculoskeletal or pain management | 2 | | 8,7 |
| 2 | | 1 | 1 | 1,3,5 |
| 2 | | 1 | 1 | 1,2,3,8 |
| 2 | | 1 | 8 | 3,4 |
| 1 | Musculoskeletal or pain management | 1 | 2 | 4,6 |

| Q23 | Q23_1_TEXT_chatGPT category | Q24 | Q25 | Q26 |
|---|---|---|---|---|
| Were you treated for the misdiagnosed condition? - Selected Choice | Were you treated for the misdiagnosed condition? - a. Yes, please specify treatment - Text | Were you referred to a specialist after the initial misdiagnosis? | If yes, what type of specialist were you referred to? - Selected Choice | What diagnostic tests were performed to identify the cause of your symptoms? (Select all that apply) - Selected Choice |
| | | | | 3,4,5,6 |
| 1 | Cancer-specific treatment given inappropriately or prematurely | 1 | 2 | 4,6 |
| | | 1 | 5 | 3,6 |
| 1 | Musculoskeletal or pain management | 1 | 8 | 6,7 |

| Q23 | Q23_1_TEXT_chatGPT category | Q24 | Q25 | Q26 |
|---|---|---|---|---|
| Were you treated for the misdiagnosed condition? - Selected Choice | Were you treated for the misdiagnosed condition? - a. Yes, please specify treatment - Text | Were you referred to a specialist after the initial misdiagnosis? | If yes, what type of specialist were you referred to? - Selected Choice | What diagnostic tests were performed to identify the cause of your symptoms? (Select all that apply) - Selected Choice |
| 2 | | 2 | 8 | 5 |
| 2 | | 1 | 6 | 6,7 |
| | | | | 3,5 |
| 1 | | 1 | 2 | 3,4 |
| 2 | | 2 | | 6 |
| | | | | 3,4,5,6,8 |
| 1 | Antibiotics or infection-related | 1 | 2 | 4,5,6,7 |

| Q23 | Q23_1_TEXT_chatGPT category | Q24 | Q25 | Q26 |
|---|---|---|---|---|
| **Were you treated for the misdiagnosed condition? - Selected Choice** | **Were you treated for the misdiagnosed condition? - a. Yes, please specify treatment - Text** | **Were you referred to a specialist after the initial misdiagnosis?** | **If yes, what type of specialist were you referred to? - Selected Choice** | **What diagnostic tests were performed to identify the cause of your symptoms? (Select all that apply) - Selected Choice** |
| 2 | | 1 | 2 | 3,4,6,8 |
| 2 | | 1 | 2 | 3,4,5,6,8,7 |
| 2 | | | | 3,4,5,6,8 |
| 1 | GI-related treatments | 1 | 1 | 2,3,4,6,8 |

| Q23 | Q23_1_TEXT_chatGPT category | Q24 | Q25 | Q26 |
|---|---|---|---|---|
| Were you treated for the misdiagnosed condition? - Selected Choice | Were you treated for the misdiagnosed condition? - a. Yes, please specify treatment - Text | Were you referred to a specialist after the initial misdiagnosis? | If yes, what type of specialist were you referred to? - Selected Choice | What diagnostic tests were performed to identify the cause of your symptoms? (Select all that apply) - Selected Choice |
| 2 | | 2 | | 4 |
| 2 | | 2 | | 3,4,5,6 |
| 2 | | 2 | | 4,5 |
| 2 | | 1 | 2 | 3,4,5,6,8,7 |
| | | | | 4,6 |
| | | | | 1,2,4,6,8 |
| 1 | Musculoskeletal or pain management | 1 | 8 | 3,6 |
| 2 | | 2 | | |

| Q23 | Q23_1_TEXT_chatGPT category | Q24 | Q25 | Q26 |
|---|---|---|---|---|
| Were you treated for the misdiagnosed condition? - Selected Choice | Were you treated for the misdiagnosed condition? - a. Yes, please specify treatment - Text | Were you referred to a specialist after the initial misdiagnosis? | If yes, what type of specialist were you referred to? - Selected Choice | What diagnostic tests were performed to identify the cause of your symptoms? (Select all that apply) - Selected Choice |
| 1 | Hormonal or endocrine treatments | 2 | 8 | 3,4,8,7 |
| 2 | | 1 | 2 | 3,4,5,6,8 |
| 2 | | 1 | 6 | 8,7<br>3,4,5,6,7 |

| Q23 | Q23_1_TEXT_chatGPT category | Q24 | Q25 | Q26 |
|---|---|---|---|---|
| Were you treated for the misdiagnosed condition? - Selected Choice | Were you treated for the misdiagnosed condition? - a. Yes, please specify treatment - Text | Were you referred to a specialist after the initial misdiagnosis? | If yes, what type of specialist were you referred to? - Selected Choice | What diagnostic tests were performed to identify the cause of your symptoms? (Select all that apply) - Selected Choice |
| 2 | | 2 | | 3 |
| 2 | | 2 | | 4,6,8 |
| 2 | | 2 | | 3,4,6,7 |
| 2 | | | 4 | 3,4,6,8 |
| 2 | | | | |
| 1 | Musculoskeletal or pain management | 1 | 1 | 2 |
| 2 | | 1 | 4 | 1,2,3,4,5,6,8 |

| Q23 | Q23_1_TEXT_chatGPT category | Q24 | Q25 | Q26 |
|---|---|---|---|---|
| Were you treated for the misdiagnosed condition? - Selected Choice | Were you treated for the misdiagnosed condition? - a. Yes, please specify treatment - Text | Were you referred to a specialist after the initial misdiagnosis? | If yes, what type of specialist were you referred to? - Selected Choice | What diagnostic tests were performed to identify the cause of your symptoms? (Select all that apply) - Selected Choice |
| | | | | 4,5,6 |
| | | | | 6 |

| Q23 | Q23_1_TEXT_chatGPT category | Q24 | Q25 | Q26 |
| --- | --- | --- | --- | --- |
| Were you treated for the misdiagnosed condition? - Selected Choice | Were you treated for the misdiagnosed condition? - a. Yes, please specify treatment - Text | Were you referred to a specialist after the initial misdiagnosis? | If yes, what type of specialist were you referred to? - Selected Choice | What diagnostic tests were performed to identify the cause of your symptoms? (Select all that apply) - Selected Choice |
| 2 | | 2 | | 6 |
| 2 | | 2 | | 3,4,5,6,8,7 |
| 2 | | 2 | | 7 |
| 2 | | 1 | 2 | 3,4,5,6,8 |
| 1 | | 1 | 2 | 4 |
| | | | | 3 |

| Q23 | Q23_1_TEXT_chatGPT category | Q24 | Q25 | Q26 |
|---|---|---|---|---|
| Were you treated for the misdiagnosed condition? - Selected Choice | Were you treated for the misdiagnosed condition? - a. Yes, please specify treatment - Text | Were you referred to a specialist after the initial misdiagnosis? | If yes, what type of specialist were you referred to? - Selected Choice | What diagnostic tests were performed to identify the cause of your symptoms? (Select all that apply) - Selected Choice |
| 1 | Musculoskeletal or pain management | 1 | 8 | 6 |
| 1 | Musculoskeletal or pain management | 2 | | 7 |
| 1 | Ear/eye/neurologic-specific | 1 | 1 | 2,3,4,6,7 |
| 1 | Musculoskeletal or pain management | 1 | 8 | 3,6 |
| | | | | 3,4,7 |
| | | | | 6 |

| Q23 | Q23_1_TEXT_chatGPT category | Q24 | Q25 | Q26 |
|---|---|---|---|---|
| Were you treated for the misdiagnosed condition? - Selected Choice | Were you treated for the misdiagnosed condition? - a. Yes, please specify treatment - Text | Were you referred to a specialist after the initial misdiagnosis? | If yes, what type of specialist were you referred to? - Selected Choice | What diagnostic tests were performed to identify the cause of your symptoms? (Select all that apply) - Selected Choice |
| 2 | | 1 | 2 | 3,4,5,6,8,7 |
| 1 | GI-related treatments | 2 | | 2,5,8 |
| 2 | | 2 | | 7 |

| Q23 | Q23_1_TEXT_chatGPT category | Q24 | Q25 | Q26 |
|---|---|---|---|---|
| Were you treated for the misdiagnosed condition? - Selected Choice | Were you treated for the misdiagnosed condition? - a. Yes, please specify treatment - Text | Were you referred to a specialist after the initial misdiagnosis? | If yes, what type of specialist were you referred to? - Selected Choice | What diagnostic tests were performed to identify the cause of your symptoms? (Select all that apply) - Selected Choice |
| 2 | | 1 | 5 | 1,2,3,4,8 |
| 1 | | 2 | | 7 |
| 1 | Stent | 1 | 3 | 4,5 |
| 1 | inappropriately or prematurely | 2 | | 3 |
| 2 | | 2 | | 4,5 |
| 2 | | 2 | | |

| Q23 | Q23_1_TEXT_chatGPT category | Q24 | Q25 | Q26 |
|---|---|---|---|---|
| Were you treated for the misdiagnosed condition? - Selected Choice | Were you treated for the misdiagnosed condition? - a. Yes, please specify treatment - Text | Were you referred to a specialist after the initial misdiagnosis? | If yes, what type of specialist were you referred to? - Selected Choice | What diagnostic tests were performed to identify the cause of your symptoms? (Select all that apply) - Selected Choice |
| | | | | 3,4,5 |
| 2 | | 1 | 2 | 4 |
| 1 | | 2 | | 3,5 |
| 2 | | 2 | | |
| 2 | | 1 | 8 | 6 |

| Q23 | Q23_1_TEXT_chatGPT category | Q24 | Q25 | Q26 |
|---|---|---|---|---|
| Were you treated for the misdiagnosed condition? - Selected Choice | Were you treated for the misdiagnosed condition? - a. Yes, please specify treatment - Text | Were you referred to a specialist after the initial misdiagnosis? | If yes, what type of specialist were you referred to? - Selected Choice | What diagnostic tests were performed to identify the cause of your symptoms? (Select all that apply) - Selected Choice |
| 1 | Musculoskeletal or pain management | 1 | 8 | 7 |
| 2 | | 1 | 8 | 3,4,6 |
| 2 | | | | 3,4,6 |
| 1 | GI-related treatments | 1 | 1 | 1,2,3 |
| 2 | | 1 | 5 | 1,2,3 |
| 1 | Hormonal or endocrine treatments | 2 | | 7 |

| Q23 | Q23_1_TEXT_chatGPT category | Q24 | Q25 | Q26 |
|---|---|---|---|---|
| Were you treated for the misdiagnosed condition? - Selected Choice | Were you treated for the misdiagnosed condition? - a. Yes, please specify treatment - Text | Were you referred to a specialist after the initial misdiagnosis? | If yes, what type of specialist were you referred to? - Selected Choice | What diagnostic tests were performed to identify the cause of your symptoms? (Select all that apply) - Selected Choice |
| 1 | | 2 | | 1,2,3,8 |
| 2 | | 2 | | 7 |
| 2 | | 2 | | 1,3,8 |
| 1 | Antibiotics or infection-related | 1 | 8 | 8 |
| 2 | | 2 | | 7 |

| Q23 | Q23_1_TEXT_chatGPT category | Q24 | Q25 | Q26 |
|---|---|---|---|---|
| Were you treated for the misdiagnosed condition? - Selected Choice | Were you treated for the misdiagnosed condition? - a. Yes, please specify treatment - Text | Were you referred to a specialist after the initial misdiagnosis? | If yes, what type of specialist were you referred to? - Selected Choice | What diagnostic tests were performed to identify the cause of your symptoms? (Select all that apply) - Selected Choice |
| 2 | | 2 | | 6,7 |
| 2 | | 2 | 8 | 5 |

| Q23 | Q23_1_TEXT_chatGPT category | Q24 | Q25 | Q26 |
|---|---|---|---|---|
| Were you treated for the misdiagnosed condition? - Selected Choice | Were you treated for the misdiagnosed condition? - a. Yes, please specify treatment - Text | Were you referred to a specialist after the initial misdiagnosis? | If yes, what type of specialist were you referred to? - Selected Choice | What diagnostic tests were performed to identify the cause of your symptoms? (Select all that apply) - Selected Choice |
| 1 | Musculoskeletal or pain management | 1 | 8 | 6,7 |
| 2 | | 1 | 1 | 1,2,4 |
| 2 | | 1 | 8 | 7 |
| | | | | 8 |
| 1 | Ear/eye/neurologic-specific | 2 | | 4,5 |

| Q23 | Q23_1_TEXT_chatGPT category | Q24 | Q25 | Q26 |
| --- | --- | --- | --- | --- |
| Were you treated for the misdiagnosed condition? - Selected Choice | Were you treated for the misdiagnosed condition? - a. Yes, please specify treatment - Text | Were you referred to a specialist after the initial misdiagnosis? | If yes, what type of specialist were you referred to? - Selected Choice | What diagnostic tests were performed to identify the cause of your symptoms? (Select all that apply) - Selected Choice |
| | | | | 4,6 |
| 2 | | 2 | | |

| Q23 | Q23_1_TEXT_chatGPT category | Q24 | Q25 | Q26 |
|---|---|---|---|---|
| Were you treated for the misdiagnosed condition? - Selected Choice | Were you treated for the misdiagnosed condition? - a. Yes, please specify treatment - Text | Were you referred to a specialist after the initial misdiagnosis? | If yes, what type of specialist were you referred to? - Selected Choice | What diagnostic tests were performed to identify the cause of your symptoms? (Select all that apply) - Selected Choice |
| 1 | Musculoskeletal or pain management | 2 | | 8 |
| 2 | | 1 | 1 | 2 |
| 2 | | 2 | | |

| Q23 | Q23_1_TEXT_chatGPT category | Q24 | Q25 | Q26 |
|---|---|---|---|---|
| Were you treated for the misdiagnosed condition? - Selected Choice | Were you treated for the misdiagnosed condition? - a. Yes, please specify treatment - Text | Were you referred to a specialist after the initial misdiagnosis? | If yes, what type of specialist were you referred to? - Selected Choice | What diagnostic tests were performed to identify the cause of your symptoms? (Select all that apply) - Selected Choice |
| | | | | 7 |
| 1 | | 1 | 2 | 1,3,4,8 |

| Q23 | Q23_1_TEXT_chatGPT category | Q24 | Q25 | Q26 |
|---|---|---|---|---|
| Were you treated for the misdiagnosed condition? - Selected Choice | Were you treated for the misdiagnosed condition? - a. Yes, please specify treatment - Text | Were you referred to a specialist after the initial misdiagnosis? | If yes, what type of specialist were you referred to? - Selected Choice | What diagnostic tests were performed to identify the cause of your symptoms? (Select all that apply) - Selected Choice |
| | | | | 3,6,8,7 |
| 2 | | 1 | 3<br>8 | 4,5,6,8,7 |
| 2 | | 2 | | 2,3 |
| 2 | | 2 | 8 | 4,7 |

| Q23 | Q23_1_TEXT_chatGPT category | Q24 | Q25 | Q26 |
| --- | --- | --- | --- | --- |
| Were you treated for the misdiagnosed condition? - Selected Choice | Were you treated for the misdiagnosed condition? - a. Yes, please specify treatment - Text | Were you referred to a specialist after the initial misdiagnosis? | If yes, what type of specialist were you referred to? - Selected Choice | What diagnostic tests were performed to identify the cause of your symptoms? (Select all that apply) - Selected Choice |
| 2 | | 1 | 3 | 3,4,8,7 |
| 2 | | 1 | 2 | 3,6 |

| Q23 | Q23_1_TEXT_chatGPT category | Q24 | Q25 | Q26 |
| --- | --- | --- | --- | --- |
| Were you treated for the misdiagnosed condition? - Selected Choice | Were you treated for the misdiagnosed condition? - a. Yes, please specify treatment - Text | Were you referred to a specialist after the initial misdiagnosis? | If yes, what type of specialist were you referred to? - Selected Choice | What diagnostic tests were performed to identify the cause of your symptoms? (Select all that apply) - Selected Choice |
| | | | 2 | 4,7 |
| 1 | Hormonal or endocrine treatments | 1 | 8 | 1,3,4,5,6,8 |

| Q23 | Q23_1_TEXT_chatGPT category | Q24 | Q25 | Q26 |
|---|---|---|---|---|
| Were you treated for the misdiagnosed condition? - Selected Choice | Were you treated for the misdiagnosed condition? - a. Yes, please specify treatment - Text | Were you referred to a specialist after the initial misdiagnosis? | If yes, what type of specialist were you referred to? - Selected Choice | What diagnostic tests were performed to identify the cause of your symptoms? (Select all that apply) - Selected Choice |
| | | | | 3,4,5,6,8 |
| 2 | | 2 | | 5 |

| Q23 | Q23_1_TEXT_chatGPT category | Q24 | Q25 | Q26 |
|---|---|---|---|---|
| Were you treated for the misdiagnosed condition? - Selected Choice | Were you treated for the misdiagnosed condition? - a. Yes, please specify treatment - Text | Were you referred to a specialist after the initial misdiagnosis? | If yes, what type of specialist were you referred to? - Selected Choice | What diagnostic tests were performed to identify the cause of your symptoms? (Select all that apply) - Selected Choice |
| 1 | Cancer-specific treatment given inappropriately or prematurely | 1 | 2 | 8,7 |
| 2 | | 2 | 8 | 7 |
| 1 | Cancer-specific treatment given inappropriately or prematurely | 1 | 2 | 1,2,3,4,5,6,7 |
| 2 | | | | |

| Q23 | Q23_1_TEXT_chatGPT category | Q24 | Q25 | Q26 |
| --- | --- | --- | --- | --- |
| Were you treated for the misdiagnosed condition? - Selected Choice | Were you treated for the misdiagnosed condition? - a. Yes, please specify treatment - Text | Were you referred to a specialist after the initial misdiagnosis? | If yes, what type of specialist were you referred to? - Selected Choice | What diagnostic tests were performed to identify the cause of your symptoms? (Select all that apply) - Selected Choice |
| 1 | | 1 | 8 | 3,4,5,6,8,7 |

| Q23 | Q23_1_TEXT_chatGPT category | Q24 | Q25 | Q26 |
|---|---|---|---|---|
| Were you treated for the misdiagnosed condition? - Selected Choice | Were you treated for the misdiagnosed condition? - a. Yes, please specify treatment - Text | Were you referred to a specialist after the initial misdiagnosis? | If yes, what type of specialist were you referred to? - Selected Choice | What diagnostic tests were performed to identify the cause of your symptoms? (Select all that apply) - Selected Choice |
| 1 | GI-related treatments | 2 | | 7 |
| 2 | | 2 | | 7 |
| | | | | 3,4,6,8,7 |
| 2 | | 1 | 8 | 4,6 |

| Q23 | Q23_1_TEXT_chatGPT category | Q24 | Q25 | Q26 |
|---|---|---|---|---|
| Were you treated for the misdiagnosed condition? - Selected Choice | Were you treated for the misdiagnosed condition? - a. Yes, please specify treatment - Text | Were you referred to a specialist after the initial misdiagnosis? | If yes, what type of specialist were you referred to? - Selected Choice | What diagnostic tests were performed to identify the cause of your symptoms? (Select all that apply) - Selected Choice |
| | | | | 3,5,6,8 |
| 2 | | 1 | 2 | 4,6,8 |
| 1 | Ear/eye/neurologic-specific | 1 | 8 | 7 |

| Q26_7_TEXT | Q27 | Q28 | Q29 | Q30 | Q31 |
| --- | --- | --- | --- | --- | --- |
| What diagnostic tests were performed to identify the cause of your symptoms? (Select all that apply) - h. Other, please specify - Text | Were any of these tests positive for cancer? | If yes, which test(s) indicated cancer? (Select all that apply) - Selected Choice | If yes, which type of cancer were you told that you had? - Selected Choice | Were you treated for the misdiagnosed cancer? - Selected Choice | At the time of your misdiagnosis, did you have a mammogram as part of your advanced or metastatic ILC diagnostic process? (if you had ILC in the past, this question refers to the recent misdiagnosis period, not the initial diagnosis) |
| | 1 | 1,3,6,7 | 3 | 2 | 1 |
| | 1 | 3 | 13 | 1 | 2 |
| breast mRI | 1 | 7 | | 2 | |
| | 2 | | | 2 | 1 |
| that location, so di[illegible] | 2 | | | | 1 |

| Q26_7_TEXT | Q27 | Q28 | Q29 | Q30 | Q31 |
|---|---|---|---|---|---|
| **What diagnostic tests were performed to identify the cause of your symptoms? (Select all that apply) - h. Other, please specify - Text** | **Were any of these tests positive for cancer?** | **If yes, which test(s) indicated cancer? (Select all that apply) - Selected Choice** | **If yes, which type of cancer were you told that you had? - Selected Choice** | **Were you treated for the misdiagnosed cancer? - Selected Choice** | **At the time of your misdiagnosis, did you have a mammogram as part of your advanced or metastatic ILC diagnostic process? (if you had ILC in the past, this question refers to the recent misdiagnosis period, not the initial diagnosis)** |
| | 1 | 1,3,4 | 13 | 2 | 1 |
| Ct guided biopsy | 1 | 4,7 | 13 | 1 | 2 |
| | 1 | 5,6 | | | 1 |
| | 1 | 5,6 | | 2 | 2 |
| | 1 | 3,6 | 13 | 1 | 2 |
| | 1 | 3 | 13 | 2 | 1 |
| No tests | 2 | | | 2 | 2 |

| Q26_7_TEXT | Q27 | Q28 | Q29 | Q30 | Q31 |
|---|---|---|---|---|---|
| What diagnostic tests were performed to identify the cause of your symptoms? (Select all that apply) - h. Other, please specify - Text | Were any of these tests positive for cancer? | If yes, which test(s) indicated cancer? (Select all that apply) - Selected Choice | If yes, which type of cancer were you told that you had? - Selected Choice | Were you treated for the misdiagnosed cancer? - Selected Choice | At the time of your misdiagnosis, did you have a mammogram as part of your advanced or metastatic ILC diagnostic process? (if you had ILC in the past, this question refers to the recent misdiagnosis period, not the initial diagnosis) |
| | 1 | 4,5,6 | 13 | 2 | 2 |
| | 1 | 3 | 13 | 1 | 1 |
| | 2 | | | 2 | 1 |
| | | | | | 2 |
| | 1 | 6 | 13 | 2 | |
| | | | | | 2 |

| Q26_7_TEXT | Q27 | Q28 | Q29 | Q30 | Q31 |
|---|---|---|---|---|---|
| What diagnostic tests were performed to identify the cause of your symptoms? (Select all that apply) - h. Other, please specify - Text | Were any of these tests positive for cancer? | If yes, which test(s) indicated cancer? (Select all that apply) - Selected Choice | If yes, which type of cancer were you told that you had? - Selected Choice | Were you treated for the misdiagnosed cancer? - Selected Choice | At the time of your misdiagnosis, did you have a mammogram as part of your advanced or metastatic ILC diagnostic process? (if you had ILC in the past, this question refers to the recent misdiagnosis period, not the initial diagnosis) |
| | 1 | 3 | | 2 | 2 |
| Xray | 2 | | | 2 | 2 |
| | 1 | 4 | | 2 | |
| | 1 | 4,6 | 13 | | 1 |
| | 1 | 2,3,4 | 13 | 2 | |
| bone scan | 1 | 4,7 | 13 | 2 | 2 |
| | 1 | 1,3,4 | 13 | 1 | 2 |

| Q26_7_TEXT | Q27 | Q28 | Q29 | Q30 | Q31 |
|---|---|---|---|---|---|
| What diagnostic tests were performed to identify the cause of your symptoms? (Select all that apply) - h. Other, please specify - Text | Were any of these tests positive for cancer? | If yes, which test(s) indicated cancer? (Select all that apply) - Selected Choice | If yes, which type of cancer were you told that you had? - Selected Choice | Were you treated for the misdiagnosed cancer? - Selected Choice | At the time of your misdiagnosis, did you have a mammogram as part of your advanced or metastatic ILC diagnostic process? (if you had ILC in the past, this question refers to the recent misdiagnosis period, not the initial diagnosis) |
| | | 4,7 | | | 1 |
| | 1 | 3 | 3 | 2 | 2 |
| | | | | | 1 |
| | 1 | 3,6 | | | |
| | 1 | 3,4,6 | | 2 | 1 |
| | 1 | 3,6 | 13 | 2 | 2 |
| | 1 | 4 | 13 | 1 | 2 |

| Q26_7_TEXT | Q27 | Q28 | Q29 | Q30 | Q31 |
|---|---|---|---|---|---|
| What diagnostic tests were performed to identify the cause of your symptoms? (Select all that apply) - h. Other, please specify - Text | Were any of these tests positive for cancer? | If yes, which test(s) indicated cancer? (Select all that apply) - Selected Choice | If yes, which type of cancer were you told that you had? - Selected Choice | Were you treated for the misdiagnosed cancer? - Selected Choice | At the time of your misdiagnosis, did you have a mammogram as part of your advanced or metastatic ILC diagnostic process? (if you had ILC in the past, this question refers to the recent misdiagnosis period, not the initial diagnosis) |
| | 1 | 3,6,7 | 13 | 2 | |
| | 2 | | | 2 | 1 |
| | 2 | | | 2 | 2 |

| Q26_7_TEXT | Q27 | Q28 | Q29 | Q30 | Q31 |
| --- | --- | --- | --- | --- | --- |
| What diagnostic tests were performed to identify the cause of your symptoms? (Select all that apply) - h. Other, please specify - Text | Were any of these tests positive for cancer? | If yes, which test(s) indicated cancer? (Select all that apply) - Selected Choice | If yes, which type of cancer were you told that you had? - Selected Choice | Were you treated for the misdiagnosed cancer? - Selected Choice | At the time of your misdiagnosis, did you have a mammogram as part of your advanced or metastatic ILC diagnostic process? (if you had ILC in the past, this question refers to the recent misdiagnosis period, not the initial diagnosis) |
|  | 1 | 1 | 13 | 2 | 2 |
| None | 2 |  |  |  | 2 |
|  | 2 |  |  |  |  |
| Mammogram | 1 | 3 | 13 |  |  |
|  | 1 |  |  |  |  |
|  |  | 7 |  |  |  |

| Q26_7_TEXT | Q27 | Q28 | Q29 | Q30 | Q31 |
|---|---|---|---|---|---|
| What diagnostic tests were performed to identify the cause of your symptoms? (Select all that apply) - h. Other, please specify - Text | Were any of these tests positive for cancer? | If yes, which test(s) indicated cancer? (Select all that apply) - Selected Choice | If yes, which type of cancer were you told that you had? - Selected Choice | Were you treated for the misdiagnosed cancer? - Selected Choice | At the time of your misdiagnosis, did you have a mammogram as part of your advanced or metastatic ILC diagnostic process? (if you had ILC in the past, this question refers to the recent misdiagnosis period, not the initial diagnosis) |
| | 1 | 3,4 | 13 | 2 | |
| Nuclear bone scan | 1 | 4,7 | 13 | 2 | 2 |
| | 1 | 2,3 | | | |
| Bone scan | 1 | 7 | | | 2 |
| | 1 | 4 | 13 | | |

| Q26_7_TEXT | Q27 | Q28 | Q29 | Q30 | Q31 |
|---|---|---|---|---|---|
| What diagnostic tests were performed to identify the cause of your symptoms? (Select all that apply) - h. Other, please specify - Text | Were any of these tests positive for cancer? | If yes, which test(s) indicated cancer? (Select all that apply) - Selected Choice | If yes, which type of cancer were you told that you had? - Selected Choice | Were you treated for the misdiagnosed cancer? - Selected Choice | At the time of your misdiagnosis, did you have a mammogram as part of your advanced or metastatic ILC diagnostic process? (if you had ILC in the past, this question refers to the recent misdiagnosis period, not the initial diagnosis) |
| | 1 | 1,3,6 | 3 | 2 | 1 |
| | | 6 | | | |
| | 1 | 6 | 13 | 2 | |

| Q26_7_TEXT | Q27 | Q28 | Q29 | Q30 | Q31 |
|---|---|---|---|---|---|
| What diagnostic tests were performed to identify the cause of your symptoms? (Select all that apply) - h. Other, please specify - Text | Were any of these tests positive for cancer? | If yes, which test(s) indicated cancer? (Select all that apply) - Selected Choice | If yes, which type of cancer were you told that you had? - Selected Choice | Were you treated for the misdiagnosed cancer? - Selected Choice | At the time of your misdiagnosis, did you have a mammogram as part of your advanced or metastatic ILC diagnostic process? (if you had ILC in the past, this question refers to the recent misdiagnosis period, not the initial diagnosis) |
| | 2 | | | | |
| | 1 | 5,6 | | 2 | 2 |
| | 1 | 3,4 | 13 | | |

| Q26_7_TEXT | Q27 | Q28 | Q29 | Q30 | Q31 |
|---|---|---|---|---|---|
| What diagnostic tests were performed to identify the cause of your symptoms? (Select all that apply) - h. Other, please specify - Text | Were any of these tests positive for cancer? | If yes, which test(s) indicated cancer? (Select all that apply) - Selected Choice | If yes, which type of cancer were you told that you had? - Selected Choice | Were you treated for the misdiagnosed cancer? - Selected Choice | At the time of your misdiagnosis, did you have a mammogram as part of your advanced or metastatic ILC diagnostic process? (if you had ILC in the past, this question refers to the recent misdiagnosis period, not the initial diagnosis) |
| | 1 | 6 | | 2 | 1 |
| | | | | 2 | 1 |
| | | | | 2 | |
| | 1 | 3,7 | 13 | 1 | 2 |
| | 2 | | | 2 | 2 |

| Q26_7_TEXT | Q27 | Q28 | Q29 | Q30 | Q31 |
|---|---|---|---|---|---|
| What diagnostic tests were performed to identify the cause of your symptoms? (Select all that apply) - h. Other, please specify - Text | Were any of these tests positive for cancer? | If yes, which test(s) indicated cancer? (Select all that apply) - Selected Choice | If yes, which type of cancer were you told that you had? - Selected Choice | Were you treated for the misdiagnosed cancer? - Selected Choice | At the time of your misdiagnosis, did you have a mammogram as part of your advanced or metastatic ILC diagnostic process? (if you had ILC in the past, this question refers to the recent misdiagnosis period, not the initial diagnosis) |
| | | | | 2 | |
| Xray, mammogram, | 2 | | | | 1 |
| | 1 | 1,3 | 13 | | 1 |
| | 1 | 1 | 13 | | |
| | 1 | 3,4 | 13 | 1 | 1 |
| | 1 | 4,6 | 13 | 1 | 2 |

| Q26_7_TEXT | Q27 | Q28 | Q29 | Q30 | Q31 |
|---|---|---|---|---|---|
| What diagnostic tests were performed to identify the cause of your symptoms? (Select all that apply) - h. Other, please specify - Text | Were any of these tests positive for cancer? | If yes, which test(s) indicated cancer? (Select all that apply) - Selected Choice | If yes, which type of cancer were you told that you had? - Selected Choice | Were you treated for the misdiagnosed cancer? - Selected Choice | At the time of your misdiagnosis, did you have a mammogram as part of your advanced or metastatic ILC diagnostic process? (if you had ILC in the past, this question refers to the recent misdiagnosis period, not the initial diagnosis) |
| | 1 | 3 | | 1 | 1 |
| | 1 | 5,6 | 13 | 1 | 2 |
| | 1 | 3,4 | 13 | 2 | 1 |
| Xrays | 1 | 6 | 13 | | 2 |

| Q26_7_TEXT | Q27 | Q28 | Q29 | Q30 | Q31 |
|---|---|---|---|---|---|
| What diagnostic tests were performed to identify the cause of your symptoms? (Select all that apply) - h. Other, please specify - Text | Were any of these tests positive for cancer? | If yes, which test(s) indicated cancer? (Select all that apply) - Selected Choice | If yes, which type of cancer were you told that you had? - Selected Choice | Were you treated for the misdiagnosed cancer? - Selected Choice | At the time of your misdiagnosis, did you have a mammogram as part of your advanced or metastatic ILC diagnostic process? (if you had ILC in the past, this question refers to the recent misdiagnosis period, not the initial diagnosis) |
| | 2 | 3 | 13 | 1 | 1 |
| office physical exa | 1 | 6 | 13 | 2 | 2 |
| | 2 | 3,4,5,6 | | 2 | |
| | 1 | 3 | 13 | 1 | 2 |
| | 1 | 6 | | 2 | 2 |
| | 1 | 3,4,6 | | | |
| Bone scan | 1 | 3,4,6 | | 2 | 2 |

| Q26_7_TEXT | Q27 | Q28 | Q29 | Q30 | Q31 |
|---|---|---|---|---|---|
| What diagnostic tests were performed to identify the cause of your symptoms? (Select all that apply) - h. Other, please specify - Text | Were any of these tests positive for cancer? | If yes, which test(s) indicated cancer? (Select all that apply) - Selected Choice | If yes, which type of cancer were you told that you had? - Selected Choice | Were you treated for the misdiagnosed cancer? - Selected Choice | At the time of your misdiagnosis, did you have a mammogram as part of your advanced or metastatic ILC diagnostic process? (if you had ILC in the past, this question refers to the recent misdiagnosis period, not the initial diagnosis) |
| | 1 | 3 | 13 | 2 | 2 |
| PET scan | 1 | 3,4,5,6,7 | 13 | 2 | 1 |
| | 1 | 3,4,5,6 | 13 | 2 | |
| | | 3 | | 2 | |
| | 1 | 3 | 4 | 1 | 2 |

| Q26_7_TEXT | Q27 | Q28 | Q29 | Q30 | Q31 |
|---|---|---|---|---|---|
| What diagnostic tests were performed to identify the cause of your symptoms? (Select all that apply) - h. Other, please specify - Text | Were any of these tests positive for cancer? | If yes, which test(s) indicated cancer? (Select all that apply) - Selected Choice | If yes, which type of cancer were you told that you had? - Selected Choice | Were you treated for the misdiagnosed cancer? - Selected Choice | At the time of your misdiagnosis, did you have a mammogram as part of your advanced or metastatic ILC diagnostic process? (if you had ILC in the past, this question refers to the recent misdiagnosis period, not the initial diagnosis) |
| | 1 | 4 | 13 | 1 | 1 |
| | 1 | 6 | 13 | 2 | 1 |
| | 1 | 5 | 12 | 2 | 2 |
| PET | 1 | 3,4,5,6 | 13 | 2 | 1 |
| | 1 | 4,6 | 13 | | |
| | 1 | 6 | | 2 | 1 |
| | 1 | 3,5,6 | 13 | 2 | 2 |
| | | | | | 2 |

| Q26_7_TEXT | Q27 | Q28 | Q29 | Q30 | Q31 |
|---|---|---|---|---|---|
| What diagnostic tests were performed to identify the cause of your symptoms? (Select all that apply) - h. Other, please specify - Text | Were any of these tests positive for cancer? | If yes, which test(s) indicated cancer? (Select all that apply) - Selected Choice | If yes, which type of cancer were you told that you had? - Selected Choice | Were you treated for the misdiagnosed cancer? - Selected Choice | At the time of your misdiagnosis, did you have a mammogram as part of your advanced or metastatic ILC diagnostic process? (if you had ILC in the past, this question refers to the recent misdiagnosis period, not the initial diagnosis) |
| PET scan | 1 | 4,5,7 | 13 | 2 | 2 |
| | 1 | 3,4,6 | 13 | 1 | 2 |
| A chat | 2 | | | | 1 |
| Fes pet scan | 1 | 3,4,5,6 | | | |

| Q26_7_TEXT | Q27 | Q28 | Q29 | Q30 | Q31 |
|---|---|---|---|---|---|
| What diagnostic tests were performed to identify the cause of your symptoms? (Select all that apply) - h. Other, please specify - Text | Were any of these tests positive for cancer? | If yes, which test(s) indicated cancer? (Select all that apply) - Selected Choice | If yes, which type of cancer were you told that you had? - Selected Choice | Were you treated for the misdiagnosed cancer? - Selected Choice | At the time of your misdiagnosis, did you have a mammogram as part of your advanced or metastatic ILC diagnostic process? (if you had ILC in the past, this question refers to the recent misdiagnosis period, not the initial diagnosis) |
| | 1 | 3 | 13 | 1 | 1 |
| | 1 | 4,6 | 3 | 2 | 2 |
| bone biopsy | 1 | 3,5,7 | 13 | 1 | 2 |
| | 1 | 3,4,6,7 | 13 | 2 | 2 |
| | | | | 2 | |
| | 2 | | | 2 | |
| | 1 | 7 | 13 | 2 | 1 |

| Q26_7_TEXT | Q27 | Q28 | Q29 | Q30 | Q31 |
|---|---|---|---|---|---|
| What diagnostic tests were performed to identify the cause of your symptoms? (Select all that apply) - h. Other, please specify - Text | Were any of these tests positive for cancer? | If yes, which test(s) indicated cancer? (Select all that apply) - Selected Choice | If yes, which type of cancer were you told that you had? - Selected Choice | Were you treated for the misdiagnosed cancer? - Selected Choice | At the time of your misdiagnosis, did you have a mammogram as part of your advanced or metastatic ILC diagnostic process? (if you had ILC in the past, this question refers to the recent misdiagnosis period, not the initial diagnosis) |
| | 1 | 3,4,6 | 13 | 2 | |
| | | | 13 | 1 | |
| | 2 | | | | 2 |

| Q26_7_TEXT | Q27 | Q28 | Q29 | Q30 | Q31 |
| --- | --- | --- | --- | --- | --- |
| What diagnostic tests were performed to identify the cause of your symptoms? (Select all that apply) - h. Other, please specify - Text | Were any of these tests positive for cancer? | If yes, which test(s) indicated cancer? (Select all that apply) - Selected Choice | If yes, which type of cancer were you told that you had? - Selected Choice | Were you treated for the misdiagnosed cancer? - Selected Choice | At the time of your misdiagnosis, did you have a mammogram as part of your advanced or metastatic ILC diagnostic process? (if you had ILC in the past, this question refers to the recent misdiagnosis period, not the initial diagnosis) |
| | | | | 2 | |
| | 1 | 6 | | 2 | 2 |
| Bone scan | 1 | 3,4,5,6,7 | 13 | 2 | 1 |
| None | 2 | | | | 2 |
| | 1 | 3,4 | 13 | 2 | |
| | 1 | 4 | 13 | 1 | 1 |
| | 1 | 3 | 13 | 1 | 2 |

| Q26_7_TEXT | Q27 | Q28 | Q29 | Q30 | Q31 |
|---|---|---|---|---|---|
| What diagnostic tests were performed to identify the cause of your symptoms? (Select all that apply) - h. Other, please specify - Text | Were any of these tests positive for cancer? | If yes, which test(s) indicated cancer? (Select all that apply) - Selected Choice | If yes, which type of cancer were you told that you had? - Selected Choice | Were you treated for the misdiagnosed cancer? - Selected Choice | At the time of your misdiagnosis, did you have a mammogram as part of your advanced or metastatic ILC diagnostic process? (if you had ILC in the past, this question refers to the recent misdiagnosis period, not the initial diagnosis) |
| | 1 | 6 | 13 | 2 | 2 |
| None | 2 | | 13 | 2 | 2 |
| a - colonoscopy x2. | 1 | 2,3,4,6 | 13 | 2 | 2 |
| | 1 | 3 | 13 | 1 | 2 |
| v to know if Stage III | 1 | 3 | | | 1 |
| | 1 | 6 | 13 | 1 | |

| Q26_7_TEXT | Q27 | Q28 | Q29 | Q30 | Q31 |
|---|---|---|---|---|---|
| What diagnostic tests were performed to identify the cause of your symptoms? (Select all that apply) - h. Other, please specify - Text | Were any of these tests positive for cancer? | If yes, which test(s) indicated cancer? (Select all that apply) - Selected Choice | If yes, which type of cancer were you told that you had? - Selected Choice | Were you treated for the misdiagnosed cancer? - Selected Choice | At the time of your misdiagnosis, did you have a mammogram as part of your advanced or metastatic ILC diagnostic process? (if you had ILC in the past, this question refers to the recent misdiagnosis period, not the initial diagnosis) |
| Pet | 1 | 3 | 13 | 2 | 1 |
| | 2 | 4,6,7 | | 2 | 2 |
| X ray | 2 | | | 2 | |

| Q26_7_TEXT | Q27 | Q28 | Q29 | Q30 | Q31 |
| --- | --- | --- | --- | --- | --- |
| What diagnostic tests were performed to identify the cause of your symptoms? (Select all that apply) - h. Other, please specify - Text | Were any of these tests positive for cancer? | If yes, which test(s) indicated cancer? (Select all that apply) - Selected Choice | If yes, which type of cancer were you told that you had? - Selected Choice | Were you treated for the misdiagnosed cancer? - Selected Choice | At the time of your misdiagnosis, did you have a mammogram as part of your advanced or metastatic ILC diagnostic process? (if you had ILC in the past, this question refers to the recent misdiagnosis period, not the initial diagnosis) |
| | 1 | 1,2,3,4,5 | | 2 | 2 |
| MRI re hip pain | 1 | 6 | 13 | | 2 |
| | 1 | 4 | 13 | 2 | 2 |
| | 1 | 3 | 13 | 1 | 1 |
| | 2 | 7 | | | 2 |
| | 2 | | | 2 | 2 |
| | | | | | 2 |

| Q26_7_TEXT | Q27 | Q28 | Q29 | Q30 | Q31 |
|---|---|---|---|---|---|
| What diagnostic tests were performed to identify the cause of your symptoms? (Select all that apply) - h. Other, please specify - Text | Were any of these tests positive for cancer? | If yes, which test(s) indicated cancer? (Select all that apply) - Selected Choice | If yes, which type of cancer were you told that you had? - Selected Choice | Were you treated for the misdiagnosed cancer? - Selected Choice | At the time of your misdiagnosis, did you have a mammogram as part of your advanced or metastatic ILC diagnostic process? (if you had ILC in the past, this question refers to the recent misdiagnosis period, not the initial diagnosis) |
| | 1 | 3 | 13 | | |
| | 1 | 3,4 | 13 | 2 | 1 |
| | 1 | 3 | | 2 | 1 |
| | | | | | 2 |
| | 1 | 6 | 13 | | 1 |

| Q26_7_TEXT | Q27 | Q28 | Q29 | Q30 | Q31 |
|---|---|---|---|---|---|
| What diagnostic tests were performed to identify the cause of your symptoms? (Select all that apply) - h. Other, please specify - Text | Were any of these tests positive for cancer? | If yes, which test(s) indicated cancer? (Select all that apply) - Selected Choice | If yes, which type of cancer were you told that you had? - Selected Choice | Were you treated for the misdiagnosed cancer? - Selected Choice | At the time of your misdiagnosis, did you have a mammogram as part of your advanced or metastatic ILC diagnostic process? (if you had ILC in the past, this question refers to the recent misdiagnosis period, not the initial diagnosis) |
| scription of symptor | 2 | | | 2 | 2 |
| | 1 | 3,5,6 | 13 | 2 | 1 |
| | 2 | | | 2 | |
| | 1 | 3,4,6 | 13 | 2 | |
| | 2 | | | | 1 |
| | 1 | 3 | 13 | 1 | 1 |
| Pet/ct | 1 | 4,5 | 3 | 1 | 2 |

| Q26_7_TEXT | Q27 | Q28 | Q29 | Q30 | Q31 |
|---|---|---|---|---|---|
| What diagnostic tests were performed to identify the cause of your symptoms? (Select all that apply) - h. Other, please specify - Text | Were any of these tests positive for cancer? | If yes, which test(s) indicated cancer? (Select all that apply) - Selected Choice | If yes, which type of cancer were you told that you had? - Selected Choice | Were you treated for the misdiagnosed cancer? - Selected Choice | At the time of your misdiagnosis, did you have a mammogram as part of your advanced or metastatic ILC diagnostic process? (if you had ILC in the past, this question refers to the recent misdiagnosis period, not the initial diagnosis) |
| | 2 | | | 2 | 2 |
| ed in April 2025 was | 1 | 3,5 | 13 | 2 | 1 |
| | 1 | 1,3,7 | 3 | 2 | 2 |
| | 2 | | | | 2 |
| N/A | 2 | | | | |

| Q26_7_TEXT | Q27 | Q28 | Q29 | Q30 | Q31 |
| --- | --- | --- | --- | --- | --- |
| What diagnostic tests were performed to identify the cause of your symptoms? (Select all that apply) - h. Other, please specify - Text | Were any of these tests positive for cancer? | If yes, which test(s) indicated cancer? (Select all that apply) - Selected Choice | If yes, which type of cancer were you told that you had? - Selected Choice | Were you treated for the misdiagnosed cancer? - Selected Choice | At the time of your misdiagnosis, did you have a mammogram as part of your advanced or metastatic ILC diagnostic process? (if you had ILC in the past, this question refers to the recent misdiagnosis period, not the initial diagnosis) |
| FES PET scan | 1 | 5,6,7 | 13 | 2 | |
| | 2 | 7 | 13 | 2 | 1 |

| Q26_7_TEXT | Q27 | Q28 | Q29 | Q30 | Q31 |
|---|---|---|---|---|---|
| What diagnostic tests were performed to identify the cause of your symptoms? (Select all that apply) - h. Other, please specify - Text | Were any of these tests positive for cancer? | If yes, which test(s) indicated cancer? (Select all that apply) - Selected Choice | If yes, which type of cancer were you told that you had? - Selected Choice | Were you treated for the misdiagnosed cancer? - Selected Choice | At the time of your misdiagnosis, did you have a mammogram as part of your advanced or metastatic ILC diagnostic process? (if you had ILC in the past, this question refers to the recent misdiagnosis period, not the initial diagnosis) |
| Xray | 1 | 6 | 13 | 2 | 2 |
| | 1 | 1,2,3 | 13 | 2 | |
| PET SCAN | 1 | 3 | 13 | | |
| | 2 | | | | |
| | 2 | | | 2 | |

| Q26_7_TEXT | Q27 | Q28 | Q29 | Q30 | Q31 |
|---|---|---|---|---|---|
| What diagnostic tests were performed to identify the cause of your symptoms? (Select all that apply) - h. Other, please specify - Text | Were any of these tests positive for cancer? | If yes, which test(s) indicated cancer? (Select all that apply) - Selected Choice | If yes, which type of cancer were you told that you had? - Selected Choice | Were you treated for the misdiagnosed cancer? - Selected Choice | At the time of your misdiagnosis, did you have a mammogram as part of your advanced or metastatic ILC diagnostic process? (if you had ILC in the past, this question refers to the recent misdiagnosis period, not the initial diagnosis) |
| | 1 | 4,6 | | | |
| | 2 | | | | |

| Q26_7_TEXT | Q27 | Q28 | Q29 | Q30 | Q31 |
| --- | --- | --- | --- | --- | --- |
| What diagnostic tests were performed to identify the cause of your symptoms? (Select all that apply) - h. Other, please specify - Text | Were any of these tests positive for cancer? | If yes, which test(s) indicated cancer? (Select all that apply) - Selected Choice | If yes, which type of cancer were you told that you had? - Selected Choice | Were you treated for the misdiagnosed cancer? - Selected Choice | At the time of your misdiagnosis, did you have a mammogram as part of your advanced or metastatic ILC diagnostic process? (if you had ILC in the past, this question refers to the recent misdiagnosis period, not the initial diagnosis) |
| | 2 | | | 2 | |
| | 2 | | | 2 | |

| Q26_7_TEXT | Q27 | Q28 | Q29 | Q30 | Q31 |
|---|---|---|---|---|---|
| What diagnostic tests were performed to identify the cause of your symptoms? (Select all that apply) - h. Other, please specify - Text | Were any of these tests positive for cancer? | If yes, which test(s) indicated cancer? (Select all that apply) - Selected Choice | If yes, which type of cancer were you told that you had? - Selected Choice | Were you treated for the misdiagnosed cancer? - Selected Choice | At the time of your misdiagnosis, did you have a mammogram as part of your advanced or metastatic ILC diagnostic process? (if you had ILC in the past, this question refers to the recent misdiagnosis period, not the initial diagnosis) |
| None | | | | | |
| | 1 | 3,4 | 13 | 2 | 2 |

| Q26_7_TEXT | Q27 | Q28 | Q29 | Q30 | Q31 |
|---|---|---|---|---|---|
| What diagnostic tests were performed to identify the cause of your symptoms? (Select all that apply) - h. Other, please specify - Text | Were any of these tests positive for cancer? | If yes, which test(s) indicated cancer? (Select all that apply) - Selected Choice | If yes, which type of cancer were you told that you had? - Selected Choice | Were you treated for the misdiagnosed cancer? - Selected Choice | At the time of your misdiagnosis, did you have a mammogram as part of your advanced or metastatic ILC diagnostic process? (if you had ILC in the past, this question refers to the recent misdiagnosis period, not the initial diagnosis) |
| PET scan | 1 | 3,5,6 | 13 | | 2 |
| | | | | | 1 |
| Physical exam | 2 | | | 1 | 2 |
| | 1 | 3 | | 2 | 2 |
| Pet scan | 1 | 4,5 | 13 | 2 | |

| Q26_7_TEXT | Q27 | Q28 | Q29 | Q30 | Q31 |
|---|---|---|---|---|---|
| What diagnostic tests were performed to identify the cause of your symptoms? (Select all that apply) - h. Other, please specify - Text | Were any of these tests positive for cancer? | If yes, which test(s) indicated cancer? (Select all that apply) - Selected Choice | If yes, which type of cancer were you told that you had? - Selected Choice | Were you treated for the misdiagnosed cancer? - Selected Choice | At the time of your misdiagnosis, did you have a mammogram as part of your advanced or metastatic ILC diagnostic process? (if you had ILC in the past, this question refers to the recent misdiagnosis period, not the initial diagnosis) |
| Cystoscopy, PET | 1 | 3,4,5,7 | 13 | 2 | |
| | 1 | 3,6 | 13 | 2 | 2 |

| Q26_7_TEXT | Q27 | Q28 | Q29 | Q30 | Q31 |
| --- | --- | --- | --- | --- | --- |
| What diagnostic tests were performed to identify the cause of your symptoms? (Select all that apply) - h. Other, please specify - Text | Were any of these tests positive for cancer? | If yes, which test(s) indicated cancer? (Select all that apply) - Selected Choice | If yes, which type of cancer were you told that you had? - Selected Choice | Were you treated for the misdiagnosed cancer? - Selected Choice | At the time of your misdiagnosis, did you have a mammogram as part of your advanced or metastatic ILC diagnostic process? (if you had ILC in the past, this question refers to the recent misdiagnosis period, not the initial diagnosis) |
| PET CT | 2 | | 13 | | 2 |
| | 1 | 3 | 13 | 1 | 1 |

| Q26_7_TEXT | Q27 | Q28 | Q29 | Q30 | Q31 |
|---|---|---|---|---|---|
| What diagnostic tests were performed to identify the cause of your symptoms? (Select all that apply) - h. Other, please specify - Text | Were any of these tests positive for cancer? | If yes, which test(s) indicated cancer? (Select all that apply) - Selected Choice | If yes, which type of cancer were you told that you had? - Selected Choice | Were you treated for the misdiagnosed cancer? - Selected Choice | At the time of your misdiagnosis, did you have a mammogram as part of your advanced or metastatic ILC diagnostic process? (if you had ILC in the past, this question refers to the recent misdiagnosis period, not the initial diagnosis) |
| | 1 | 3,4,5,6 | 13 | 1 | 1 |
| | 2 | | | 2 | 1 |

| Q26_7_TEXT | Q27 | Q28 | Q29 | Q30 | Q31 |
|---|---|---|---|---|---|
| What diagnostic tests were performed to identify the cause of your symptoms? (Select all that apply) - h. Other, please specify - Text | Were any of these tests positive for cancer? | If yes, which test(s) indicated cancer? (Select all that apply) - Selected Choice | If yes, which type of cancer were you told that you had? - Selected Choice | Were you treated for the misdiagnosed cancer? - Selected Choice | At the time of your misdiagnosis, did you have a mammogram as part of your advanced or metastatic ILC diagnostic process? (if you had ILC in the past, this question refers to the recent misdiagnosis period, not the initial diagnosis) |
| iopsies from surger | 1 | 7 | 9 | 2 | 2 |
| N/A | 2 | 4 | 13 | 2 | 2 |
| Mammogram | 1 | 3 | 3 | 1 | 2 |

| Q26_7_TEXT | Q27 | Q28 | Q29 | Q30 | Q31 |
|---|---|---|---|---|---|
| What diagnostic tests were performed to identify the cause of your symptoms? (Select all that apply) - h. Other, please specify - Text | Were any of these tests positive for cancer? | If yes, which test(s) indicated cancer? (Select all that apply) - Selected Choice | If yes, which type of cancer were you told that you had? - Selected Choice | Were you treated for the misdiagnosed cancer? - Selected Choice | At the time of your misdiagnosis, did you have a mammogram as part of your advanced or metastatic ILC diagnostic process? (if you had ILC in the past, this question refers to the recent misdiagnosis period, not the initial diagnosis) |
| PET scan | 1 | 3,7 | 13 | 1<br>2 | 2 |

| Q26_7_TEXT | Q27 | Q28 | Q29 | Q30 | Q31 |
|---|---|---|---|---|---|
| What diagnostic tests were performed to identify the cause of your symptoms? (Select all that apply) - h. Other, please specify - Text | Were any of these tests positive for cancer? | If yes, which test(s) indicated cancer? (Select all that apply) - Selected Choice | If yes, which type of cancer were you told that you had? - Selected Choice | Were you treated for the misdiagnosed cancer? - Selected Choice | At the time of your misdiagnosis, did you have a mammogram as part of your advanced or metastatic ILC diagnostic process? (if you had ILC in the past, this question refers to the recent misdiagnosis period, not the initial diagnosis) |
| ng loss in seemed ur | 1 | 5,6 | 13 | 2 | 1 |
| | | | | 2 | |
| need them and was experiencing normal perimenopausal sym | | | | 2 | 2 |
| PET/CT scan | 1 | 3,4,5,6,7 | 13 | | |
| | 1 | 4,6 | 13 | 1 | 2 |

| Q26_7_TEXT | Q27 | Q28 | Q29 | Q30 | Q31 |
|---|---|---|---|---|---|
| What diagnostic tests were performed to identify the cause of your symptoms? (Select all that apply) - h. Other, please specify - Text | Were any of these tests positive for cancer? | If yes, which test(s) indicated cancer? (Select all that apply) - Selected Choice | If yes, which type of cancer were you told that you had? - Selected Choice | Were you treated for the misdiagnosed cancer? - Selected Choice | At the time of your misdiagnosis, did you have a mammogram as part of your advanced or metastatic ILC diagnostic process? (if you had ILC in the past, this question refers to the recent misdiagnosis period, not the initial diagnosis) |
| | 1 | 3 | 13 | | 2 |
| | 1 | 4,6 | | | 1 |
| Audiometry | 2 | | | | 2 |

| Q32 | Q33 | Q34 | Q35 | Q36 | Q37 |
| --- | --- | --- | --- | --- | --- |
| If yes, how long after your symptoms did you have your first mammogram? | Was the mammogram able to detect the ILC? | Did you undergo additional testing? (Select all that apply) - Selected Choice | How long after your initial mammogram did you undergo these additional tests? | Were you undergoing active surveillance for breast cancer at the time of your metastatic ILC diagnosis? | If yes, what type of surveillance? (Select all that apply) - Selected Choice |
| 2 | 2 | 1,2,3 | 1 | 1 | |
| | 3 | 4 | 5 | 2 | 8 |
| 1 | 2 | 2,4 | 2 | 2 | 3 |
| 4 | 2 | 1 | 3 | 1 | |
| 2 | 2 | 1,2,4 | 5 | 2 | 7 |

| Q32 | Q33 | Q34 | Q35 | Q36 | Q37 |
|---|---|---|---|---|---|
| **If yes, how long after your symptoms did you have your first mammogram?** | **Was the mammogram able to detect the ILC?** | **Did you undergo additional testing? (Select all that apply) - Selected Choice** | **How long after your initial mammogram did you undergo these additional tests?** | **Were you undergoing active surveillance for breast cancer at the time of your metastatic ILC diagnosis?** | **If yes, what type of surveillance? (Select all that apply) - Selected Choice** |
| 2 | 1 | 1,2,4 | 2 | 1 | |
| 5 | 3 | | | 2 | 3 |
| 1 | 2 | 6 | | 2 | 2,3 |
| 5 | 2 | 6 | 6 | 2 | 3 |
| | | | | 1 | |
| 2 | 2 | 1,2,4 | 1 | 1 | |
| | | 6 | | 1 | |
| | | | | 2 | 3 |

| Q32 | Q33 | Q34 | Q35 | Q36 | Q37 |
| --- | --- | --- | --- | --- | --- |
| If yes, how long after your symptoms did you have your first mammogram? | Was the mammogram able to detect the ILC? | Did you undergo additional testing? (Select all that apply) - Selected Choice | How long after your initial mammogram did you undergo these additional tests? | Were you undergoing active surveillance for breast cancer at the time of your metastatic ILC diagnosis? | If yes, what type of surveillance? (Select all that apply) - Selected Choice |
| | 3 | 4 | 6 | 1 | |
| 1 | 2 | 1,4 | 1 | 2 | 3 |
| 3 | 2 | 1 | 1 | 1 | |
| | | 4 | 1 | 1 | |
| | | 4 | | 2 | 8 |
| | 1 | 1,4 | 1 | 2 | 1,3,6 |

| Q32 | Q33 | Q34 | Q35 | Q36 | Q37 |
|---|---|---|---|---|---|
| If yes, how long after your symptoms did you have your first mammogram? | Was the mammogram able to detect the ILC? | Did you undergo additional testing? (Select all that apply) - Selected Choice | How long after your initial mammogram did you undergo these additional tests? | Were you undergoing active surveillance for breast cancer at the time of your metastatic ILC diagnosis? | If yes, what type of surveillance? (Select all that apply) - Selected Choice |
| | 1 | 1,2,4 | 1 | 1 | |
| | | | | 1 | |
| | | | | 2 | 3 |
| 1 | 2 | 1 | 5 | 1 | |
| 1 | 2 | 1,2,4 | 2 | 2 | |
| | | 1,4 | 1 | 2 | 1,4,8 |
| | | | | 2 | 8 |
| | | | | 1 | |

| Q32 | Q33 | Q34 | Q35 | Q36 | Q37 |
|---|---|---|---|---|---|
| If yes, how long after your symptoms did you have your first mammogram? | Was the mammogram able to detect the ILC? | Did you undergo additional testing? (Select all that apply) - Selected Choice | How long after your initial mammogram did you undergo these additional tests? | Were you undergoing active surveillance for breast cancer at the time of your metastatic ILC diagnosis? | If yes, what type of surveillance? (Select all that apply) - Selected Choice |
| 1 | 2 | 1 | 1 | 2 | 3 |
| | | | | 2 | 5 |
| 1 | 2 | 1,2,4 | 1 | 1 | |
| | | | 1 | | |
| 2 | 2 | 1,2,3,4 | 2 | 2 | 3 |
| 1 | 1 | 1,2 | 2 | 1 | |
| | 3 | 4 | | 1 | |

| Q32 | Q33 | Q34 | Q35 | Q36 | Q37 |
| --- | --- | --- | --- | --- | --- |
| If yes, how long after your symptoms did you have your first mammogram? | Was the mammogram able to detect the ILC? | Did you undergo additional testing? (Select all that apply) - Selected Choice | How long after your initial mammogram did you undergo these additional tests? | Were you undergoing active surveillance for breast cancer at the time of your metastatic ILC diagnosis? | If yes, what type of surveillance? (Select all that apply) - Selected Choice |
| | | 2,4,5 | | 2 | 8 |
| 1 | 2 | 1 | 1 | 1 | |
| | 2 | | | | |
| | 2 | 1,4 | 1 | 2 | 6 |

| Q32 | Q33 | Q34 | Q35 | Q36 | Q37 |
| --- | --- | --- | --- | --- | --- |
| If yes, how long after your symptoms did you have your first mammogram? | Was the mammogram able to detect the ILC? | Did you undergo additional testing? (Select all that apply) - Selected Choice | How long after your initial mammogram did you undergo these additional tests? | Were you undergoing active surveillance for breast cancer at the time of your metastatic ILC diagnosis? | If yes, what type of surveillance? (Select all that apply) - Selected Choice |
| | | | | 2 | 6 |
| | 2 | 4 | | 2 | 5,8 |
| | 3 | 6 | 6 | 1 | |

| Q32 | Q33 | Q34 | Q35 | Q36 | Q37 |
|---|---|---|---|---|---|
| If yes, how long after your symptoms did you have your first mammogram? | Was the mammogram able to detect the ILC? | Did you undergo additional testing? (Select all that apply) - Selected Choice | How long after your initial mammogram did you undergo these additional tests? | Were you undergoing active surveillance for breast cancer at the time of your metastatic ILC diagnosis? | If yes, what type of surveillance? (Select all that apply) - Selected Choice |
| | 3 | 1,4 | 1 | 1 | |

| Q32 | Q33 | Q34 | Q35 | Q36 | Q37 |
|---|---|---|---|---|---|
| If yes, how long after your symptoms did you have your first mammogram? | Was the mammogram able to detect the ILC? | Did you undergo additional testing? (Select all that apply) - Selected Choice | How long after your initial mammogram did you undergo these additional tests? | Were you undergoing active surveillance for breast cancer at the time of your metastatic ILC diagnosis? | If yes, what type of surveillance? (Select all that apply) - Selected Choice |
| 1 | 2 | 1,2,3 | 1 | 1 | |
| | 2 | 1,2,3,4 | 2 | 2 | 1,2,5,6 |

| Q32 | Q33 | Q34 | Q35 | Q36 | Q37 |
|---|---|---|---|---|---|
| If yes, how long after your symptoms did you have your first mammogram? | Was the mammogram able to detect the ILC? | Did you undergo additional testing? (Select all that apply) - Selected Choice | How long after your initial mammogram did you undergo these additional tests? | Were you undergoing active surveillance for breast cancer at the time of your metastatic ILC diagnosis? | If yes, what type of surveillance? (Select all that apply) - Selected Choice |
| | 2 | 1,2,4 | 1 | 2 | 1,2 |
| | 2 | 1,2,3,4 | 2 | 2 | 1,2,3,4,5,6,7 |
| | | | | 2 | 3,5 |
| | 1 | 1,2,3,4 | 1 | 2 | 3 |
| 3 | 1 | 4 | 1 | 1 | |

| Q32 | Q33 | Q34 | Q35 | Q36 | Q37 |
|---|---|---|---|---|---|
| **If yes, how long after your symptoms did you have your first mammogram?** | **Was the mammogram able to detect the ILC?** | **Did you undergo additional testing? (Select all that apply) - Selected Choice** | **How long after your initial mammogram did you undergo these additional tests?** | **Were you undergoing active surveillance for breast cancer at the time of your metastatic ILC diagnosis?** | **If yes, what type of surveillance? (Select all that apply) - Selected Choice** |
| 5 | 2 | 1,2 | 2 | 1 | |
| 5 | 2 | 1,2 | 1 | 1 | |
| | | | | 2 | |
| | 3 | | | 2 | 1 |
| | | | | 2 | 1,2,5,6 |
| | 3 | 1,2,4 | 5 | 2 | 1,5,6 |

| Q32 | Q33 | Q34 | Q35 | Q36 | Q37 |
|---|---|---|---|---|---|
| If yes, how long after your symptoms did you have your first mammogram? | Was the mammogram able to detect the ILC? | Did you undergo additional testing? (Select all that apply) - Selected Choice | How long after your initial mammogram did you undergo these additional tests? | Were you undergoing active surveillance for breast cancer at the time of your metastatic ILC diagnosis? | If yes, what type of surveillance? (Select all that apply) - Selected Choice |
| | | | | 2 | 6 |
| 3 | 2 | 1 | 1 | 2 | 3,4 |
| 2 | 2 | 1,2,4 | 1 | 1 | |
| | 3 | 5 | | 1 | |
| | 2 | 1,3,4,5 | 2 | 1 | |
| | | 1,4 | 5 | 2 | 8 |

| Q32 | Q33 | Q34 | Q35 | Q36 | Q37 |
|---|---|---|---|---|---|
| If yes, how long after your symptoms did you have your first mammogram? | Was the mammogram able to detect the ILC? | Did you undergo additional testing? (Select all that apply) - Selected Choice | How long after your initial mammogram did you undergo these additional tests? | Were you undergoing active surveillance for breast cancer at the time of your metastatic ILC diagnosis? | If yes, what type of surveillance? (Select all that apply) - Selected Choice |
| 1 | 2 | 1,2,4 | 2 | 1 | |
| 1 | 2 | 1,2,3,4 | 3 | 2 | 1,3,4,7 |
| | 1 | 1,4 | 1 | 1 | |
| | 2 | 4 | 1 | 1 | 8 |
| | 3 | 2,4,5 | 6 | 1 | |

| Q32 | Q33 | Q34 | Q35 | Q36 | Q37 |
| --- | --- | --- | --- | --- | --- |
| If yes, how long after your symptoms did you have your first mammogram? | Was the mammogram able to detect the ILC? | Did you undergo additional testing? (Select all that apply) - Selected Choice | How long after your initial mammogram did you undergo these additional tests? | Were you undergoing active surveillance for breast cancer at the time of your metastatic ILC diagnosis? | If yes, what type of surveillance? (Select all that apply) - Selected Choice |
| 4 | 2 | 1,4,5 | 4 | 2 | 1,2,3,4,5,6,7 |
| | | 1,2,4,5 | 1 | 2 | 3,6 |
| | 2 | | 1 | 1 | |
| | | 4 | | 1 | |
| | 3 | 2,4 | 5 | 1 | |
| | | | | 1 | |
| | 3 | 2,4 | 6 | 2 | 1,2,4,6 |

| Q32 | Q33 | Q34 | Q35 | Q36 | Q37 |
|---|---|---|---|---|---|
| If yes, how long after your symptoms did you have your first mammogram? | Was the mammogram able to detect the ILC? | Did you undergo additional testing? (Select all that apply) - Selected Choice | How long after your initial mammogram did you undergo these additional tests? | Were you undergoing active surveillance for breast cancer at the time of your metastatic ILC diagnosis? | If yes, what type of surveillance? (Select all that apply) - Selected Choice |
| | 3 | 5 | 6 | 1 | |
| 1 | 1 | 1,2,4,5 | 1 | 1 | |
| 2 | 1 | 1,2,3,4,5 | 2 | 2 | 3 |
| 5 | 2 | 4 | 5 | 2 | 3 |
| | | | | 2 | 1,5,8 |

| Q32 | Q33 | Q34 | Q35 | Q36 | Q37 |
|---|---|---|---|---|---|
| If yes, how long after your symptoms did you have your first mammogram? | Was the mammogram able to detect the ILC? | Did you undergo additional testing? (Select all that apply) - Selected Choice | How long after your initial mammogram did you undergo these additional tests? | Were you undergoing active surveillance for breast cancer at the time of your metastatic ILC diagnosis? | If yes, what type of surveillance? (Select all that apply) - Selected Choice |
| 1 | 1 | 2 | 1 | 2 | 6 |
| 1 | 2 | 1,2,4,5 | 3 | 2 | 3 |
|  | 2 | 2,3 | 2 | 2 | 1,2 |
| 1 | 2 | 1,2,3,4 | 2 | 1 |  |
|  | 2 | 2,4 | 2 | 1 |  |
| 1 | 2 | 2,3 | 1 | 2 | 6,7 |
|  | 3 | 4,5 | 2 | 1 |  |
|  | 3 | 6 | 6 | 1 |  |

| Q32 | Q33 | Q34 | Q35 | Q36 | Q37 |
|---|---|---|---|---|---|
| If yes, how long after your symptoms did you have your first mammogram? | Was the mammogram able to detect the ILC? | Did you undergo additional testing? (Select all that apply) - Selected Choice | How long after your initial mammogram did you undergo these additional tests? | Were you undergoing active surveillance for breast cancer at the time of your metastatic ILC diagnosis? | If yes, what type of surveillance? (Select all that apply) - Selected Choice |
| | 3 | 1,4,5 | 5 | 1 | |
| | 2 | 1,2,4 | 4 | 1 | |
| 5 | 2 | 6 | | 1 | |
| | 2 | 2,4,5 | 1 | 2 | 1,2,3,6 |

| Q32 | Q33 | Q34 | Q35 | Q36 | Q37 |
|---|---|---|---|---|---|
| **If yes, how long after your symptoms did you have your first mammogram?** | **Was the mammogram able to detect the ILC?** | **Did you undergo additional testing? (Select all that apply) - Selected Choice** | **How long after your initial mammogram did you undergo these additional tests?** | **Were you undergoing active surveillance for breast cancer at the time of your metastatic ILC diagnosis?** | **If yes, what type of surveillance? (Select all that apply) - Selected Choice** |
| 2 | 2 | 1 | 1 | 2 | 1,2,5 |
| | | 6 | 3 | 1 | |
| | 2 | 1 | 4 | 1 | |
| | | | | 1 | |
| | 1 | 1,2,4 | 1 | 1 | |
| | 1 | 1,2,3,4,5 | 2 | 2 | 3 |
| | 2 | 1,2 | | 2 | 3,4,5 |

| Q32 | Q33 | Q34 | Q35 | Q36 | Q37 |
|---|---|---|---|---|---|
| If yes, how long after your symptoms did you have your first mammogram? | Was the mammogram able to detect the ILC? | Did you undergo additional testing? (Select all that apply) - Selected Choice | How long after your initial mammogram did you undergo these additional tests? | Were you undergoing active surveillance for breast cancer at the time of your metastatic ILC diagnosis? | If yes, what type of surveillance? (Select all that apply) - Selected Choice |
| | 2 | 1,2,4 | 1 | 1 | |
| | 1 | 1,2 | 1 | 1 | |
| 3 | 2 | 6 | | 1 | |

| Q32 | Q33 | Q34 | Q35 | Q36 | Q37 |
|---|---|---|---|---|---|
| **If yes, how long after your symptoms did you have your first mammogram?** | **Was the mammogram able to detect the ILC?** | **Did you undergo additional testing? (Select all that apply) - Selected Choice** | **How long after your initial mammogram did you undergo these additional tests?** | **Were you undergoing active surveillance for breast cancer at the time of your metastatic ILC diagnosis?** | **If yes, what type of surveillance? (Select all that apply) - Selected Choice** |
| | 3 | 1,2,4 | 1 | 1 | 5 |
| | | | | 1 | |
| | | | | 2 | 6 |
| | 2 | 1,4 | 1 | 1 | |
| 1 | 2 | 1,2,4,5 | 1 | 1 | |
| | | | | 1 | |
| 2 | 2 | 2,4 | 3 | 1 | |

| Q32 | Q33 | Q34 | Q35 | Q36 | Q37 |
|---|---|---|---|---|---|
| If yes, how long after your symptoms did you have your first mammogram? | Was the mammogram able to detect the ILC? | Did you undergo additional testing? (Select all that apply) - Selected Choice | How long after your initial mammogram did you undergo these additional tests? | Were you undergoing active surveillance for breast cancer at the time of your metastatic ILC diagnosis? | If yes, what type of surveillance? (Select all that apply) - Selected Choice |
| 1 | 2 | 1,2,3,4 | 1 | 1 | |
| 2 | 2 | 6 | 6 | 2 | 3,6 |
| | 2 | 5 | 1 | 1 | |
| 5 | 3 | 2,4 | 6 | 1 | |
| 2 | 1 | 1,2,4 | 1 | 2 | 6 |
| | | 2 | 5 | 2 | 8 |

| Q32 | Q33 | Q34 | Q35 | Q36 | Q37 |
| --- | --- | --- | --- | --- | --- |
| If yes, how long after your symptoms did you have your first mammogram? | Was the mammogram able to detect the ILC? | Did you undergo additional testing? (Select all that apply) - Selected Choice | How long after your initial mammogram did you undergo these additional tests? | Were you undergoing active surveillance for breast cancer at the time of your metastatic ILC diagnosis? | If yes, what type of surveillance? (Select all that apply) - Selected Choice |
| 2 | 1 | 1,2,4 | 2 | 1 | |
| 5 | 3 | 6 | | 2 | 3 |
| | 1 | 1,4 | 1 | 1 | |

| Q32 | Q33 | Q34 | Q35 | Q36 | Q37 |
|---|---|---|---|---|---|
| If yes, how long after your symptoms did you have your first mammogram? | Was the mammogram able to detect the ILC? | Did you undergo additional testing? (Select all that apply) - Selected Choice | How long after your initial mammogram did you undergo these additional tests? | Were you undergoing active surveillance for breast cancer at the time of your metastatic ILC diagnosis? | If yes, what type of surveillance? (Select all that apply) - Selected Choice |
| | 3 | | | 2 | 1,2,6 |
| | | 6 | 6 | 1 | |
| | 3 | 6 | 6 | 2 | 6 |
| | 1 | 1,2,3,4,5 | 2 | 2 | 2,3,6 |
| | | 4,5 | 2 | 1 | |
| | 1 | 1,3,4 | 1 | 1 | |
| | 2 | 2 | 3 | 1 | |
| | | | | 1 | |

| Q32 | Q33 | Q34 | Q35 | Q36 | Q37 |
|---|---|---|---|---|---|
| **If yes, how long after your symptoms did you have your first mammogram?** | **Was the mammogram able to detect the ILC?** | **Did you undergo additional testing? (Select all that apply) - Selected Choice** | **How long after your initial mammogram did you undergo these additional tests?** | **Were you undergoing active surveillance for breast cancer at the time of your metastatic ILC diagnosis?** | **If yes, what type of surveillance? (Select all that apply) - Selected Choice** |
| | | | 1 | 1 | |
| 1 | 1 | 1,2,3,4 | 1 | 2 | 1,2,3,4,5 |
| | 2 | 1,2,4 | | 2 | 3 |
| 3 | 2 | 1,2,4 | 2 | 1 | |

| Q32 | Q33 | Q34 | Q35 | Q36 | Q37 |
|---|---|---|---|---|---|
| If yes, how long after your symptoms did you have your first mammogram? | Was the mammogram able to detect the ILC? | Did you undergo additional testing? (Select all that apply) - Selected Choice | How long after your initial mammogram did you undergo these additional tests? | Were you undergoing active surveillance for breast cancer at the time of your metastatic ILC diagnosis? | If yes, what type of surveillance? (Select all that apply) - Selected Choice |
| 1 | 1 | 1,2 | 1 | 1 | |
| 4 | 2 | 2,5 | 4 | 1 | |
| 2 | 1 | 1,4 | 1 | 1 | |
| | | | | 1 | |
| 1 | 2 | 1 | 1 | 2 | 1,2,3,4,5,6 |
| 2 | 2 | 1,2,4 | 1 | 1 | |
| | | 5 | | 1 | |

| Q32 | Q33 | Q34 | Q35 | Q36 | Q37 |
|---|---|---|---|---|---|
| If yes, how long after your symptoms did you have your first mammogram? | Was the mammogram able to detect the ILC? | Did you undergo additional testing? (Select all that apply) - Selected Choice | How long after your initial mammogram did you undergo these additional tests? | Were you undergoing active surveillance for breast cancer at the time of your metastatic ILC diagnosis? | If yes, what type of surveillance? (Select all that apply) - Selected Choice |
| 1 | 2 | 1,5 | 5 | 1 | |
| 1 | 2 | 4 | 1 | 2 | 1,3,4,6 |
| | | | | 1 | |
| | | | | 2 | 3,4 |

| Q32 | Q33 | Q34 | Q35 | Q36 | Q37 |
| --- | --- | --- | --- | --- | --- |
| If yes, how long after your symptoms did you have your first mammogram? | Was the mammogram able to detect the ILC? | Did you undergo additional testing? (Select all that apply) - Selected Choice | How long after your initial mammogram did you undergo these additional tests? | Were you undergoing active surveillance for breast cancer at the time of your metastatic ILC diagnosis? | If yes, what type of surveillance? (Select all that apply) - Selected Choice |
| 1 | 1 | 4 | 1 | 1 | |

| Q32 | Q33 | Q34 | Q35 | Q36 | Q37 |
|---|---|---|---|---|---|
| If yes, how long after your symptoms did you have your first mammogram? | Was the mammogram able to detect the ILC? | Did you undergo additional testing? (Select all that apply) - Selected Choice | How long after your initial mammogram did you undergo these additional tests? | Were you undergoing active surveillance for breast cancer at the time of your metastatic ILC diagnosis? | If yes, what type of surveillance? (Select all that apply) - Selected Choice |
| | | 5 | 6 | 2 | 3,5 |

| Q32 | Q33 | Q34 | Q35 | Q36 | Q37 |
| --- | --- | --- | --- | --- | --- |
| If yes, how long after your symptoms did you have your first mammogram? | Was the mammogram able to detect the ILC? | Did you undergo additional testing? (Select all that apply) - Selected Choice | How long after your initial mammogram did you undergo these additional tests? | Were you undergoing active surveillance for breast cancer at the time of your metastatic ILC diagnosis? | If yes, what type of surveillance? (Select all that apply) - Selected Choice |

| Q32 | Q33 | Q34 | Q35 | Q36 | Q37 |
|---|---|---|---|---|---|
| If yes, how long after your symptoms did you have your first mammogram? | Was the mammogram able to detect the ILC? | Did you undergo additional testing? (Select all that apply) - Selected Choice | How long after your initial mammogram did you undergo these additional tests? | Were you undergoing active surveillance for breast cancer at the time of your metastatic ILC diagnosis? | If yes, what type of surveillance? (Select all that apply) - Selected Choice |

| Q32 | Q33 | Q34 | Q35 | Q36 | Q37 |
|---|---|---|---|---|---|
| If yes, how long after your symptoms did you have your first mammogram? | Was the mammogram able to detect the ILC? | Did you undergo additional testing? (Select all that apply) - Selected Choice | How long after your initial mammogram did you undergo these additional tests? | Were you undergoing active surveillance for breast cancer at the time of your metastatic ILC diagnosis? | If yes, what type of surveillance? (Select all that apply) - Selected Choice |
| | | | | 1 | |
| | 3 | 6 | 6 | 2 | 3,6 |
| | 2 | 1,2,4,5 | | 2 | 2,4,5 |

| Q32 | Q33 | Q34 | Q35 | Q36 | Q37 |
| --- | --- | --- | --- | --- | --- |
| If yes, how long after your symptoms did you have your first mammogram? | Was the mammogram able to detect the ILC? | Did you undergo additional testing? (Select all that apply) - Selected Choice | How long after your initial mammogram did you undergo these additional tests? | Were you undergoing active surveillance for breast cancer at the time of your metastatic ILC diagnosis? | If yes, what type of surveillance? (Select all that apply) - Selected Choice |
| | | | | 2 | 2,3,6 |
| 1 | 2 | 1 | 1 | 1 | |
| 5 | 3 | 6 | 6 | 2 | 6,8 |
| | | | | 1 | 3 |

| Q32 | Q33 | Q34 | Q35 | Q36 | Q37 |
|---|---|---|---|---|---|
| If yes, how long after your symptoms did you have your first mammogram? | Was the mammogram able to detect the ILC? | Did you undergo additional testing? (Select all that apply) - Selected Choice | How long after your initial mammogram did you undergo these additional tests? | Were you undergoing active surveillance for breast cancer at the time of your metastatic ILC diagnosis? | If yes, what type of surveillance? (Select all that apply) - Selected Choice |
|  | 3 | 2,4 | 6 | 1 |  |

| Q32 | Q33 | Q34 | Q35 | Q36 | Q37 |
|---|---|---|---|---|---|
| If yes, how long after your symptoms did you have your first mammogram? | Was the mammogram able to detect the ILC? | Did you undergo additional testing? (Select all that apply) - Selected Choice | How long after your initial mammogram did you undergo these additional tests? | Were you undergoing active surveillance for breast cancer at the time of your metastatic ILC diagnosis? | If yes, what type of surveillance? (Select all that apply) - Selected Choice |
| | | | | 2 | 1,2,3,4,5,8 |
| 2 | 2 | 1,2,3,4 | 1 | 2 | 4,8 |

| Q32 | Q33 | Q34 | Q35 | Q36 | Q37 |
|---|---|---|---|---|---|
| If yes, how long after your symptoms did you have your first mammogram? | Was the mammogram able to detect the ILC? | Did you undergo additional testing? (Select all that apply) - Selected Choice | How long after your initial mammogram did you undergo these additional tests? | Were you undergoing active surveillance for breast cancer at the time of your metastatic ILC diagnosis? | If yes, what type of surveillance? (Select all that apply) - Selected Choice |
| 1 | 1 | 1,2,4 | 1 | 1 | |
| 5 | 2 | 1,2,3,4 | 3 | 2 | |

| Q32 | Q33 | Q34 | Q35 | Q36 | Q37 |
|---|---|---|---|---|---|
| If yes, how long after your symptoms did you have your first mammogram? | Was the mammogram able to detect the ILC? | Did you undergo additional testing? (Select all that apply) - Selected Choice | How long after your initial mammogram did you undergo these additional tests? | Were you undergoing active surveillance for breast cancer at the time of your metastatic ILC diagnosis? | If yes, what type of surveillance? (Select all that apply) - Selected Choice |
| | | 4 | 1 | 1 | |
| | 1 | 1,4 | 1 | 2 | 6 |
| | | 6 | | 1 | |
| | | | 1 | 1 | |

| Q32 | Q33 | Q34 | Q35 | Q36 | Q37 |
|---|---|---|---|---|---|
| If yes, how long after your symptoms did you have your first mammogram? | Was the mammogram able to detect the ILC? | Did you undergo additional testing? (Select all that apply) - Selected Choice | How long after your initial mammogram did you undergo these additional tests? | Were you undergoing active surveillance for breast cancer at the time of your metastatic ILC diagnosis? | If yes, what type of surveillance? (Select all that apply) - Selected Choice |
| 3 | 2 | 2,3 | 3 | 2 | 3 |

| Q32 | Q33 | Q34 | Q35 | Q36 | Q37 |
|---|---|---|---|---|---|
| If yes, how long after your symptoms did you have your first mammogram? | Was the mammogram able to detect the ILC? | Did you undergo additional testing? (Select all that apply) - Selected Choice | How long after your initial mammogram did you undergo these additional tests? | Were you undergoing active surveillance for breast cancer at the time of your metastatic ILC diagnosis? | If yes, what type of surveillance? (Select all that apply) - Selected Choice |
| 1 | 2 | 2,4 | 1 | 2 | 5,6,8 |
| | | | | 2 | 5 |
| | | 6 | 6 | 2 | 8 |
| | 3 | | | 2 | 3,5,6 |
| | 3 | | | 2 | 3,5,6 |

| Q32 | Q33 | Q34 | Q35 | Q36 | Q37 |
|---|---|---|---|---|---|
| If yes, how long after your symptoms did you have your first mammogram? | Was the mammogram able to detect the ILC? | Did you undergo additional testing? (Select all that apply) - Selected Choice | How long after your initial mammogram did you undergo these additional tests? | Were you undergoing active surveillance for breast cancer at the time of your metastatic ILC diagnosis? | If yes, what type of surveillance? (Select all that apply) - Selected Choice |
| 2 | 3 | 1,2,3,4 | 6 | 2 | 3,4,8 |
| 1 | | 2 | 1 | 2 | 4,6 |
| | | 6 | | 2 | 2,4,6 |

1

1

1
