## Supplemental Table 4, ILC misdiagnosis patient survey free text examples for "Quantitative and qualitative patient-reported analysis of misdiagnosis and/or late diagnosis of metastatic lobular cancer"

### mILC Free Text summary - examples

| **GI** | |
|---|---|
| **Theme** | **Description** |
| **Progression to Stage 4 / Metastasis due to Delay** | Several patients reported that misdiagnosis or delayed diagnosis led to cancer progressing to metastatic disease (e.g., bones, abdomen, peritoneum). |
| **Initial Symptoms Misattributed / Misdiagnosed** | GI symptoms were not initially linked to breast cancer. Some patients reported they were told it was ME/CFS, IBS, or other unrelated conditions. |
| **Lack of Imaging or Inadequate Imaging Interpretation** | Imaging results were misread or not followed up (e.g., mammograms, MRIs), contributing to missed diagnoses. |
| **Biopsy Delays / Missed Biopsies** | Biopsies were delayed or required multiple attempts to identify the cancer. |
| **Dismissive Clinical Response / Not Believed** | Patients experienced medical dismissal or lack of investigation despite persistent symptoms. |
| **Miscommunication / Inadequate Patient Education** | Missed opportunities to educate patients on symptoms of metastatic disease. |
| **Impact on Life Quality / Emotional Toll** | Includes stress-induced medical events (e.g., stroke), caregiving challenges, or emotional burden. |
| **Systemic Healthcare Delays (esp. rural)** | Rural or under-resourced healthcare settings caused diagnostic delays. |
| **Long-Term Diagnostic Journey / Multiple Years of Delay** | Some patients described a multi-year delay before receiving correct diagnosis. |

| **GU** | |
|---|---|
| **Theme** | **Description** |
| **Progression to Stage 4 / Metastasis due to Delay** | Patients noted that delayed or missed diagnoses led to metastasis, often to bones or abdomen. |
| **Initial Symptoms Dismissed / Misattributed (Especially GU-specific)** | Symptoms like urinary frequency or discomfort were dismissed or incorrectly attributed (e.g., “cleaning too vigorously”). |
| **Delay in Testing / Treatment Initiation** | Multiple examples of significant delay in diagnosis, treatment, or access to advanced imaging/tests. |
| **Stress-Related Health Impact** | The emotional toll and stress from delays resulted in physical consequences. |
| **Patient-Led Escalation / Self-Advocacy** | Patients often had to push for proper testing, imaging, or referral themselves. |
| **Misdiagnosis Between IDC vs ILC** | Some patients reported initial diagnosis as IDC despite ILC findings in pathology reports. |
| **Missed Preventive Opportunities / Regret** | Some respondents felt treatment or surgeries (e.g., mastectomy) could have been avoided. |
| **Healthcare Access Disparities / System Gaps** | Barriers like being refused tests at major centers, but accepted at others highlight care inconsistency. |
| **Unclear / No Impact** | One respondent reported “No,” and one was ambiguous |

#### mILC Free Text summary - examples

| neurologic | |
|---|---|
| **Theme** | **Description** |
| **Delayed Diagnosis and/or Treatment** | Multiple patients reported delays in diagnosis or initiation of treatment (e.g., 5 months, 6 months, or even over 15 years). These delays were often due to missed imaging findings or dismissal of symptoms. |
| **Diagnostic Imaging Failure (Especially Mammogram)** | Several respondents explicitly noted that mammograms failed to detect ILC, contributing to delays in diagnosis and treatment. |
| **Patient Self-Advocacy Was Required** | Patients had to push for biopsies, scans, or second opinions to receive correct diagnosis and care. |
| **Systemic Misdiagnosis / Misattribution of Symptoms** | Misdiagnoses included ME/CFS, gastrointestinal causes, or assumption of IDC instead of ILC. |
| **Stress-Related Health Consequences** | One patient experienced a minor stroke, explicitly attributing it to stress from delayed or inadequate care. |
| **Metastasis Progression Due to Delay** | Some patients noted that by the time of accurate diagnosis, disease had already become metastatic (e.g., skull metastasis, stomach spread). |
| **Failure to Recognize ILC Pattern / Inadequate Testing** | Cases where lobular histology was missed or ignored initially, resulting in misaligned treatment decisions. |
| **Treatment Refusal or Access Barrier** | One patient was refused advanced diagnostics |

| hematological | |
|---|---|
| **Theme** | **Description** |
| **Delayed Diagnosis and/or Treatment** | Several patients reported delays ranging from 5 months to 6 years. This allowed the cancer to progress, often from bones to stomach or other areas. |
| **Missed or Dismissed Early Symptoms** | Some individuals sought care early (e.g., for back pain, bone issues), but symptoms were ignored or not linked to cancer until later. |
| **Imaging/Diagnostic Failure** | Similar to other groups, imaging findings (e.g., MRI bone lesions) were either misinterpreted or not acted upon, delaying treatment. |
| **Patient Self-Advocacy or Seeking Second Opinion** | Patients had to see many doctors or switch institutions to get a proper diagnosis |
| **Misclassification: IDC vs. ILC** | Pathology showed ILC but initial diagnosis treated as IDC, impacting management and interpretation of spread. |
| **Emotional/Physical Toll from Delays** | Stress-related outcomes (e.g., minor stroke), chronic pain, and emotional distress were frequently reported. |
| **Missed Treatment Opportunities or Access Barriers** | Several respondents mentioned being denied specific therapies or tests |
| **Stress-Related Health Events** | One patient again noted a stress-induced stroke, echoing a recurring theme across subgroups. |
| **Systemic Gaps in Care/Recognition** | Difficulty getting metastasis confirmed despite bone lesions; ambiguity in linking symptoms to cancer progression. |
| **Unclear or No Impact** | One patient felt the symptoms (back pain) led to early detection. |
| **Treatment Delay Consequences** | Some explicitly mentioned metastasis spread or permanent pain as a result of delay. |
